# Effectiveness of Noninvasive Brain Stimulation Protocols on Drug Craving and Consumption/Relapse in Substance Use Disorders: A Systematic Review and Meta-analysis of 208 Clinical Trials and 36 Protocols

**DOI:** 10.1101/2025.09.21.25335559

**Authors:** Ghazaleh Soleimani, Afra Souki, Sara Honari, Travis E Baker, Andre R Brunoni, Mohsen Ebrahimi, Eduardo A Garza-Villarreal, Tony P George, Rita Z Goldstein, Manish Kumar Jha, Tonisha Kearney-Ramos, Rayus Kuplicki, Bernard Le Foll, Kelvin O Lim, Martin P Paulus, Arash Rahmani, Gregory Sahlem, Victor M Tang, Hosna Tavakoli, Alireza Valyan, Ti-Fei Yuan, Mehran Zare-Bidoky, Marom Bikson, Colleen A Hanlon, Michael Nitsche, Hamed Ekhtiari

**Affiliations:** Department of Psychiatry and Behavioral Sciences, University of Minnesota, MN, USA; Department of Biomedical Engineering, University of Minnesota, MN, USA; Department of Psychology, University of Tehran, Tehran, Iran; Psychiatry and Behavioral Sciences Research Center, Mashhad University of Medical Sciences, Mashhad, Iran; Center for Molecular and Behavioral Neuroscience, Rutgers University, Newark, New Jersey 07102; University of São Paulo Medical School, Psychiatry, São Paulo, Brazil; Iranian National Center for Addiction Studies (INCAS), Tehran, Iran; Institute of Neurobiology, Universidad Nacional de México campus Juriquilla, Queretaro, Mexico; Department of Psychiatry, Temerty Faculty of Medicine, University of Toronto, Toronto, ON, Canada; Addictions Division, Centre for Addiction and Mental Health (CAMH), Toronto, ON, Canada; Departments of Psychiatry and Neuroscience, Icahn School of Medicine at Mount Sinai, New York City, New York; Department of Psychiatry, University of Texas Southwestern Medical Center, Dallas, TX, USA; Department of Psychiatry & Behavioral Sciences, Duke University Medical Center, Durham, NC 27710; Laureate Institute for Brain Research (LIBR), OK, USA; Department of Psychiatry, University of Toronto, Toronto, ON, Canada; Brain Stimulation Lab, Department of Psychiatry & Behavioral Sciences, Stanford School of Medicine, Stanford, CA, USA; Addictions Division, Centre for Addiction and Mental Health, Toronto, ON, Canada; Institute for Cognitive Sciences Studies (ICSS), Tehran, Iran; Allameh Tabataba’i University, Tehran, Iran; Shanghai Key laboratory of Psychotic disorders, Shanghai Mental Health Center, Shanghai Jiao Tong University School of Medicine, Shanghai, China; Iranian National Center for Addiction Studies, Tehran University of Medical Sciences, Tehran, Iran; Department of Biomedical Engineering, The City College of New York, New York, NY, USA; Department of Cancer Biology, Wake Forest University School of Medicine, NC, USA; BrainsWay, Burlington, MA, USA; Department of Psychology and Neurosciences, Leibniz Research Center for Working Environment and Human Factors, Dortmund, Germany; University Clinic of Psychiatry and Psychotherapy, Protestant Hospital of Bethel Foundation, University Hospital OWL, Bielefeld University, Bielefeld, Germany; German Center for Mental Health (DZPG), Bochum, Germany

**Keywords:** transcranial electrical stimulation, transcranial magnetic stimulation, addiction psychiatry, systematic review, meta-analysis, substance use disorders, craving, relapse

## Abstract

**Background:** Transcranial Magnetic, Electrical, and Focused-Ultrasound Stimulation (TMS/tES/tFUS) are major noninvasive brain stimulation (NIBS) techniques used to treat various psychiatric disorders, including substance use disorders (SUDs). Although NIBS with varying stimulation parameters shows promising effects on drug-related behaviors such as craving and consumption/relapse, the question of which protocol is most effective remains unresolved.

**Method:** To address this gap, we conducted a living systematic review and meta-analysis to quantify the effects of TMS/tES/tFUS on SUD. Controlled trials of TMS/tES/tFUS for all types of SUD were selected up to January 1, 2024.

**Findings:** The final systematic review included 208 trials (121 TMS, 86 tES, 1 tFUS), with 116 randomized sham-controlled trials (59 TMS, 57 tES) eligible for meta-analysis due to a low risk of bias. Data from 5,106 participants in active and 4,914 in sham groups were analyzed. TMS showed medium effects on craving (g = 0.52, 95% CI: 0.29-0.75, p < 0.001, I^2 = 89.36) and consumption (g = 0.41, CI: 0.26-0.56, p < 0.001, I^2 = 61.56). tES showed a medium effect on craving (g = 0.40, CI: 0.25-0.55, p < 0.001, I^2 = 60.69) and a small effect on consumption (g = 0.27, CI: 0.15-0.38, p = 0.013, I^2 = 22.67). Among the 36 different protocols examined, subgroup analyses identified the strongest effect for reducing both craving and consumption with high-frequency deep TMS using the H4 coil (single study) (g = 3.92 and 1.12, respectively, p < 0.001), with a maximum electric field over the frontopolar cortex. This effect was followed by high-frequency rTMS over the left DLPFC (g = 0.66 and 0.52, respectively, p < 0.05), as well as bilateral anodal-right cathodal-left (g = 0.49 and 0.42, respectively, p < 0.0001) and anodal-left cathodal-right (g = 0.38 and 0.31, respectively, p < 0.05) tES with direct current (tDCS) over the DLPFC, with maximum electric field on the frontopolar cortex.

**Interpretation:** Our results provide evidence that TMS and tES stimulation over frontopolar and DLPFC regions produce medium to large effect sizes in reducing drug craving and consumption/relapse in SUD. While requiring further replication in future studies, these findings highlight the promise of these interventions.

## 1. Introduction

Substance use disorders (SUDs) are major global health challenges, affecting millions of individuals worldwide and contributing significantly to morbidity and mortality. Alcohol use disorder affects approximately 5% of the global population (∼400 million people),^1^ and other SUDs impact about 0.7% of the population (∼36 million worldwide).^2^ These conditions impose a substantial burden on individuals, families, societies, and healthcare systems, highlighting the urgent need for effective global prevention and treatment strategies. Numerous compounds have undergone scrutiny as potential pharmacotherapies for SUDs. However, as of early 2025, only a limited number of medications have demonstrated robust efficacy (consistent, reproducible, and clinically durable) and gained approval from the United States Food and Drug Administration (FDA).^3^ However, for some SUDs, such as methamphetamine use disorder (MUD) and cannabis use disorder (CUD), there are currently no FDA-approved pharmacological treatments, leaving clinicians with limited options beyond behavioraltherapies.

The significant global burden of SUDs, combined with their neurotoxic and neuroadaptive effects on the brain, underscores a critical treatment gap due to the lack of effective pharmacotherapies. The high rates of relapse and ongoing health consequences in individuals with SUDs emphasize the pressing need for novel therapeutic approaches. Neuromodulation, particularly non-invasive brain stimulation (NIBS), has emerged as a promising alternative intervention, targeting the pathophysiological mechanisms underlying SUD.^4^ In this regard, transcranial electrical (tES), magnetic (TMS), and focused ultrasound (tFUS) stimulation have shown promise as potential treatments for SUDs.^5,6^ NIBS techniques employ varied stimulation protocols, including different frequencies, intensities, and target areas, to modulate brain activity and support therapeutic outcomes. Treatment outcomes in NIBS studies commonly include reduced drug craving, enhanced self-regulation, with key measures often assessing both short-term drug consumption and longer-term relapse prevention.

A major focus of recent NIBS research has been the application of anodal tES or high-frequency TMS to the dorsolateral prefrontal cortex (DLPFC), a region commonly targeted in depression protocols to enhance cortical excitability. This interest is driven by overlapping DLPFC-mediated executive function deficits observed in both depression and SUDs.^7^ However, there are multiple trials targeting other sites, and the optimal site(s) and protocols for effective addiction treatment remain unclear.^8^ To address this gap, we conducted a living systematic review and meta-analysis with three primary aims: (1) to characterize the heterogeneity of NIBS parameter spaces used in SUD research; (2) to quantify the overall effect size of NIBS on drug craving and consumption/relapse, and evaluate how stimulation parameters influence outcomes; and (3) to identify the most effective stimulation protocols. These findings aim to inform the development of more effective interventions and support policy and regulatory decisions in global addiction treatment efforts.

Regarding a recently published review of review papers on (non)invasive methods in the field of SUD, by the end of 2024, there are at least five systematic review papers (with or without meta-analysis) in this area.^9,10^ One of the most recent and comprehensive ones was published in 2023 by Mehta et al., which covered studies up to the end of October 2023 and only reported the meta-analysis for alcohol and nicotine use disorders.^6^ Here, (1) in the systematic review part, we reported results up to the end of 2023, which will be updated annually through the International network of tES and TMS for Addiction Medicine (INTAM) infrastructure,^5^ (2) in the meta-analysis part, we conducted a comprehensive subgroup analysis covering all types of SUDs (rather than focusing on two samples as Mehta et al. did), and (3) we performed subgroup analyses of stimulation protocols specifically to identify the most effective protocol for each stimulation type, including deep and figure-of-eight coil TMS as well as tES studies.

## 2. Method

### 2.1. Search Strategy in the Systematic Review Phase

A comprehensive literature search was conducted in PubMed for studies published up to January 1, 2024, involving tES, TMS, and tFUS as interventions for SUDs. Search terms are detailed in Table S1. The review follows the latest Preferred Reporting Items for Systematic Review and Meta-Analysis Protocols (PRISMA-P).^11^ The initial search identified 508 TMS, 663 tES, and 156 tFUS articles. Two independent investigators (AS, HT) screened and extracted data, with a 3^rd^ reviewer (GS) resolving discrepancies by consulting with the senior author (HE) (see supplementary materials). Extracted data and key figures are available on the INTAM OSF page (https://osf.io/sv8ky) and will be updated annually, with comprehensive results published every five years.

### 2.2. Study Inclusion for the Meta-Analysis Phase

For the meta-analysis, a refined subset of studies was selected from the systematic review dataset. Studies included if they: (1) were randomized controlled trials reporting craving and/or consumption/relapse as outcome measures, and (2) provided sufficient statistical data—such as means and standard deviations (SDs) for pre- and post-intervention, mean change scores, F-values, or relapse rates—to enable effect size estimation for active versus sham stimulation. Studies were excluded if they: (1) lacked a sham-controlled condition, or (2) did not report sufficient data for effect size calculation and failed to provide these data upon request from the first or last author.

### 2.3. Assessment of the Risk of Bias of the Included Studies

Cochrane’s risk-of-bias tool (RoB 2) was used to rate the methodological quality of the included studies.^12^ The five main domains covered by this tool are (1) bias due to the randomization process and timing of identification or recruitment of participants, (2) deviation from intended intervention, (3) missing outcome data, (4) measurement of outcomes, and (5) selection of the reported result. Two authors (AS, SH) independently assessed each item for each study. If there were any unresolved disagreements, these were discussed with a third person (MZB), and the final decision was then made.

### 2.4. Hedges’ g Calculations

Meta-analytic models were implemented using the “metafor” package in R. Effect sizes were calculated by comparing changes in drug craving/consumption/relapses between active stimulation and sham control groups. Hedges’ g was used as the effect size metric, accompanied by 95% confidence intervals (CIs). All analyses were conducted using random-effect models to account for between-study heterogeneity. Positive effect sizes indicated greater reductions in craving, consumption, or relapse following active stimulation compared to sham.

### 2.5. Publication Bias Assessment

To assess the potential impact of publication bias, which could either inflate or deflate effect sizes in meta-analyses,^13,14^ two distinct evaluations were conducted. (1) The original funnel plot was compared with a modified version generated through the application of the trim and fill technique.^15^ (2) Egger’s regression test was implemented, which gauges the degree of funnel plot asymmetry by assessing the intercept resulting from a regression of standard normal deviation against precision.^16^ A P-value for the intercept falling below 0.05 indicated the presence of substantial publication bias across the array of comparisons.

### 2.6. Subgroup Analysis

Pooled effect sizes (small (≈0.2), medium (≈0.5), and large (≈0.8)) were calculated to clarify the effects of different parameters, including substance types, number of sessions, study design (parallel vs crossover), target laterality, stimulation target, stimulation intensity, and stimulation duration. To provide insights about different stimulation protocols, pooled effect sizes were also calculated based on stimulation type (excitatoryvs inhibitory) and stimulation target.

## 3. Results

### 3.1. Systematic Review Phase Results

121 TMS, 86 tES, and 1 tFUS studies met our inclusion criteria (see PRISMA flowchart in Figure 1). Notably, individual studies often included multiple experiments (e.g., targeting different brain regions), resulting in a greater number of experiments than the total number of studies.

**Figure 1.**
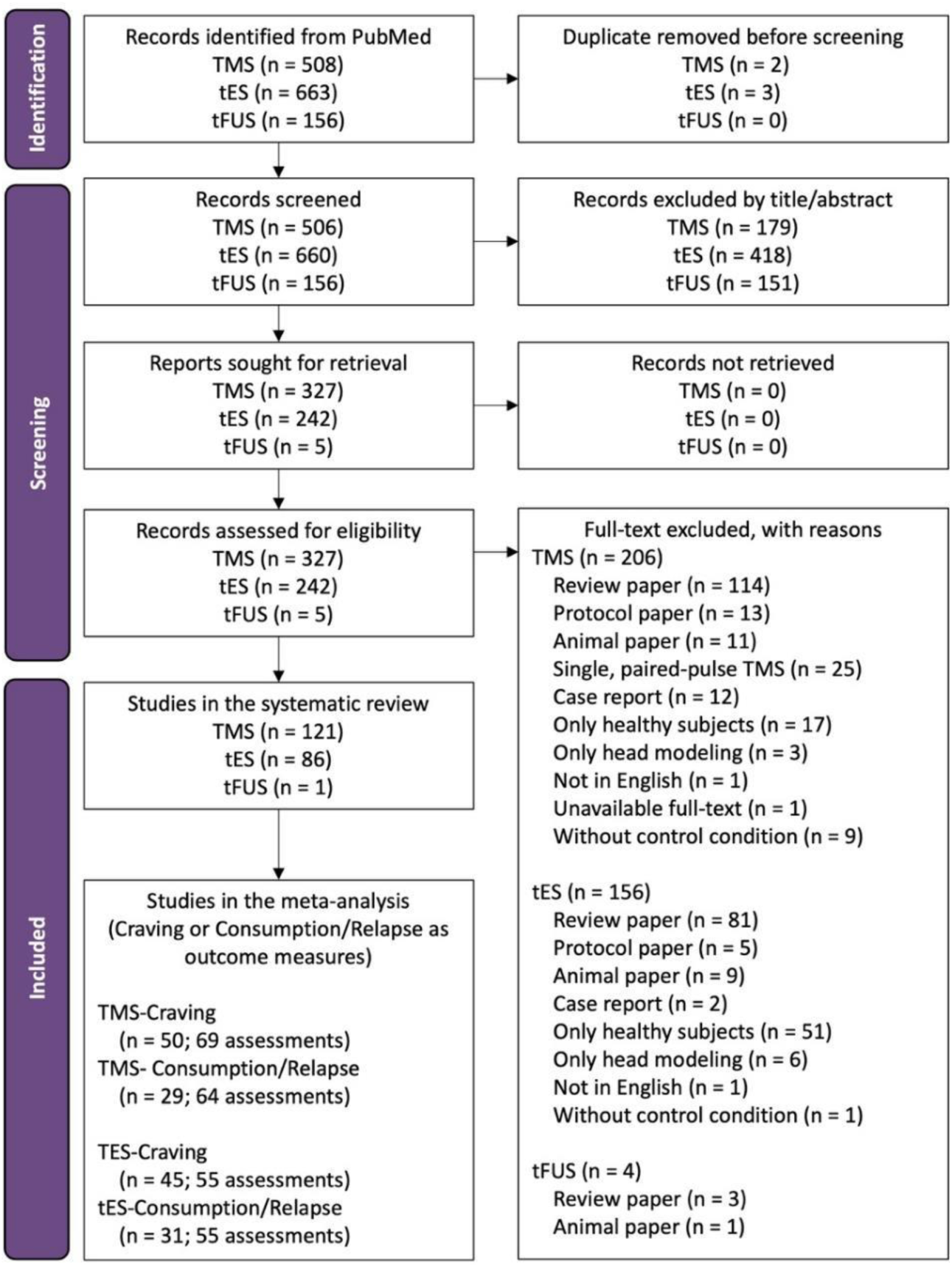
PRISMA Flowcharts for tES, TMS, and tFUS Studies in Substance Use Disorders. This PRISMA flow diagram represents published trials of tES, TMS, and tFUS for individuals with substance use disorders from inception up to 1 January 2024. The last two sections of the PRISMA flow diagram indicate the total number of studies included in the systematic review and meta-analysis (studies that reported craving or consumption/relapse as outcome measures were included in the meta-analysis). Some studies reported both craving and consumption/relapse as outcomes, while others included multiple comparisons (e.g., varying stimulation intensities or control conditions). Consequently, a single publication could contribute multiple assessments/experiments to the analysis. In the systematic review part, “n” refers to the number of publications included, whereas in the meta-analysis part, "n" refers to the total number of assessments (e.g., craving or consumption/relapse assessments). <u>Abbreviations</u>: tES: transcranial electrical stimulation, TMS: transcranial magnetic stimulation, tFUS: transcranial focused ultrasound stimulation. n=13 studies only defined the protocol, and n=3 studies used results from substance use disorders solely for electric field simulations without pre-post measures. Additionally, we were unable to retrieve the full text for one older study.

The included studies were conducted across 21 countries, with the highest contributions from the United States (n=49, 23.7%) and China (n=50, 24.2%). Studies were conducted on different substances, including nicotine (n=62, 28.2%), alcohol (n=60, 27.3%), amphetamines (n=40, 18.2%), cocaine (n=25, 11.4%), opioids (n=25, 11.4%), and cannabis (n=8, 3.6%). The time trend indicates that 60% of the studies were published in the last five years, with a publication peak in 2022 (Figure 2).

**Figure 2.**
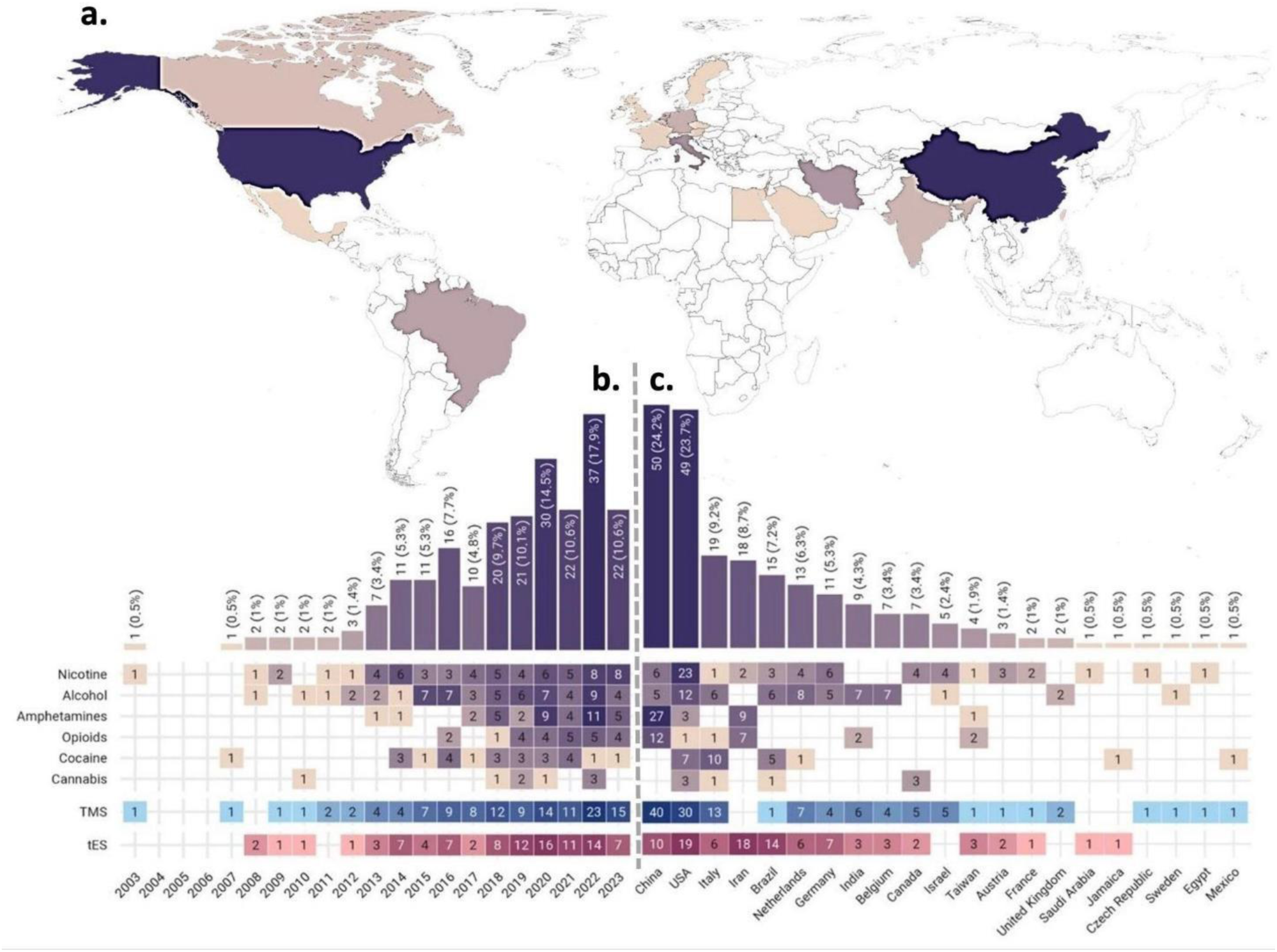
Global Distribution and Trends in tES/TMS Studies for Substance Use Disorders. The figure illustrates the global trends in tES/TMS research on substance use disorders. Colors range from light yellow to purple, with darker shades indicating a higher number of publications. Only papers with full text available in English were included, which may lead to a relative over-representation of English-speaking countries. In the lower panels, studies are color-coded by the type of intervention: tES in red and TMS in blue. (a) Global Map of Study Distribution: A heatmap displays the number of published tES/TMS studies in each country, with darker shades representing higher publication densities. (b) Annual Trends in Publications by Country: This panel presents the number of tES/TMS studies categorized by year of publication, grouped by addictive substance and country. (c) Study Distribution by Substance of Use and Country: This panel categorizes the number of tES/TMS studies in each country based on the type of addictive substance studied. Studies are color-coded by intervention type: tES in red and TMS in blue. Note that some studies included multiple types of addictive substances, resulting in a total count of 220 items, which exceeds the actual number of studies included. <u>Abbreviations</u>: tES: transcranial electrical stimulation; TMS: transcranial magnetic stimulation.

A total of 5,843 individuals received active stimulation (Figure 3), including 4,303 males, 1,200 females, and 340 participants with unspecified sex. Participants represented various substance use conditions: active users/non-treatment-seekers (44%), treatment-seekers/pre-treatment (23%), detoxification (20%), and recovery (14%) phases. All individuals received one or more sessions of active TMS (n=3,740) or tES (n=2,103).

**Figure 3:**
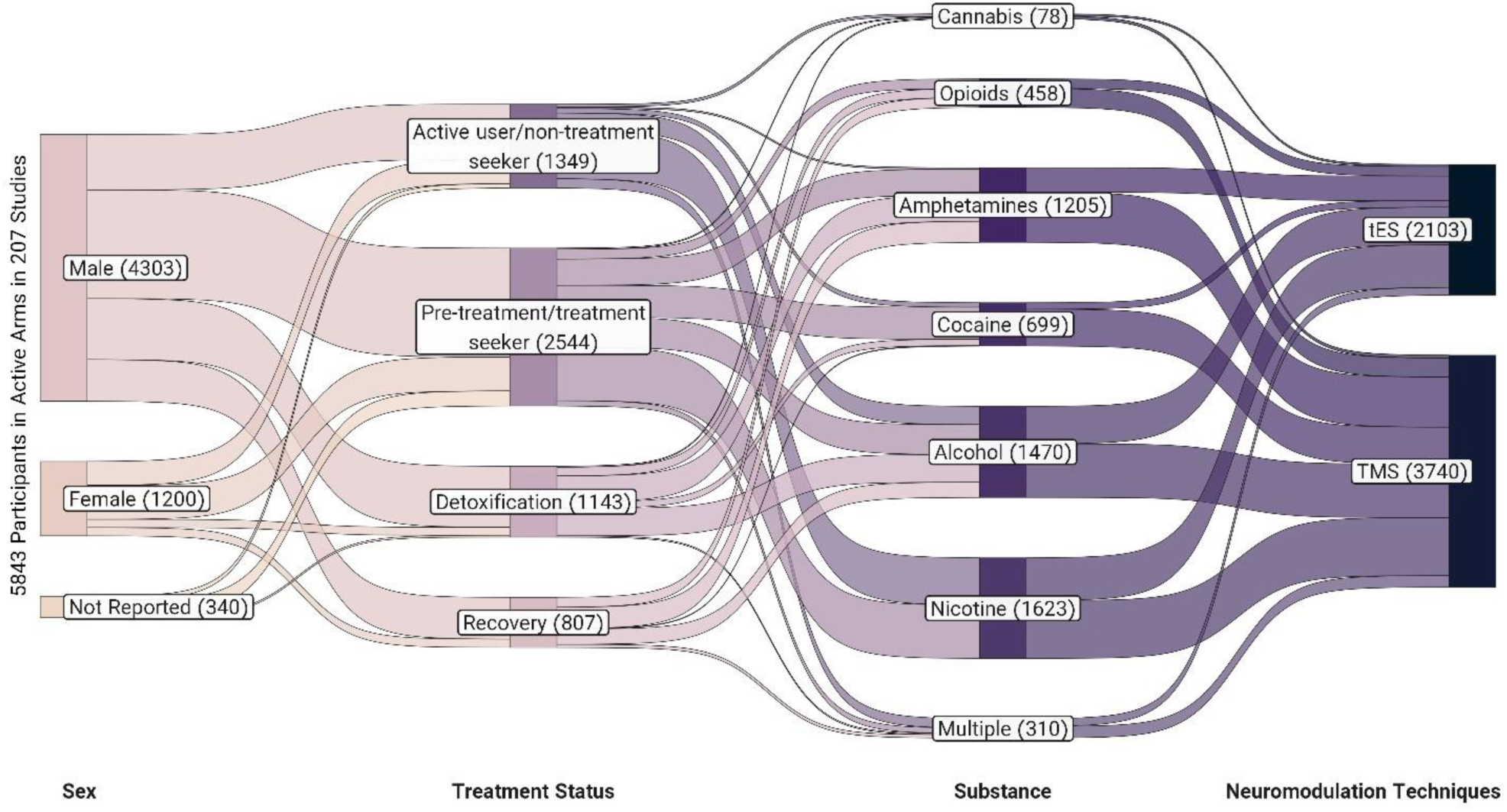
Participant Demographics in tES/TMS Trials for Substance Use Disorders. (Total number of subjects in the active stimulation arm = 5843). This Sankey diagram illustrates the distribution of participants in the active arm, divided by gender, treatment status of their addiction, types of substances used, and types of intervention. The width of the boxes in each column indicates the relative prevalence of each category, while the width of the connecting ribbons represents the proportion of participants shared between categories. Note: ’Multiple’ denotes studies that included participants with various substance use disorders or those that did not specify the dominant substance used by the patients. <u>Abbreviations</u>: tES: transcranial electrical stimulation, TMS: transcranial magnetic stimulation.

Both tES and TMS studies targeted a range of brain regions, categorized into 10 main subregions (See supplementary materials Figure S.1). The majority of experiments focused on the dorsolateral prefrontal cortex (DLPFC, 81.5%; left DLPFC: 52.39%, right DLPFC: 29.11%). Other targets included frontopolar cortex (6.5%), insular cortex (4.4%), inferior frontal gyrus (IFG, 2.9%), frontal-parietal-temporal junction (FPT, 2.2%), anterior cingulate cortex (ACC, 0.7%), cerebellar cortex (0.7%), superior frontal gyrus (SFG, 0.4%), posterior cingulate cortex (PCC, 0.4%), and precuneus (0.4%). Figure 4 visualizes the distribution of TMS and tES experiments by stimulation type—excitatory (anodal tDCS, high-frequency TMS), inhibitory (cathodal tDCS, low-frequency TMS), and tACS—and by target region.

**Figure 4:**
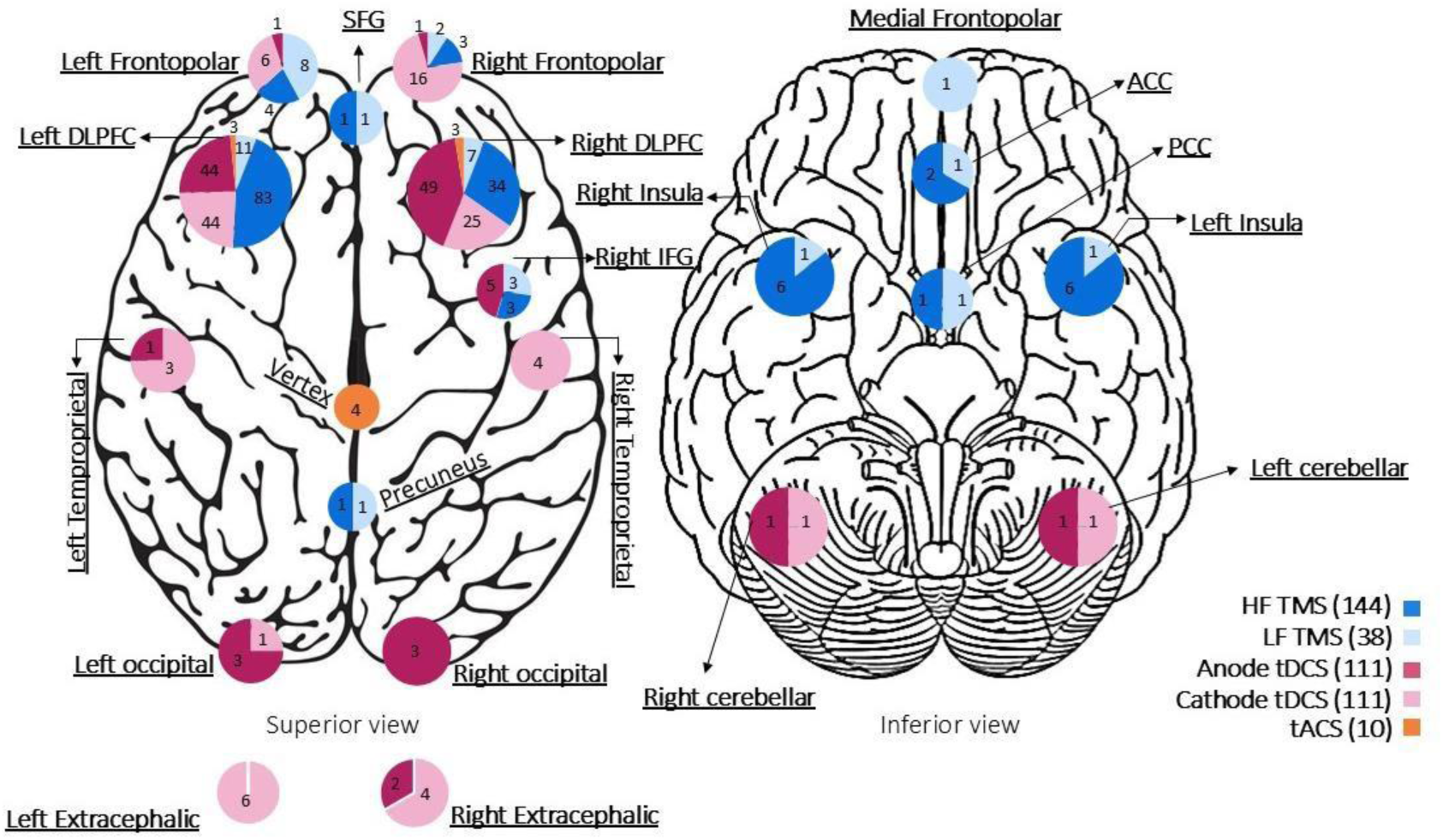
Stimulation targets for tES/TMS trials for substance use disorders. Stimulation targets based on the type of stimulation are shown for the cortical surface, <u>Excitatory</u>: HF TMS and anodal tDCS, tACS, <u>Inhibitory</u>: LF TMS, and cathodal tDCS. The depicted number of cortical locations is equal to 299 and exceeds 207 (the total number of tES/TMS studies in this systematic review) due to the application of multiple stimulation protocols in some studies. Notably, in one tDCS study, an HD electrode with anode and cathode placement over the left DLPFC was employed. In studies investigating bilateral and deep TMS stimulation protocols, as well as in one temporoparietal stimulation study, each reported target was considered as two targets: one in each hemisphere. The term "Frontopolar" collectively refers to the medial prefrontal cortex (mPFC), ventromedial prefrontal cortex (vmPFC), and superior medial frontal cortex (sMFC) regions across the studies. <u>Abbreviations</u>: tES: transcranial electrical stimulation, tDCS: transcranial direct current stimulation, tACS: transcranial alternating stimulation, TMS: transcranial magnetic stimulation, HF TMS: high-frequency TMS (≥ 5 Hz), LF TMS: low-frequency TMS (≤ 1 Hz), SFG: superior frontal gyrus, DLPFC: dorsolateral prefrontal cortex, IFG: inferior frontal gyrus, ACC: anterior cingulate cortex, PCC: posterior cingulate cortex.

Among the 121 TMS studies reviewed, most employed conventional (fixed/not-patterned frequency) repetitive TMS (rTMS, n=75) followed by intermittent theta burst stimulation (iTBS, n=16), continuous theta burst stimulation (cTBS, n=8), deep transcranial magnetic stimulation (dTMS, n=13), or combinations of different TMS types, such as cTBS and iTBS (n=9). Among the 86 tES studies, the vast majority used transcranial direct current stimulation (tDCS, n=83), with only three studies applying transcranial alternating current stimulation (tACS, n=3). Within the tDCS studies, only one study used 4x1 high-definition (HD) electrodes (n=1), while most employed large conventional electrode pads (n=78); electrode type was not reported for seven tDCS studies (supplementary materials Figure S.2). Additional methodological details are presented in supplementary Figures S2–S6, highlighting variability in electrode/coil positioning (most commonly: EEG 10–20 system; n = 112), primary outcome measures (craving: n = 159; consumption/relapse/abstinence: n = 111), and control conditions (most commonly sham-controlled; n = 110). The most frequently used stimulation parameters were 2 mA intensity for tES (n = 65) and 10 Hz frequency for TMS (n = 57). Notably, significant improvements were most often reported following high-frequency TMS over the left DLPFC (n = 153) and anodal tDCS over the right DLPFC (n = 61).

### 3.2. Risk of Bias of the Included Studies

None of the included studies had a high risk of bias. Forty-eight percent (n=56) of the studies had some concerns. The main reason for the studies’ risk of biases being rated as some concerns was related to the “selection of the reported result domain” (n=53), followed by the “randomization process” (n=3). The thorough rating for each study can be found in Supplementary Figure S.7.

### 3.3. Meta-Analysis Phase Results

From a total of 208 studies, 1 was related to tFUS. Out of 207 tES/TMS studies in our systematic review, our meta-analysis included 50 TMS studies for craving (69 total assessments) and 29 TMS studies for consumption/relapse (64 total assessments). In tES studies, 45 studies for craving (55 total assessments) and 31 studies (total 55 assessments) for consumption/relapses were included. All other reported outcome measures are categorized in supplementary materials Table S.4. For TMS, we included 1,661 participants in the active group, and 1,610 participants in the sham group for craving, and 711 active and 695 sham participants for consumption. In total, our meta-analysis evaluated data from 5,106 participants in the active groups and 4,914 participants in the sham groups. Our results showed a significant medium effect of TMS on craving (Hedges’ g = 0.52, CI = [0.29, 0.75], p < 0.001, I² = 89.36) and a medium effect on consumption/relapse (Hedges’ g = 0.41, CI = [0.26, 0.56], p < 0.001, I² = 61.56). tES demonstrated a similar medium effect on craving (Hedges’ g = 0.40, CI = [0.25, 0.55], p < 0.001, I² = 60.69) and a small effect on consumption/relapse (Hedges’ g = 0.27, CI = [0.15, 0.38], p = 0.013, I² = 22.67). Associatedforest and funnel plots are available in the supplementary materials, Figures S8, and the specific questionnaires used in the different studies to quantify craving and consumption/relapse are detailed in Figure S.10.

### 3.4. Subgroup Analysis

In our subgroup analysis, we examined the effects of different stimulation protocols on craving and consumption/relapse outcomes. The analysis identified high-frequency deep TMS with the H4 coil as having the strongest effect on both craving (Hedges’ g = 3.92) and consumption (Hedges’ g = 1.12), with a highly significant p-value of <0.0001 (single study). The H4 coil is designed to target the anterior cingulate cortex (ACC) and the bilateral insula, which are key regions involved in the salience network (this coil induces the maximum electric field strength over the frontopolar area). This effect was followed by high-frequency repetitive TMS (rTMS) targeting the dorsolateral prefrontal cortex (DLPFC), which also showed significant (p < 0.05) reductions in craving (Hedges’ g = 0.65, in a total 21 studies) and consumption (Hedges’ g = 0.52, in a total 14 studies). Among the tES protocols, anodal tDCS over the left DLPFC with return electrode over the right DLPFC showed significant (p < 0.05) effects in reducing both craving (Hedges’ g = 0.38, 14 studies) and consumption (Hedges’ g = 0.3, 10 studies). Similarly, cathodal tDCS over the left DLPFC with the return electrode over the right DLPFC showed strong evidence (p < 0.0001) for reducing craving (Hedges’ g = 0.49, 23 studies) and consumption (Hedges’ g = 0.42, 15 studies). To show the primary target region in each protocol, we created computational head models to visualize the electric field distribution over the cortex. Based on the results, the frontopolar area in the H4 coil trials, DLPFC stimulation in tES trials, with the stimulating electrode positioned over the left or right DLPFC and the return electrode placed on the contralateral DLPFC, and the left DLPFC in rTMS trials (which demonstrated the largest Hedges’ g values) received the maximum electric field strength (Figure 5). Further subgroup analyses to show the effects of different parameters such as substance type, session type, randomization, hemisphere targeted, stimulation target, stimulation polarity or frequency, symmetry of stimulation or type of the coil, intensity, and duration of treatment can be found in supplementary materials Figure S.9. (e.g., for substance type, significant effects of tES on craving were found for opioids (Hedges’ *g* ≈ 1.26, *p* = 0.000), multiple substances (Hedges’ *g* ≈ 0.85, *p* = 0.022), nicotine (Hedges’ *g* ≈ 0.45, *p* = 0.011), and amphetamines (Hedges’ *g* ≈ 0.44, *p* = 0.000). For tES consumption, a significant effect was observed for multiple substances (Hedges’ *g* ≈ 0.98, *p* = 0.000). TMS significantly reduced craving for opioids (Hedges’ *g* ≈ 1.15, *p* = 0.006) and amphetamines (Hedges’ *g* ≈ 0.93, *p* = 0.000), and significantly reduced consumption for cocaine (Hedges’ *g* ≈ 1.25, *p* = 0.022) and nicotine (Hedges’ *g* ≈ 0.47, *p* = 0.000). All other subgroup effects were not statistically significant).

**Figure 5:**
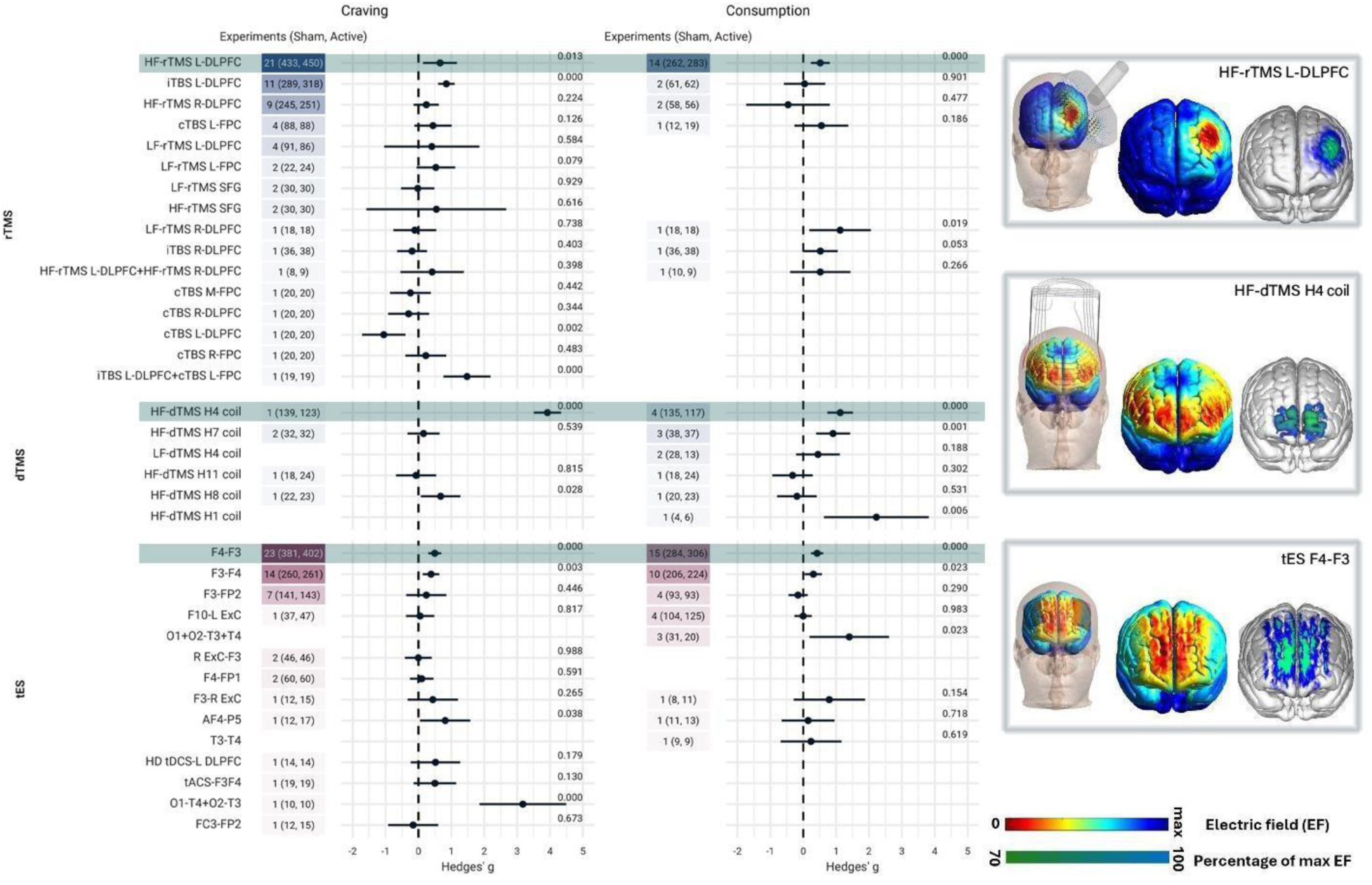
Exploring Overall Effects of 36 Protocols in tES/TMS Studies on Craving and Consumption. This figure illustrates the impact of tES/TMS on craving and consumption/relapse by examining different types of stimulation protocols. The total number of experiments is color-coded, with tES studies in red and TMS studies in blue. Additionally, the total number of participants in the active and sham arms is indicated in parentheses. Each horizontal line within the plots represents a distinct stimulation protocol, with dots for effect sizes and horizontal lines for confidence intervals. Computational head models show the protocols with the largest significant Hedges’ g values in rTMS, dTMS, and tES trials, highlighted in green. In the tES section, the first position indicates anode placement, and the second indicates cathode placement. <u>Abbreviations</u>: tES: transcranial electrical stimulation, TMS: transcranial magnetic stimulation, dTMS: deep transcranial magnetic stimulation. The figure includes three color codes: red, blue, and green. tES protocols are coded in red, while TMS protocols are coded in blue. The heatmap represents the number of studies, with darker tones indicating a higher number of assessments that utilized the corresponding protocol. On the right panel, rTMS, dTMS, and tES protocols are sorted based on the number of assessments that are used in each protocol. Protocols with significant Hedge’s g values in both craving and consumption outcomes are highlighted in green, and their computational head models are presented.

## 4. Discussion

This systematic review and meta-analysis evaluated the parameter space of NIBS techniques — TMS, tES, and tFUS — for SUDs and quantified their effects on drug craving and consumption/relapse. A total of 208 studies published through early 2024 (TMS=121, tES=86, tFUS=1) were included, revealing substantial heterogeneity in experimental design, stimulation protocols, target regions, and outcome measures. The meta-analysis incorporated 50 TMS studies (69 assessments), 45 tES studies (55 assessments) examining craving, and 29 TMS studies (64 assessments) and 31 tES studies (55 assessments) examining consumption/relapse, constituting the largest analysis to date in this field.

The results showed that TMS had significant medium effects on both craving and consumption/relapse reduction (*g* = 0.52 and *g* = 0.41, respectively), while tES had a medium effect on craving (*g* = 0.40) and a small effect on consumption/relapse (*g* = 0.27). Subgroup analyses revealed notable variability in participant characteristics, study designs, stimulation parameters, and treatment efficacy. Among 34 evaluated protocols, the most effective approaches included: (1) tES targeting the DLPFC with a bilateral montage (stimulating electrode over the left or right DLPFC and return electrode on the contralateral side with maximal electric field over the frontopolar cortex), (2) high-frequency deep TMS using an H4 coil targeting the ACC and bilateral insula (regions within the salience network, with maximal electric field over the frontopolar cortex), and (3) high-frequency rTMS over the left DLPFC. Additional subgroup findings indicated that tES was most effective in opioid, nicotine, and polysubstance use disorders, while TMS showed significant effects in nicotine, cocaine, amphetamine, and opioid use disorders. Both single- and multi-session tES trials significantly reduced craving, whereas only multi-session TMS trials were effective in reducing consumption. Other stimulation parameters did not show consistent effects across craving and consumption/relapse outcomes.

The largest prior meta-analysis by Song et al. (2019) included 1,095 participants with conditions including alcohol, nicotine, illicit drug, and food-related disorders.^17^ While their results indicated that active stimulation (tDCS and TMS) outperformed sham, the final meta-analysis results differed in some aspects from our findings. The divergence of findings could be related to differences in the included trials. Nevertheless, similar to our findings, their TMS results showed a medium effect on craving (g=0.52, CI=[0.29, 0.75], p<0.001, I²=89.36) and consumption (g=0.45, CI=[0.25, 0.65], p<0.001, I²=67.7). tES showed a medium effect on craving (g=0.40, CI=[0.25, 0.55], p<0.001) and a small effect on consumption (g=0.19, CI=[0.06, 0.32], p=0.015) (12). These results contrast with Mostafavi et al. (2020), which reported no significant effects of tDCS/rTMS in 617 participants with alcohol use disorders,^18^ but align with many other meta-analyses in the field.^6,17,19–22^

Comparing multi-session and single-session protocols, we found that increasing the number of TMS sessions was linked to larger reductions in craving and consumption, consistent with the meta-analysis by Song et al. (2019),^17^ who reported a larger impact from multi-session protocols. Within the same meta-analysis, a focused analysis of rTMS over the DLPFC also found a significant positive correlation between the total number of stimulation pulses and craving reduction effect size in studies employing excitatory stimulation over the left DLPFC (P = 0.01).^17^ However, our results for tES were more heterogeneous: while both single (n=24) and multi-session (n=31) tES protocols significantly reduced craving, only single-session (n=7), not multi-session (n=25), protocols showed a significant effect on consumption. This may be potentially due to the early assessment conducted shortly after the single stimulation, the high dropout rate in follow-ups for multi-session trials, which is a common challenge in SUD studies, or the low number of single-session studies of consumption. In this vein, it is important to note that while sufficient multi-session TMS trials (n=50 for craving and n=43 for consumption) were available, the number of trials for single-session TMS was limited (n=19 for craving and n=2 for consumption), potentially similarly affecting the robustness of these findings. Additionally, the effectiveness of rTMS in reducing craving was protocol-dependent, with variability observed across studies. The differences in outcomes may also reflect the heterogeneity in protocols, including variations in stimulation intensity, duration, frequency, and electrode/coil positioning. For example, while some protocols demonstrated strong positive effects, others yielded unclear or even negative outcomes. This variability underscores the need to interpret aggregated results cautiously, as averaging across diverse protocols may obscure the positive effects of specific approaches. Interestingly, no significant effects were observed for alcohol use disorder in the studies included in the meta-analysis, which may be due to insufficiently tailored protocols or the limited number of high-quality trials focusing on this substance.

Despite the promising results, only one protocol has received FDA clearance to date. Our meta-analysis identified this FDA-cleared protocol—dTMS with the H4 coil—as having the largest effect size compared to the other included 34 protocols (including rTMS, dTMS with different coils, and tES). However, more studies are needed with this coil with craving and consumption as outcome measures.^23^ The next most efficacious protocol, high-frequency rTMS over the left DLPFC, showed a medium but significant effect on both outcomes, supported by 21 published papers. This protocol has received CE approval in Europe for psychoactive substance use disorder (PSUD) based on the results of only two SUD trials in cocaine addiction.^24–26^ This efficacy is in line with the meta-analysis by Zhang et al. (2019),^27^ who reported that excitatory rTMS of the left DLPFC was effective in reducing substance craving and consumption (Hedge’s g = -0.62), with a significant positive association between the total number of stimulation pulses and effect size (P = 0.01). Tseng et al. (2022)^28^ also found that 10-Hz rTMS over the left DLPFC was associated with the largest decrease in smoking frequency (standardized mean difference = -1.22). Additionally, Petit et al. (2022)^29^ reported that the participants who received tES/TMS were 2.39 times more likely to achieve sustained abstinence compared to those receiving sham stimulation, with an even greater likelihood observed for rTMS over the left DLPFC (risk ratio = 4.34) and for deep rTMS targeting the lateral prefrontal cortex and insula bilaterally (risk ratio = 4.64). However, TMS applied to the right DLPFC showed mixed effects. For tES, placing the anode over either the left or right hemisphere significantly affected craving reduction. However, laterality had no significant impact on reducing consumption or relapse. The most effective tES protocols involved bilateral DLPFC stimulation with the anode placed over either the left or right DLPFC, which induced the maximum electric fields over the frontopolar area.^30^

## 5. Limitations

This meta-analysis has several limitations that should be considered when interpreting the findings. Our literature search was restricted to English-language databases, potentially missing relevant studies published in other languages. There was substantial clinical heterogeneity (e.g., participant characteristics, substances used) and methodological variability (e.g., study design and outcome assessments) among the included studies. Despite these differences, the data from these studies were aggregated and analyzed as if they represented a single, uniform sample and intervention, which may have influenced the findings. Additionally, our meta-analysis included a variety of tools for measuring craving and consumption, with 17 different questionnaires for craving, 12 for objective, and 4 for subjective measures of consumption (Figure S.10). This diversity in measurement tools may introduce inconsistencies and limit the comparability of results across studies. Differences in study designs, such as variations in session duration, frequency, and intervention intensity, may also have influenced the overall effect sizes, contributing to the observed heterogeneity. These factors emphasize the need for identifying the optimal features (for enhancing select outcomes) that could then be used to develop more standardized protocols in future research to enable more precise comparisons.

## 6. Conclusion

This systematic review and meta-analysis, the largest to date, provided a comprehensive overview of the parameter space and therapeutic effects of NIBS technologies for SUDs. Analyzing 207 TMS and tES studies across 21 countries, along with one tFUS study, the findings underscore both the promise and complexity of NIBS interventions in this field. These findings suggest that: (1) both TMS and tES show significant therapeutic effects for SUDs, with varying levels of effectiveness, (2) both conventional and deep TMS, as well as bilateral DLPFC-targeted tES, demonstrate clinical utility, with the strongest evidence observed for deep TMS (dTMS)—though this is currently based on a single published study and requires replication, (3) the frontopolar cortex—receiving the highest electric field from the FDA-approved H4 dTMS coil and bilateral DLPFC tES montages—emerges as a promising target, supported by converging neuroimaging and lesion-mapping evidence, ^31,32^ (4) while dTMS shows a larger effect size, the left DLPFC remains a validated and widely studied target. Together, these findings highlight the potential of targeting both frontopolar-limbic and DLPFC-executive control circuits as dual intervention pathways in SUD treatment.^33^ The global contribution to these studies suggests strong scalability of NIBS technologies, although cost-effectiveness and accessibility remain important areas for future investigation.

## Data Availability

All data produced in the present study are available upon reasonable request to the authors.

## Supplementary materials

### Search terms

**Table S.1.**
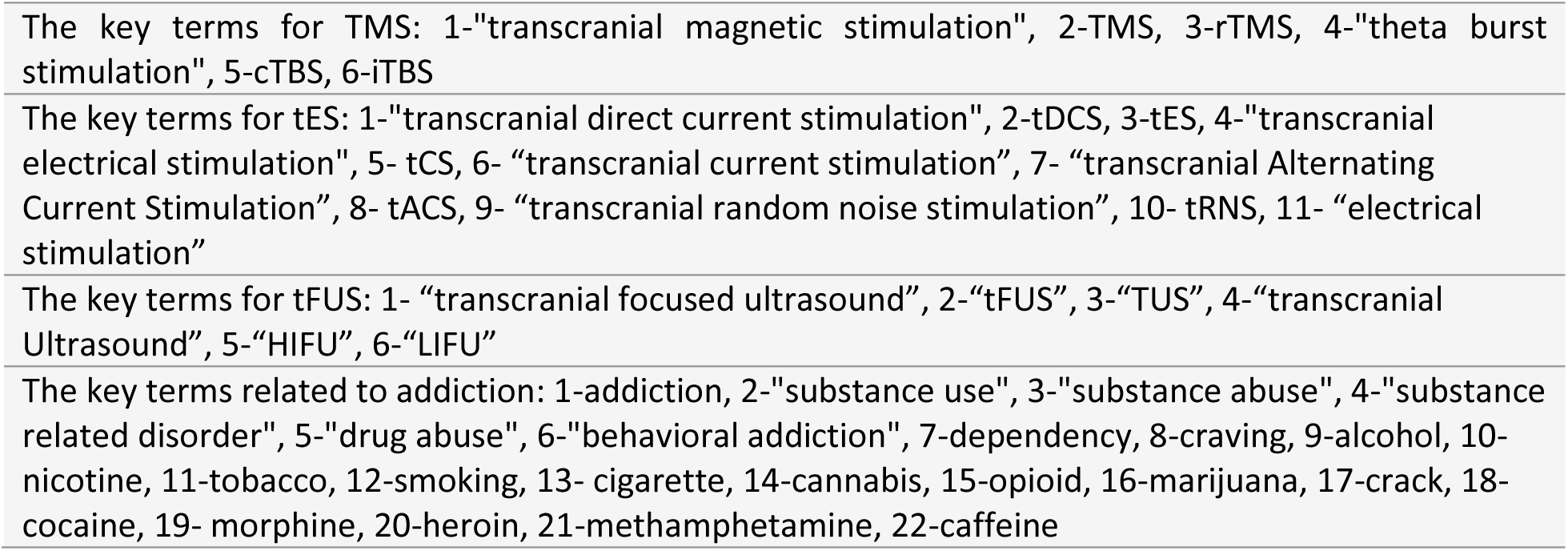
key terms used to search in the PubMed database.

|  |
| --- |
| The key terms for TMS: 1-"transcranial magnetic stimulation", 2-TMS, 3-rTMS, 4-"theta burst stimulation", 5-cTBS, 6-iTBS |
| The key terms for tES: 1-"transcranial direct current stimulation", 2-tDCS, 3-tES, 4-"transcranial electrical stimulation", 5- tCS, 6- "transcranial current stimulation", 7- "transcranial Alternating Current Stimulation", 8- tACS, 9- "transcranial random noise stimulation", 10- tRNS, 11- "electrical stimulation" |
| The key terms for tFUS: 1- "transcranial focused ultrasound", 2-"tFUS", 3-"TUS", 4-"transcranial Ultrasound", 5-"HIFU", 6-"LIFU" |
| The key terms related to addiction: 1-addiction, 2-"substance use", 3-"substance abuse", 4-"substance related disorder", 5-"drug abuse", 6-"behavioral addiction", 7-dependency, 8-craving, 9-alcohol, 10-nicotine, 11-tobacco, 12-smoking, 13- cigarette, 14-cannabis, 15-opioid, 16-marijuana, 17-crack, 18-cocaine, 19- morphine, 20-heroin, 21-methamphetamine, 22-caffeine |

### Data extraction process

After finalizing the inclusion process, literature search results were imported into three separate sheets for TMS, tES, and tFUS studies. Data extracted from the literature were filled into a spreadsheet by AS and HT. Consistency between the authors was honed through a calibration exercise in which all authors evaluated and discussed their ratings for 20 randomly chosen studies.^1^ GS further refined the data extraction form to reduce inconsistency and ambiguity after the exercise. Data on study design features and basic methodological parameters were extracted first, and each article was reviewed independently by two investigators (AS and HT) in two separate spreadsheets, with inconsistencies resolved in discussions with GS and HE. Despite following a similar data extraction policy to the previously published INTAM systematic review,^2^ a comprehensive screening of the literature by 2018 was conducted once again to ensure the absence of any inconsistencies. This step was taken to guarantee the accuracy and reliability of the extracted data. By re-screening the entire range of publications within the specified timeframe, potential discrepancies or errors in the data extraction process were identified and rectified. This approach aimed to maintain the integrity and consistency of the review, providing a robust foundation for the analysis and conclusions drawn from the included studies.

### Inclusion/Exclusion criteria and data extraction

The initial search yielded a total of 508 articles on TMS, 663 articles on tES, and 156 articles on tFUS. Two independent investigators (AS, HT) reviewed the list of included studies and collected the coded data, and the 3^rd^ investigator (GS) reviewed the collected databases and solved eventual conflicts. During the initial screening, based on titles and abstracts, 179 TMS records, 418 tES records, and 151 tFUS records were excluded. The reasons for exclusion encompassed articles that were book chapters, commentaries, author corrections, editorials, and studies focused on disorders other than substance use disorders (SUDs). One TMS study was identified as a duplicate under a different title. Subsequently, 327 TMS articles, 242 tES, and 5 tFUS articles progressed to the next stage, which involved full-text screening to identify eligible articles. A precise full-text review was conducted, resulting in the inclusion of 121 TMS, 86 tES, and 1 tFUS articles. Exclusion criteria at this stage included review articles, studies with only healthy subjects, case reports and case series, studies involving subjects other than humans, study protocols, electric field modeling, and studies published in languages other than English. Single or paired-pulse TMS studies were excluded, while studies including repetitive transcranial magnetic stimulation (rTMS) and deep transcranial magnetic stimulation (dTMS) (a type of rTMS) were included. Notably, no published study utilizing transcranial random noise stimulation (tRNS) in the field of tES was identified. The PRISMA flowchart illustrating the inclusion/exclusion procedure for TMS, tES, and tFUS studies are presented in Figure 1, respectively.

Publication details, including the country of publication (defined as the first affiliation of the first author, or the affiliation of the majority of the authors if the country of the first author was unclear), the publication year (based on PubMed’s indexing), the main substance(s) of interest in the study, the main site of stimulation (were the stimulation electrode or coil is placed), the number of active stimulation sessions (participants received active tES or TMS), the number of subjects in the active arms, and the number of female subjects in the active arm were extracted. We categorized the current state of participants to four distinct time intervals at which the interventions were administered: (1) before the participant sought standard treatment (2) while the subject was treatment seeking but before undergoing standard treatment, (3) within the first month of standard treatment (mainly detoxification and stabilization) and (4) after the initial recovery period (more than one month). Additionally, the duration of the follow-up period, the control condition (active, sham, another therapy, another clinical population, or no-control conditions), the randomization method, the coil/electrode positioning system, the stimulation dose (for TMS: site of pulses, stimulation type, frequency, intensity, and the number of pulses; for tES: anode/cathode size and location, stimulation intensity, and duration), and the primary/secondary outcome measures were extracted. Additionally, a binary rating system (Yes/No) was used to indicate whether a study observed any significant effects on a specific outcome measure in response to the stimulation. We also checked if computational head models were generated for electric field distribution pattern assessment.

**Table S.2.** Data Extraction for tES Studies.

| Study | Substance | Active arm | Current status | Session | Follow up | Control | Randomization | Positioning | Target | Anode | Cathode | Duration | Intensity | Findings |
| --- | --- | --- | --- | --- | --- | --- | --- | --- | --- | --- | --- | --- | --- | --- |
| Boggio et al.,(2008)<sup>3</sup> | Alcohol | 11 | Detoxification | 1 | NA | Active controlled and sham controlled | Crossover | EEG System | left DLPFC- Anodal tDCS/right DLPFC-Anodal tDCS | 35 | 35 | 20 | 2 | Craving ↓ |
| Fregni et al.,(2008)<sup>4</sup> | Nicotine | 23 | Active user non-treatment seeker | 1 | NA | Active controlled and sham controlled | Crossover | EEG System | left DLPFC- Anodal tDCS/right DLPFC-Anodal tDCS | 35 | 100 | 20 | 2 | Cue-induced craving ↓ |
| Boggio et al.,(2009)<sup>5</sup> | Nicotine | 13 | Active user non-treatment seeker | 5 | NA | Sham controlled | Parallel | EEG System | left DLPFC- Anodal tDCS | 35 | 100 | 20 | 2 | Craving ↓<br>Cigarette consumption ↓ |
| Boggio et al.,(2010)<sup>6</sup> | Cannabis | 17 | Detoxification | 1 | NA | Active controlled and sham controlled | Parallel | EEG System | left DLPFC- Anodal tDCS/right DLPFC-Anodal tDCS | 35 | 35 | 15 | 2 | Propensity for risk-taking ↑<br>Craving ↓ (Right anodal) |
| Nakamura-Palacios et al.,(2012)<sup>7</sup> | Alcohol | 49 | Detoxification | 1 | NA | Sham controlled | Crossover | EEG System | left DLPFC- Anodal tDCS | 35 | 35 | 10 | 1 | Mean amplitude of P3 associated with alcohol-related sounds ↑<br>Cognitive function performance ↑ |
| da Silva et al.,(2013)<sup>8</sup> | Alcohol | 6 | Detoxification | 5 | < 1 month | Sham controlled | Parallel | EEG System | left DLPFC- Anodal tDCS | 35 | 35 | 20 | 2 | Craving ↓<br>Depression symptoms ↓<br>Change in executive function ↑<br>Relapse ↓<br>Block the increase in neural activation triggered by alcohol related and neutral cues. |
| Pripfl et al.,(2013)<sup>9</sup> | Nicotine | 36 | Active user non-treatment seeker | 1 | NA | Active controlled and sham controlled | Crossover | EEG System | left DLPFC- Anodal tDCS/right DLPFC-Anodal tDCS | 5.3 | 35 | 15 | 0.45 | Risk-taking in the 'cold' decision ↓<br>Risk-taking in the 'hot' decision ↑ |
| Xu et al.,(2013)<br><sup>10</sup> | Nicotine | 24 | Pre-treatment<br>treatment<br>seeker | 1 | NA | Sham<br>controlled | Crossover | EEG<br>System | left DLPFC-Anodal tDCS | 35 | 35 | 20 | 2 | Negative affects ↓ |
| Conti et al.,(2014)<br><sup>11</sup> | Cocaine | 7 | Detoxification | 5 | 3 months | Sham<br>controlled | Parallel | EEG<br>System | right DLPFC-Anodal tDCS | 35 | 35 | 20 | 2 | Left DLPFC activity ↑ (Neutral<br>cues)<br>Left DLPFC activity ↓ (Drug<br>cues)<br>Right DLPFC, FPC, OFC, ACC<br>activity ↑ (Drug cues)<br>Relapse ↓ |
| Conti & Nakamura-Palacios,(2014)<br><sup>12</sup> | Cocaine | 7 | Detoxification | 1 | NA | Sham<br>controlled | Parallel | EEG<br>System | right DLPFC-Anodal tDCS | 35 | 35 | 20 | 2 | ACC activity after visualization<br>of drug cues ↓ |
| Klauss et al.,(2014)<br><sup>13</sup> | Alcohol | 16 | Detoxification | 10 | 6 months | Sham<br>controlled | Parallel | EEG<br>System | right DLPFC-Anodal tDCS | 35 | 35 | 13 | 2 | Relapse ↓<br>Perception of quality of life ↑ |
| Gorini et al.,(2014)<br><sup>14</sup> | Cocaine | 18 | Detoxification | 1 | NA | Active<br>controlled<br>and sham<br>controlled | Crossover | EEG<br>System | left DLPFC-Anodal<br>tDCS/right<br>DLPFC-Anodal<br>tDCS | 32 | 32 | 20 | 1.5 | Risk taking (from BART) ↓<br>Safe behaviors (from GDT) ↑<br>(right DLPFC)<br>Risk taking (from GDT) ↑ (left<br>DLPFC) |
| Fecteau et al.,(2014)<br><sup>15</sup> | Nicotine | 12 | Pre-treatment<br>treatment<br>seeker | 5 | NA | Sham<br>controlled | Crossover | EEG<br>System | right DLPFC-Anodal tDCS | 35 | 35 | 30 | 2 | Cigarette consumption ↓<br>Rejected offers of cigarettes<br>↑ |
| Meng et al.,(2014)<br><sup>16</sup> | Nicotine | 30 | Active user non-treatment<br>seeker | 1 | NA | Active<br>controlled<br>and sham<br>controlled | Parallel | EEG<br>System | right TPC-Cathodal<br>tDCS/left TPC-Cathodal<br>tDCS/right<br>TPC-Cathodal<br>tDCS | 33.<br>18 | 33.<br>18 | 20 | 1 | Bilateral cathodal stimulation<br>resulted in a decreasing trend<br>in attention to smoking-related<br>cues; however, the effects<br>were not significantly different<br>from sham stimulation.<br>Cigarette consumption ↓ |
| Shahbabaie et al.,(2014)<br><sup>17</sup> | Amphetamin<br>es | 30 | Detoxification | 1 | NA | Sham<br>controlled | Crossover | EEG<br>System | right DLPFC-Anodal tDCS | 35 | 35 | 20 | 2 | Immediate craving ↓<br>Cue-induced craving ↑ |
| Batista et al.,(2015)<br><sup>18</sup> | Cocaine | 17 | Detoxification | 5 | < 1<br>month | Sham<br>controlled | Parallel | EEG<br>System | right DLPFC-Anodal tDCS | 35 | 35 | 20 | 2 | Craving ↓ |
| den Uyl et al.,(2015)<br><sup>19</sup> | Alcohol | 29 | Active user non-treatment seeker | 1 | NA | Active controlled and sham controlled | Parallel | NA | left DLPFC-Anodal tDCS/right IFG-Anodal tDCS | 35 | 35 | 10 | 1 | Mild craving ↓ (in the DLPFC group)<br>Reaction time ↓ |
| Prippl & Lamm,(2015) | Nicotine | 17 | Active user non-treatment seeker | 1 | NA | Active controlled and sham controlled | Crossover | EEG System | left DLPFC-Anodal tDCS/right DLPFC-Anodal tDCS | 5.3 | 35 | 15 | 0.45 | Negative affect in emotion appraisal ↓ (In anodal right group) |
| Smith et al.,(2015)<br><sup>21</sup> | Nicotine | 14 | Active user non-treatment seeker | 5 | < 1 month | Sham controlled | Parallel | EEG System | left DLPFC-Anodal tDCS | 5.0<br>8 | 5.0<br>8 | 20 | 2 | MATRICES Consensus Cognitive Battery (MCCB) Composite score ↑<br>MCCB Working Memory and Attention-Vigilance ↑ |
| den Uyl et al.,(2017)<br><sup>22</sup> | Alcohol | 30 | Detoxification | 4 | 12 months | Active controlled and sham controlled and another therapy | Parallel | NA | left DLPFC-Anodal tDCS | 35 | 100 | 20 | 2 | Craving ↓<br>Approach bias ↓<br>Relapse ↓ |
| den Uyl et al.,(2016)<br><sup>23</sup> | Alcohol | 19 | Active user non-treatment seeker | 3 | NA | Active controlled and sham controlled and another therapy | Parallel | EEG System | left DLPFC-Anodal tDCS | 35 | 35 | 15 | 1 | Cue-induced craving ↓ |
| Falcone et al.,(2016)<br><sup>24</sup> | Nicotine | 25 | Active user non-treatment seeker | 1 | NA | Sham controlled | Crossover | EEG System | left DLPFC-Anodal tDCS | 25 | 25 | 20 | 1 | Latency to smoke ↑<br>Cigarette consumption ↓ |
| Nakamura-Palacios et al.,(2016) | Alcohol/Cocaine | 21 | Detoxification | 5 | NA | Sham controlled | Parallel | EEG System | right DLPFC-Anodal tDCS | 35 | 35 | 20/<br>26 | 2 | Relapse ↓<br>Craving ↓<br>vmPFC activation ↑ |
| Wang et al.,(2016)<br><sup>26</sup> | Opioids | 10 | Recovery | 1 | NA | Sham controlled | Parallel | NA | left TPC-Cathodal tDCS | 35 | 35 | 20 | 1.5 | Craving ↓ |
| Wietschorke et al.,(2016)<br><sup>27</sup> | Alcohol | 15 | Detoxification | 1 | NA | Sham controlled | Parallel | EEG System | right DLPFC-Anodal tDCS | 35 | 35 | 20 | 1 | Intention and desire to drink ↓<br>Startle amplitudes ↑ for alcohol-related cues (indicating more negative processing in alcohol-dependent patients) |
| Krocze et al.,(2016)<br>28 | Nicotine | 13 | Active user non-treatment seeker | 1 | NA | Sham controlled | Parallel | NA | left DLPFC-Anodal tDCS | 35 | 35 | 15 | 2 | Functional connectivity between the dlPFC and the OFC ↑ |
| Shahbabale, Ebrahimpour, et al.,(2018)<br>29 | Amphetamines | 15 | Detoxification | 1 | NA | Sham controlled | Crossover | EEG System | right DLPFC-Anodal tDCS | 35 | 35 | 20 | 2 | Craving ↓<br>Significant modulation of DMN, ECN, and SN networks |
| Yang et al.,(2017)<br>30 | Nicotine | 32 | Active user non-treatment seeker | 1 | NA | Sham controlled | Crossover | EEG System | left DLPFC-Anodal tDCS | 35 | 100 | 30 | 1 | Craving ↓ |
| Klauss, Anders, Felipe, Nitsche, et al.,(2018)<br>31 | Alcohol | 23 | Recovery | 10 | 3 months | Sham controlled | Parallel | EEG System | right DLPFC-Anodal tDCS | 35 | 35 | 20 | 2 | Craving ↓<br>Relapse ↓ |
| Vitor de Souza Brangioni et al.,(2018)<br>32 | Nicotine | 19 | Active user non-treatment seeker | 5 | < 1 month | Sham controlled | Parallel | EEG System | left DLPFC-Anodal tDCS | 35 | 35 | 20 | 1 | Cigarettes consumption ↓<br>Higher levels of motivation were associated with a larger tDCS response. |
| Mondino et al.,(2018)<br>33 | Nicotine | 17 | Pre-treatment treatment seeker | 10 | < 1 month | Sham controlled | Parallel | EEG System | right DLPFC-Anodal tDCS | 35 | 100 | 20 | 2 | Craving ↓<br>Brain activity (right posterior cingulate) ↓ |
| den Uyl et al.,(2018)<br>34 | Alcohol | 20 | Active user non-treatment seeker | 4 | 12 months | Sham controlled and another therapy | Parallel | EEG System | left DLPFC-Anodal tDCS | 35 | 100 | 20 | 2 | Only active tDCS combined with real attentional bias modification (ABM) showed an overall avoidance bias toward alcohol-related cues. No main or interaction effects of tDCS and ABM on the bias scores, craving, or relapse(effects on relapse after active tDCS were in the expected direction). |
| Sharifi-Fardshad et al.,(2018)<br>35 | Opioids | 20 | Recovery | 1 | NA | Sham controlled | Parallel/Crossover | EEG System | right DLPFC-Anodal tDCS/left DLPFC-Anodal tDCS | 35 | 35 | 20 | 2 | Drug craving ↓ |
| Klauss, Anders, Felipe, Ferreira, et al.,(2018) <sup>36</sup> | Cocaine | 19 | Detoxification | 10 | 2 months | Sham controlled | Parallel | EEG System | right DLPFC-Anodal tDCS | 35 | 35 | 20 | 2 | Craving scores ↓ (over five measurements, in both sham- and real tDCS groups)<br>Relapse rates were high and quite similar between groups in the 30- and 60-days follow-up after discharge from the hospital. |
| Shahbabaie, Hatami, et al.,(2018) <sup>37</sup> | Amphetamines | 75 | Detoxification | 1 | NA | Active controlled and sham controlled | Parallel | EEG System | right DLPFC-Anodal tDCS/right DLPFC-Anodal tDCS/left DLPFC-Anodal tDCS/left DLPFC-Anodal tDCS/left DLPFC-Anodal tDCS | 35 | 35 | 26 | 2 | Attentional bias ↓ (left DLPFC/right shoulder and left DLPFC/right DLPFC montages) |
| Eskandari et al.,(2019) <sup>38</sup> | Opioids | 20 | Detoxification | 1 | NA | Active controlled and sham controlled | Parallel |  | left DLPFC-Anodal tDCS/right DLPFC-Anodal tDCS | NA | NA | 20 |  | No significant difference between the two active groups<br>Right DLPFC: BDNF ↑ anxiety ↓ craving ↓<br>Left DLPFC: depression ↓ anxiety ↓ stress ↓ craving ↓ |
| Hajloo et al.,(2019) <sup>39</sup> | Nicotine | 20 | Active user non-treatment seeker | 10 | 1 month | Sham controlled | Parallel | EEG System | left DLPFC-Anodal tDCS | NA | NA | 20 | 2 | Number of cigarette smoking ↓ (both daily and social smoking groups)<br>Craving ↓ |
| Martinotti et al.,(2019) <sup>40</sup> | Alcohol/Cannabis/Cocaine/Opioids | 18 | Pre-treatment treatment seeker | 5 | NA | Sham controlled | Parallel | EEG System | left DLPFC-Cathodal tDCS | 25 | 25 | 20 | 1.5 | Hamilton Depression Rating Scale, Hamilton Anxiety Rating Scale, Barratt Impulsiveness Scale, and visual analog scale (VAS) in all subjects ↓<br>Significant time by group interaction for VAS in favor of the active group.<br>Regression analysis showed that the pre-VAS score had a significant influence on the post-VAS score. |
| Tarehian et al.,(2019) <sup>41</sup> | Opioids | 20 | Recovery | 10 | < 1 month | Sham controlled and another therapy | Parallel | EEG System | right DLPFC-Anodal tDCS | 35 | 35 | 20 | 2 | Craving ↓ (in the active tDCS group compared to sham and methadone-only groups)<br>Depression and anxiety symptoms ↓ (in the active tDCS group compared to the other two groups) |
| Claus et al.,(2019) <sup>42</sup> | Alcohol | 20 | Active user non-treatment seeker | 4 | 1 month | Sham controlled and another therapy | Parallel | EEG System | right IFG-Anodal tDCS | 11 | 11 | 20 | 2 | Alcohol approach biases ↔ remained significant at baseline, but neither cognitive bias modification, tDCS, nor the interaction reduced bias at follow-up.<br>Drinks per drinking day (DDD) ↓ (1-week/1-month follow-ups)<br>DDD and percent heavy drinking days ↔ no significant effects of the intervention |
| Ghorbani Behnam et al.,(2019) <sup>43</sup> | Nicotine | 70 | Pre-treatment treatment seeker | 20 | 6 months | Active controlled and sham controlled and another therapy | Parallel | EEG System | left DLPFC-Anodal tDCS | 35 | 100 | 20 | 2 | Abstinence rates ↔ 6-month point: Group D (25.7%), Group A (20%), Group B (7%), Sham groups C and D (3.1% and 3%)<br>Nicotine dependency ↓ (Group D compared to Group A)<br>Abstinence rates ↔ no significant differences between Group D and Group A (salivary cotinine) |
| Alghamdi et al., (2019)<br>44 | Nicotine | 12 | Pre-treatment treatment seeker | 3 | 4 months | Sham controlled | Parallel | EEG System | right DLPFC-Cathodal tDCS | 25 | 25 | 20 | 1.5 | Significant effect of time but no effect of group for the number of smoked cigarettes. Number of cigarettes in both groups ↓<br>No significant changes in long-term follow-up. |
| Falcone et al., (2019)<br>45 | Nicotine | 71 | Pre-treatment treatment seeker | 3 | < 1 month | Active controlled and sham controlled | Parallel | EEG System | left DLPFC-Anodal tDCS | 25 | 25 | 20 | 1/2 | Abstinence rates ↔ no significant differences across dose groups during the quit period (sham: 2.5, 1 mA: 2.5, 2 mA: 2.4 days)<br>Smoking rate ↔ no significant changes among dose groups (sham: 12.6, 1 mA: -11.8, 2 mA: -11.7 CPD)<br>Latency to smoke and cigarettes smoked ↔ no effects of dose group during the smoking lapse paradigm<br>Side effects ↔ mild (<5 out of 10); active and sham treatments indistinguishable by participants |
| Witkiewitz et al., (2019)<br>46 | Alcohol | 47 | Pre-treatment treatment seeker | 8 | 2 months | Sham controlled and another therapy | Parallel | EEG System | right IFG-Anodal tDCS | 15 | 15 | 30 | 2 | Drinks per drinking day ↓<br>Number of groups attended ↔ significant dose effect<br>Percent heavy drinking days and alcohol cue reactivity ↔ significant effects of time and dose for groups attended<br>Primary and secondary outcomes ↔ no significant differences between active and sham tDCS |
| Verveer, van der Veen, et al., (2020)<br>47 | Cocaine | 29 | Detoxification | 10 | 3 months | Sham controlled | Parallel | EEG System | right DLPFC-Anodal tDCS | 35 | 35 | 13 | 2 | Cocaine use days and craving ↔ no significant effects of active tDCS<br>Relapse rates ↔ frequent in both active (48%) and sham (69.2%) tDCS groups, despite ↓ craving during the first two weeks<br>Cognitive functions ↔ no observed effects<br>Crack cocaine relapse rates ↓ (active tDCS group) |
| Verveer, Remmerswaal, van der Veen, et al., (2020)<br>48 | Nicotine | 34 | Active user non-treatment seeker | 6 | 3 months | Sham controlled | Parallel | EEG System | right DLPFC-Anodal tDCS | 35 | 35 | 13 | 2 | No significant effect of active tDCS one day after stimulation protocols in terms of behavioral and neurophysiological.<br>A three-month follow-up revealed significant improvement in reaction time and a decrease in P3 amplitude in response to smoking cues. |
| Holla et al., (2020)<br>49 | Alcohol | 11 | Detoxification | 5 | 3 months | Sham controlled | Parallel | EEG System | left DLPFC-Cathodal tDCS | 35 | 35 | 20 | 2 | Integrated global efficiency of brain networks ↑ (predictor of relapse)<br>Integrated global clustering ↓ (positive correlation with impulsivity)<br>Functional connectivity of a specific sub-network involving prefrontal regions ↑ |
| Kooteh et al., (2020)<br>50 | Opioids | 14 | Recovery | 8 | NA | Active controlled and another therapy | Parallel | EEG System | right DLPFC-Anodal tDCS | 35 | 35 | 45 | 2 | Significant differences between groups in terms of craving and thoughts and fantasies about drug use. The decrease is higher in the tDCS + emotion regulation group. |
| Avonson Fischell et al.,(2020)<sup>51</sup> | Nicotine | 43 | Active user non-treatment seeker | 1 | NA | Active controlled and sham controlled | Crossover | Beam F3 | left DLPFC-Anodal tDCS/right FPC-Anodal tDCS | 35 | 35 | 25 | 2 | Default mode network (DMN) nodes during the working memory task ↓<br>Activation of ACC (greater activity in smokers) ↑<br>Nicotine withdrawal reduced task engagement and attention and reduced suppression of DMN nodes. |
| Verveer, Remmerswaal, Jongerling, et al.,(2020)<sup>52</sup> | Nicotine | 35 | Active user non-treatment seeker | 6 | 3 months | Sham controlled | Parallel | EEG System | right DLPFC-Anodal tDCS | 35 | 35 | 13 | 2 | Number of smoked cigarettes a day up to one week after the last tDCS session in both conditions ↓<br>Active treatment had no additional effect on cigarette consumption, craving, and affect. |
| Sadeghi Bimorgh et al.,(2020)<sup>53</sup> | Opioids | 14 | Detoxification | 7 | NA | Sham controlled and another therapy | Parallel | EEG System | right DLPFC-Anodal tDCS | 35 | 35 | 20 | 2 | Depression, anxiety, and stress of participants ↓ significantly in the intervention group.<br>The relapse rate showed no significant changes between the two groups. |
| Vanderhasselt et al.,(2020)<sup>54</sup> | Alcohol | 45 | Active user non-treatment seeker | 1 | NA | Sham controlled | Crossover | EEG System | right DLPFC-Anodal tDCS | 35 | 35 | 20 | 2 | Reward-triggered approach bias ↓<br>Alcohol consumption ↓<br>No significant change in self-reported craving<br>Association between, reward-triggered approach bias and alcohol consumption only in the sham group. |
| Mondino et al.,(2020)<sup>55</sup> | Nicotine | 19 | Pre-treatment treatment seeker | 1 | NA | Sham controlled and another therapy | Crossover | EEG System | left DLPFC-tACS/right DLPFC-tACS | 9 | 9 | 30 | 2 | Time spent looking at smoking-related pictures (tACS+ABM) ↓<br>Prevented the increase of self-reported desire to smoke.<br>Proportion of impulsive choices ↓ |
| Daughters et al.,(2020)<sup>56</sup> | Alcohol/Cocaine/Cannabis/Opioids | 20 | Pre-treatment treatment seeker | 1 | NA | Active controlled and sham controlled | Parallel | EEG System | left DLPFC-tACS/right DLPFC-tACS | 25 | 35 | 40 | 2 | Inhibitory control ↑<br>A large and significant effect of alpha-Tacs.<br>There were no significant effects of the condition on distress tolerance. |
| Alzadehgoradel et al.,(2020)<sup>57</sup> | Amphetamines | 19 | Detoxification | 10 | 1 month | Sham controlled | Parallel | EEG System | left DLPFC-Anodal tDCS | 35 | 35 | 20 | 2 | Task performance across all executive functions (EFs) (immediately after and 1 month) ↑<br>Craving (immediately after and 1 month) ↓<br>A significant correlation between cognitive control improvement and craving reduction. |
| Dormal et al.,(2020)<sup>58</sup> | Alcohol | 20 | Active user non-treatment seeker | 1 | NA | Sham controlled | Crossover | EEG System | left DLPFC-Anodal tDCS | 35 | 35 | 20 | 1.5 | No significant behavioral differences between tDCS and sham stimulation groups.<br>Attentional resource mobilization (indexed by the N2 component) ↑<br>Inhibition processes (indexed by the P3 component) remained unchanged. |
| Brown et al., (2020)<br>59 | Alcohol | 36 | Pre-treatment treatment seeker | 8 | NA | Sham controlled and another therapy | Parallel | EEG System | right IFG-Anodal tDCS | 15 | 15 | 30 | 2 | In response to alcohol cues:<br>Craving ratings ↓<br>Late positive potential (LPP) ↓<br>The magnitude of alcohol image craving reductions was associated with the number of mindfulness-based relapse prevention (MBRP) group sessions attended.<br>Active tDCS was not associated with craving ratings, but it was associated with greater LPP amplitudes across image types. |
| Mostafaei et al., (2022)<br>60 | Opioids | 20 | Detoxification | 10 | NA | Active controlled and sham controlled | Parallel | EEG System | left DLPFC-Anodal tDCS/right DLPFC-Anodal tDCS | NA | NA | 20 | 2 | The amplitude of slow brain waves (delta, theta, and alpha) in prefrontal, frontal, occipital, and parietal areas ↓<br>Coherence of beta, delta, and theta frequency bands in the parietal, central, and temporal regions ↑<br>Sham group: The amplitude of the alpha wave ↓<br>The coherence of delta and theta waves ↓ |
| Perri & Perrotta, (2021)<br>61 | Nicotine | 10 | Active user non-treatment seeker | 5 | NA | Sham controlled | Parallel | EEG System | right DLPFC-Anodal tDCS | 25 | 25 | 20 | 2 | Cigarette craving (50%) ↓<br>The number of cigarettes smoked remained unchanged. |
| Gaudreault et al., (2021)<br>62 | Cocaine | 8 | Recovery | 15 | 1 month | Sham controlled | Parallel | EEG System | right DLPFC-Anodal tDCS | 35 | 35 | 20 | 2 | Self-reported craving ↓<br>Perceived quality of life ↑<br>Impulsivity ↓<br>Daytime sleepiness and readiness to change drug use showed significant Group × Time interactions, with improvements observed only in the real-tDCS group.<br>One-month follow-up indicated transient effects of tDCS on sleepiness and craving. |
| Xu et al., (2021)<br>63 | Amphetamines | 24 | Recovery | 20 | NA | Sham controlled and another therapy and control group | Parallel | EEG System | right DLPFC-Anodal tDCS | 38.5 | 38.5 | 20 | 1.5 | Craving ↓ by tDCS + computerized cognitive addiction therapy (CCAT)<br>Craving in active tDCS + CCAT < sham tDCS + CCAT<br>Two Back task (TWOB) improved by tDCS + CCAT.<br>Discounting did not change with active or sham tDCS. |
| Eskandari et al., (2021)<br>64 | Opioids | 21 | Detoxification | 10 | NA | Active controlled and sham controlled | Parallel | EEG System | left DLPFC-Anodal tDCS/right DLPFC-Anodal tDCS | NA | NA | 20 | 2 | Drug craving (active and sham) ↓<br>Impulsivity and its dimensions (right/left DLPFC) ↓<br>Levels of IL-6 and TNF-α (not statistically) ↓ |
| Alizadehgoradei et al., (2021)<br>65 | Amphetamines | 16 | Detoxification | 12 | 1 month | Active controlled and sham controlled and another therapy | Parallel | EEG System | left DLPFC-Anodal tDCS | NA | NA | 20 | 1.5 | Performance in most executive functions (EFs) tasks (tDCS+) ↑ (post-test phase and follow-up phase in real tDCS + mindfulness-based substance abuse treatment (MBSAT))<br>Craving ↓ (after intervention in all treatment groups, but not the sham stimulation group)<br>The increase in EFs and the reduction in craving post versus pre-tDCS + MBSAT intervention were correlated. |
| Dubuson et al., (2021)<br>66 | Alcohol | 64 | Pre-treatment treatment seeker | 5 | 12 months | Active controlled and sham controlled and another therapy | Parallel | EEG System | right DLPFC-Anodal tDCS | 35 | 35 | 20 | 2 | Abstinence rate at the 2-week follow-up ↑ (did not persist beyond two weeks after the intervention)<br>Higher abstinence rates for the verum tDCS associated with alcohol cue inhibitory control training (ICT).<br>Neither the reduction of craving nor the improvement in executive control resulted specifically from prefrontal tDCS and ICT. |
| McKim et al., (2021)<br>67 | Alcohol | 17 | Recovery | 1 | NA | Sham controlled and healthy control | Parallel/Crossover | EEG System | left DLPFC-tACS/right DLPFC-tACS | 19.80<br>25 | 42.408<br>5 | 30 | 2 | Perseverative errors ↓ (in SUD group following tACS; reduction dependent on addiction history duration)<br>Perseverative errors ↑ (in the control group following 10 Hz-tACS)<br>Behavioral flexibility ↔ altered circuit-level dynamics by tACS<br>Chronic substance abuse ↔ associated with circuit dynamics ameliorated by tACS |
| Müller et al., (2021)<br>68 | Nicotine | 24 | Active user non-treatment seeker | 5 | NA | Sham controlled | Parallel | EEG System | left DLPFC-Anodal tDCS | 25 | 25 | 20 | 2 | Cigarette consumption ↓ (significant decrease observed after stimulation in both intervention groups)<br>CO levels ↓ (significant decrease observed after stimulation in both intervention groups)<br>Craving ↓ (significant decrease observed after stimulation in both intervention groups)<br>Executive functions ↔ no improvements in either group<br>Active group ↔ higher perceived stress linked to lower cigarette consumption; lower self-control linked to higher CO reduction |
| Lin et al., (2021)<br>69 | Nicotine/Opioids | 18 | Active user non-treatment seeker | 5 | < 1 month | Sham controlled | Parallel | EEG System | left DLPFC-Anodal tDCS/left FPC-Anodal tDCS | NA | NA | 20 | 2 | The expired CO concentration ↓<br>Number of cigarettes smoked in both groups ↓<br>No significant group difference with regard to craving tendency. |
| Patel et al., (2022)<br>70 | Cannabis | 15 | Active user non-treatment seeker | 1 | NA | Sham controlled | Parallel | Beam F3 | right DLPFC-Anodal tDCS | 35 | 35 | 15 | 2 | No significant effects of stimulation on risk-taking behavior were found.<br>There was a significant ↑ delay in discounting of a \$1000 delayed reward during stimulation for the sham group.<br>No significant effects for probability discounting. |
| Jiang et al., (2022)<br>71 | Amphetamines | 23 | Recovery | 5 | NA | Sham controlled | Parallel | MRI guided | right DLPFC-Anodal tDCS | 11.4 | 11.4 | 20 | 2 | Behavioral impulsivity ↑ (decreased accuracy for the deviant stimulus) |
| Wedler et al., (2022)<br>72 | Alcohol/Nicotine | 35 | Active user non-treatment seeker | 1 | NA | Sham controlled and healthy control | Parallel | EEG System | right DLPFC-Anodal tDCS | 35 | 100 | 20 | 1.5 | Alcohol-dependent patients (AD) did not exhibit the typical increase in aggression over time observed in other groups.<br>Response inhibition ↑ in AD and tobacco users (TU) (following active, but not sham, stimulation) |
| Fayaz Feyzi et al.,(2022)<br>73 | Amphetamines | 15 | Active user non-treatment seeker | 16 | 1 month | Sham controlled and another therapy | Parallel | EEG System | left DLPFC-Anodal tDCS | 35 | 35 | 20 | 2 | Craving ↓ (in the active tDCS group compared to sham and psychotherapy-only groups)<br>Auditory and visual memory ↑ (Wechsler), true and false answers ↑ (WCST) in the active tDCS<br>Relapse rates ↔ no significant differences between groups<br>A significant correlation in craving reduction and cognitive functioning (active tDCS group) |
| Gibson et al.,(2022)<br>74 | Alcohol | 47 | Pre-treatment treatment seeker | 8 | 2 months | Sham controlled | Parallel | EEG System | right IFG-Anodal tDCS | 15 | 15 | 30 | 2 | Treatment adherence by the tDCS condition did not predict mindfulness; however, their interaction significantly predicted post-treatment craving.<br>In the sham group, higher treatment adherence was associated with lower post-treatment craving, while no such relationship was observed in the active tDCS group. |
| Meng et al.,(2022)<br>75 | Opioids/Nicotine | 11 | Recovery | 1 | NA | Sham controlled | Parallel | EEG System | left TPC-Cathodal tDCS/right TPC-Cathodal tDCS | 33.16<br>625 | 33.166<br>25 | 20 | 1 | Daily cigarette consumption ↓ (observed in heavy smokers and those with heroin use history, lasting at least 48 hours)<br>Detectable changes of the dynamic pupil light reflex after bilateral tDCS stimulation. |
| Ekhtari et al.,(2022)<br>76 | Amphetamines | 30 | Detoxification | 1 | NA | Sham controlled | Parallel | Beam F3 | right DLPFC-Anodal tDCS | 35 | 35 | 20 | 2 | Self-reported craving ↓<br>↓ Brain reactivity to cues following sham but not active tDCS.<br>There were significant time-by-group interactions in five main clusters in the middle and inferior frontal gyri, anterior insula, inferior parietal lobule, and precuneus. There was a significant effect of stimulation type in the relationship between electrical current at the individual level and changes in task-modulated activation. Brain regions with the highest electric current in the prefrontal cortex showed a significant time-by-group interaction in task-modulated connectivity in the frontoparietal network.<br>In this trial, there was no significant effect of the one session of active-F4/Fp1 tDCS on drug craving self-report compared to sham stimulation. However, activation and connectivity differences induced by active compared to sham stimulation suggested some potential mechanisms of tDCS to modulate neural response to drug cues. |
| Khajepour et al.,(2022)<br>77 | Amphetamines | 22 | Detoxification | 1 | NA | Sham controlled | Parallel | EEG System | right DLPFC-Anodal tDCS | 3.14 | 3.14 | 20 | 2 | P3 amplitude ↓ (following tDCS session compared to the sham group) P3 amplitude changes ↔ significantly correlated with communication efficiency (brain connectivity network (BCN) topology metrics)<br>LPP amplitude ↔ no significant change due to tDCS application |
| Soleimani et al., (2022)<br>78 | Amphetamines | 15 | Detoxification | 1 | NA | Sham controlled | Crossover | EEG System | right DLPFC-Anodal tDCS | 35 | 35 | 20 | 2 | Craving ↓<br>PFC node in executive control (ECN) ↑ PPI connectivity with precuneus and visual cortex<br>PFC node in default mode (DMN) ↓ PPI connectivity with contralateral parietal cortex<br>Connectivity ↑ within/between ECN-ventral attention (VAN)<br>Connectivity ↓ between ECN-DMN<br>Functional activity ↑ in ECN/VAN PFC nodes<br>Functional activity ↓ in DMN PFC node<br>EF direction ↔ outward in DMN; inward in ECN/VAN<br>Bilateral tDCS ↔ cortical excitability ↑ in ECN/VAN; opposite effects in DMN. |
| Schwippel et al., (2022) | Alcohol | 27 | Detoxification | 1 | NA | Sham controlled | Crossover | EEG System | left DLPFC-Cathodal tDCS | 35 | 35 | 30 | 1 | Implicit bias ↔ alcohol avoidance and nondrinking identity observed in abstinent alcohol use disorder patients<br>Cathodal tDCS ↔ no modulation of alcohol-related implicit associations or control task performance |
| Kumar et al., (2022)<br>80 | Opioids | 14 | Detoxification | 10 | NA | Sham controlled | Parallel | EEG System | left DLPFC-Anodal tDCS | 3.14 | 3.14 | 20 | 2 | Drug Questionnaire (DDQ) ↓<br>Obsessive-Compulsive Drug Use Scale (OCDUS) ↓ (except for 2 subcomponents)<br>Clinical Opiate Withdrawal Scale (COWS) ↓<br>Glutamate-glutamine and GABA ↔ no significant changes at DLPFC in either active or sham groups<br>Comparable changes observed in DDQ, OCDUS (except 2 subcomponents), COWS, and neurochemicals between active and sham groups. |
| Rezvanian et al., (2022)<br>81 | Amphetamines | 15 | Pre-treatment treatment seeker | 1 | NA | Active controlled | Crossover | EEG System | right DLPFC-Anodal tDCS/left DLPFC-Anodal tDCS/right cerebellar-Anodal tDCS/left cerebellar-Anodal tDCS | NA | NA | 20 | 2 | Craving ↔ no significant changes observed after a single session of tDCS<br>Cognitive inhibition ↑ (notably in protocol 2: right DLPFC cathodal and left DLPFC anodal stimulation) |
| Camchong et al., (2023)<br>82 | Alcohol | 31 | Detoxification | 10 | 4 months | Sham controlled and another therapy | Parallel | EEG System | left DLPFC-Anodal tDCS | 25 | 25 | 13 | 2 | Causal discovery analysis (CDA)-determined connectivity ↑ (LDLPFC to incentive salience and negative emotionality addiction networks in the active tDCS group only)<br>Relapse rates ↓ (lower probability of relapse in active tDCS group compared to sham)<br>Relapse rates ↔ unexpected sex-dependent effects observed in active tDCS group |
| Chen et al., (2023)<br>83 | Amphetamines | 10 | Recovery | 10 | NA | Sham controlled and control group | Parallel | EEG System | right DLPFC-Anodal tDCS | 35 | 35 | 13 | 2 | Behavioral performance ↑ and cost of invalid cues ↓ (in drug users following tDCS delivery compared to controls)<br>P300 amplitude ↑ (specifically for neutral cues after tDCS, resembling patterns seen in non-drug users) |
| Qin et al.,(2023)<br>84 | Nicotine | 42 | Active user non-treatment seeker | 2/5 | < 1 month | Active controlled and sham controlled and another therapy | Parallel/Crossover | EEG System | right DLPFC-Anodal tDCS | 35 | 35 | 25/20 | 2 | Cigarette consumption ↓ (enhancement effect of tDCS on psychoeducation (PE)) Craving ↓ and cigarette consumption ↓ (cumulative effect of repeated PE during intervention)<br>Cigarette intake ↔ maintained at a low level for one week with PE combined with tDCS compared to PE alone. |
| He et al.,(2023)<br>85 | Amphetamines | 58 | Recovery | 10 | NA | Active controlled and sham controlled and healthy control | Parallel | EEG System | left DLPFC-Anodal tDCS/left DLPFC-Cathodal tDCS | NA | NA | 20 | 2 | Aggressiveness ↑ (in Methamphetamine (MA) addicts compared to healthy controls)<br>Aggressiveness ↓ (following anodal tDCS intervention on the left DLPFC, especially under high-intensity provocation)<br>Time × tDCS group ↔ statistically significant interaction |
| Lu et al.,(2023)<br>86 | Nicotine | 33 | Active user non-treatment seeker | 1 | NA | Active controlled and sham controlled | Parallel | EEG System | left DLPFC-Anodal tDCS/right DLPFC-Anodal tDCS | 35 | 35 | 20 | 2 | Ipsilateral precuneus activity ↓ (following anodal stimulation over one DLPFC area during the go task state)<br>Right DLPFC stimulation ↑ functional connectivity between bilateral DLPFCs and right anterior cingulate cortex (associated with cognition and executive function inhibition) |
| Soleimani et al.,(2023)<br>87 | Amphetamines | 30 | Detoxification | 1 | NA | Sham controlled | Parallel | Beam F3 | right DLPFC-Anodal tDCS | 35 | 35 | 20 | 2 | At voxel, regional, and cluster levels ↔ no FDR-corrected significant correlations was observed between changes in functional activity and electric fields.<br>Frontoparietal connectivity ↔ significant positive correlation with electric field at frontopolar stimulation site<br>Network-based analysis ↔ revealed dependency of tDCS effects on individual regional current dose |
| Dayal et al.,(2024)<br>88 | Alcohol | 22 | Detoxification | 10 | 3 months | Sham controlled and another therapy | Parallel | EEG System | left DLPFC-Anodal tDCS | 25 | 25 | 20 | 2 | Obsessive Compulsive Drinking Scale (OCDS) scores ↓ over time; however, treatment and treatment × time interaction ↔ no significant effects on OCDS scores<br>Complete abstinence rates ↔ no significant differences between active and sham treatment groups |

**Abbreviations:** tES: transcranial electrical stimulation, tDCS: transcranial direct current stimulation, tACS: transcranial alternating current stimulation, EEG: electroencephalography, MRI: magnetic resonance imaging, DLPFC: dorsolateral prefrontal cortex, FPC: frontopolar cortex, IFG: inferior frontal gyrus, TPC: temporoparietal cortex, NA: not available.

**Table S.3.**
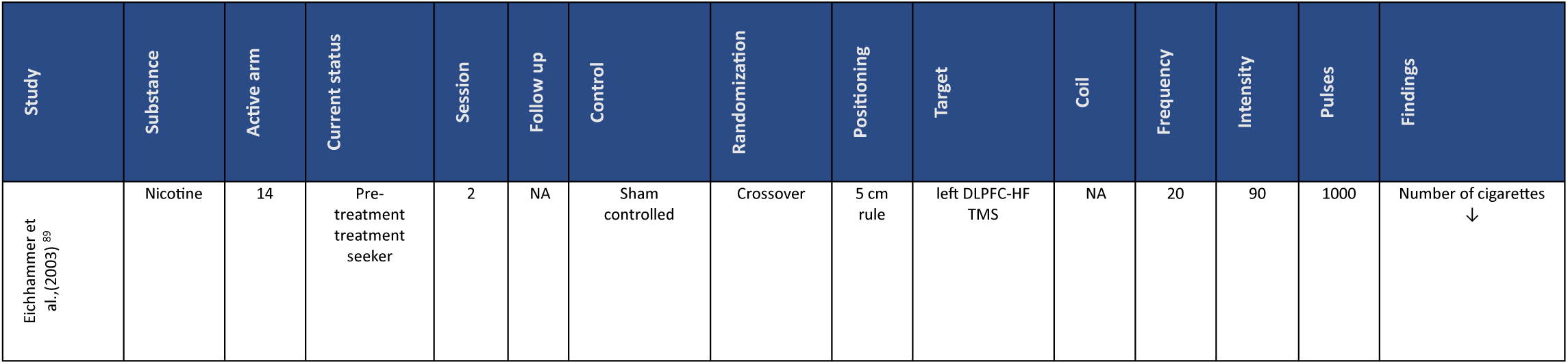
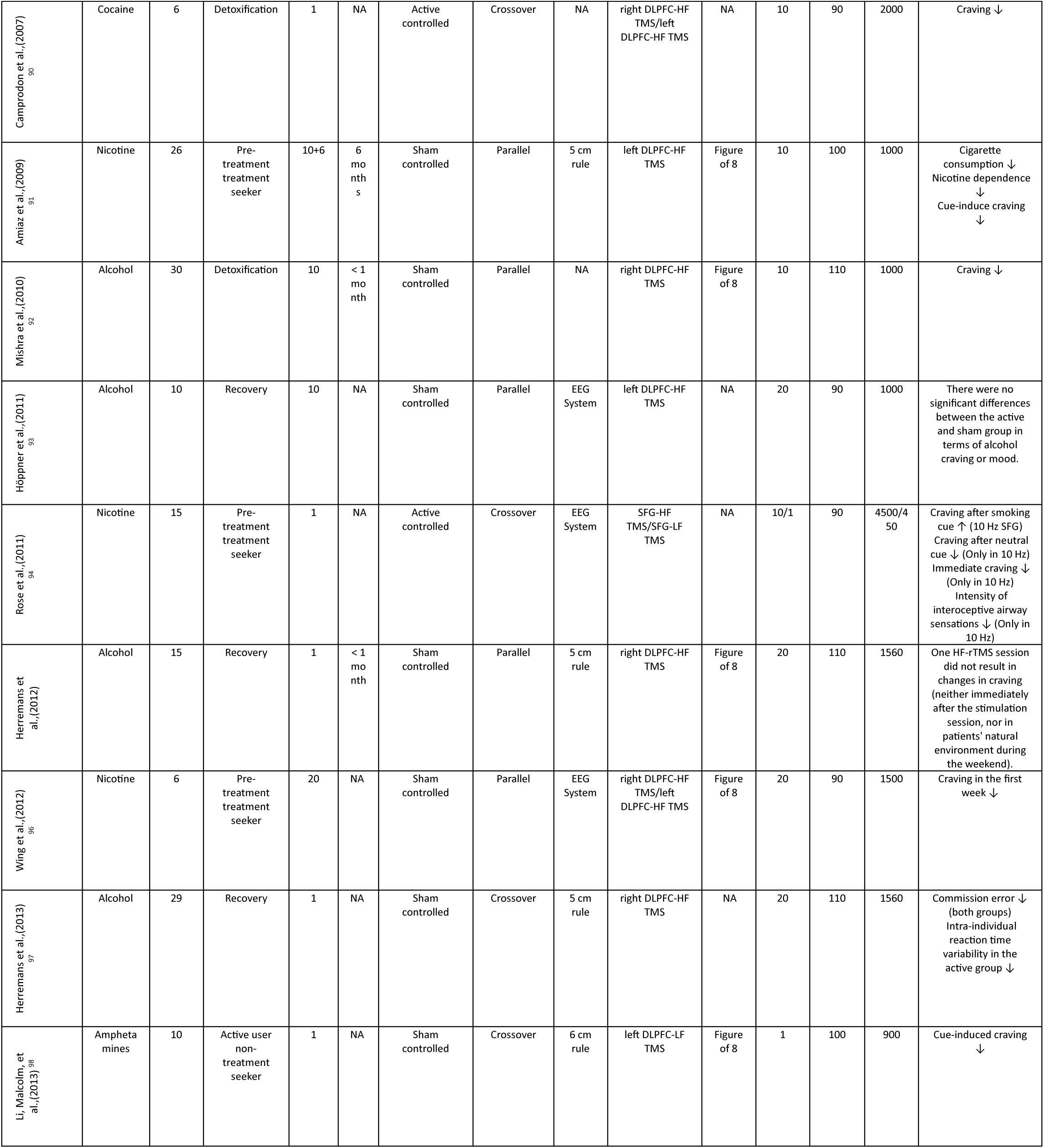
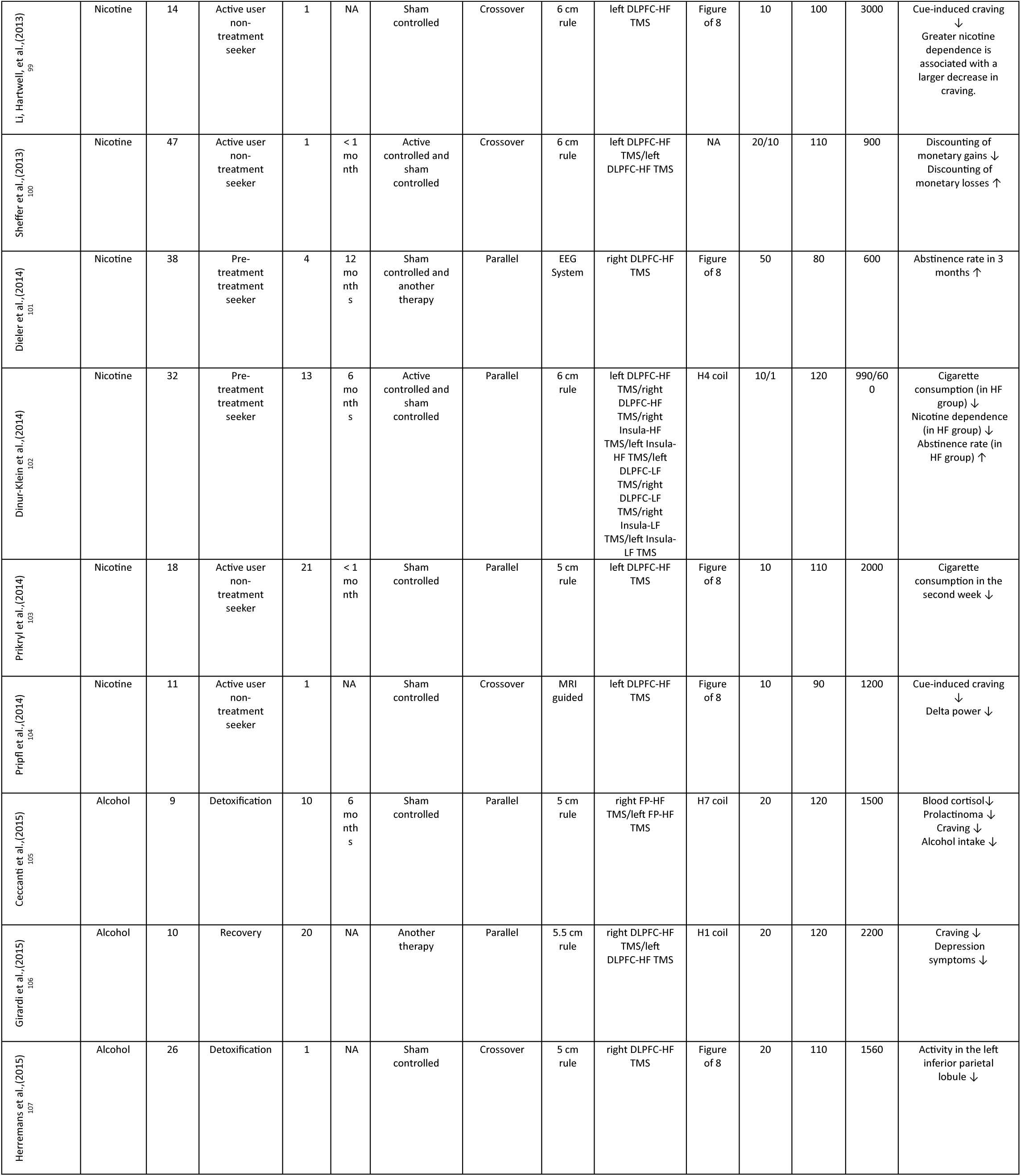
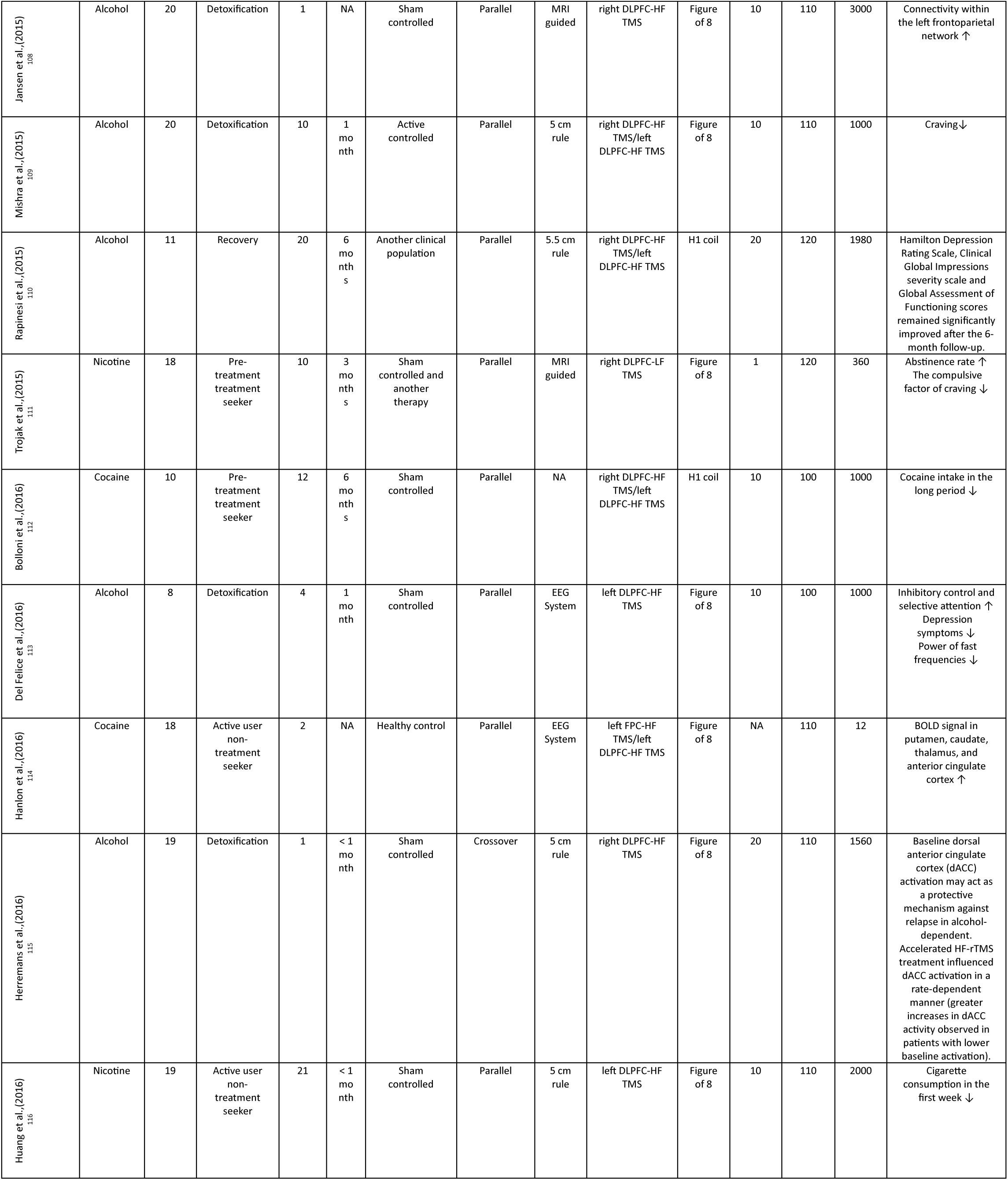
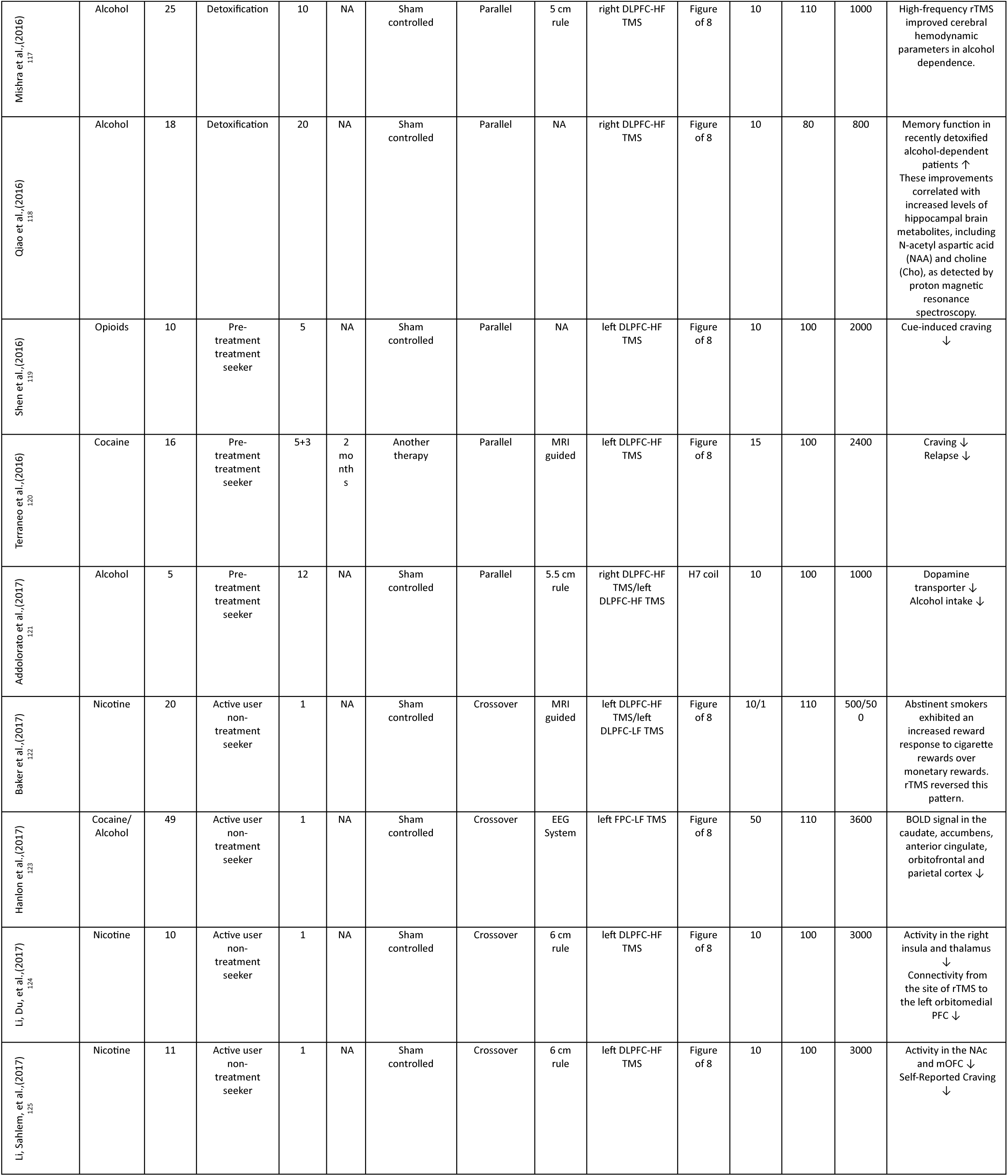
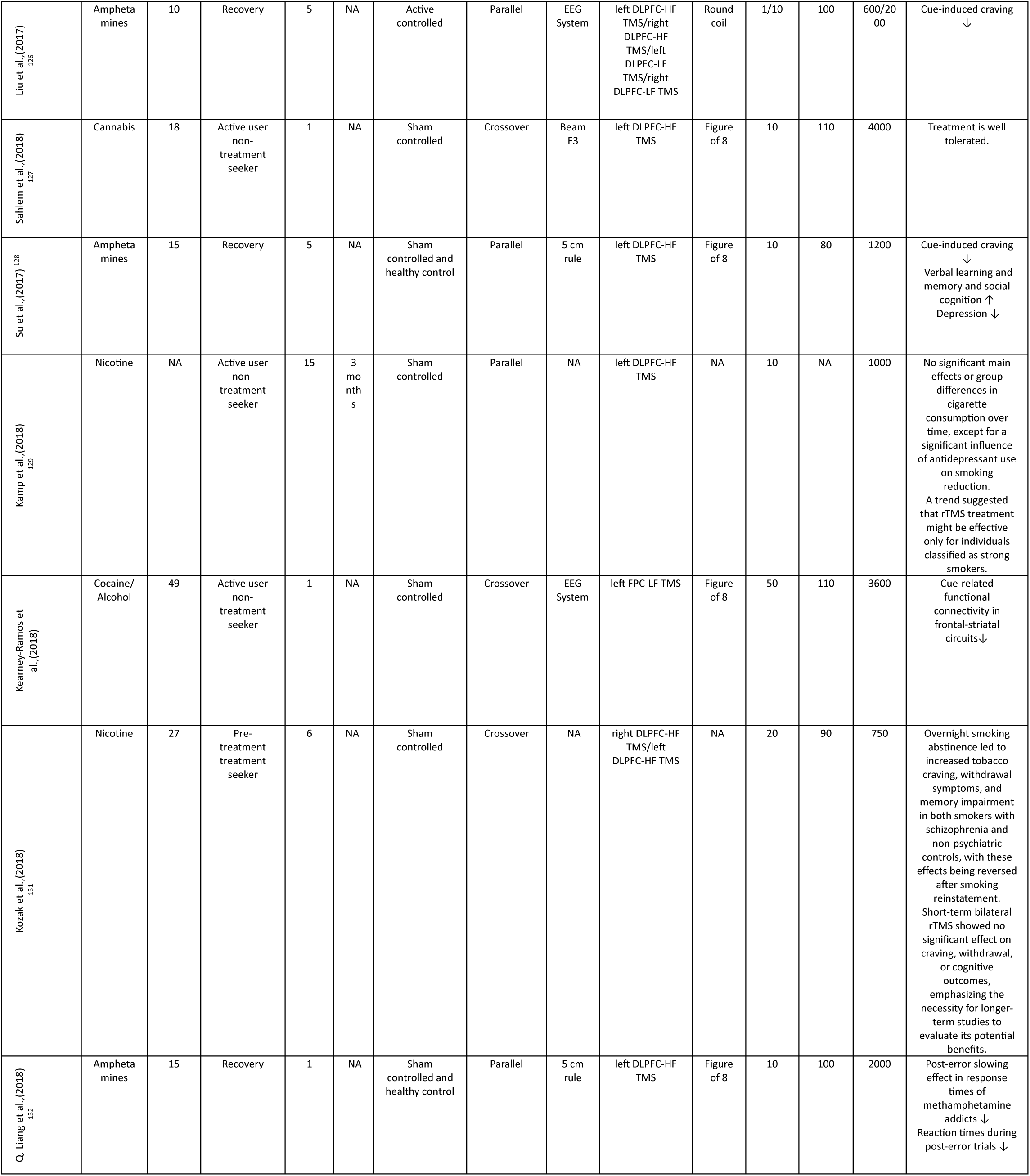
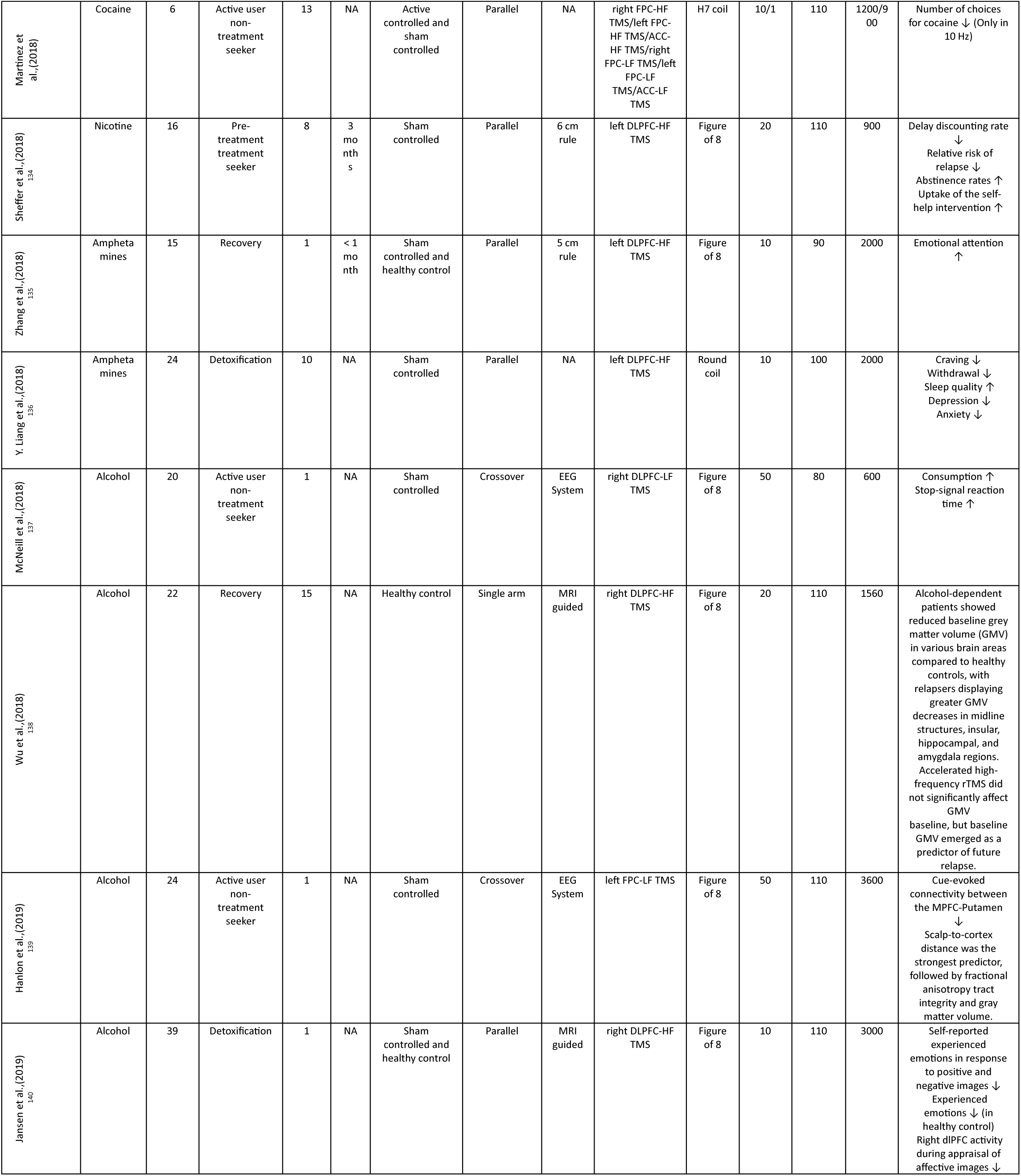
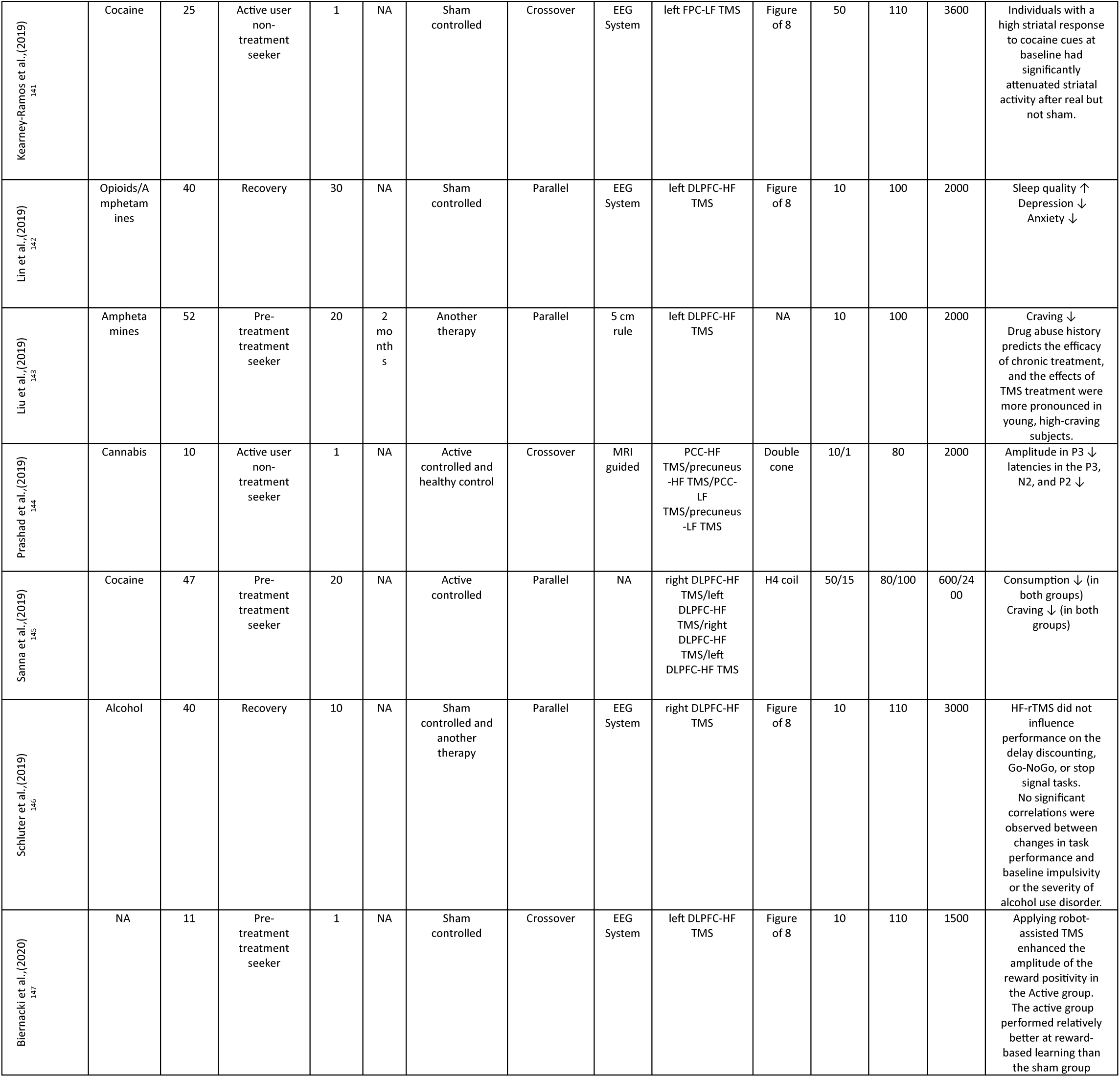
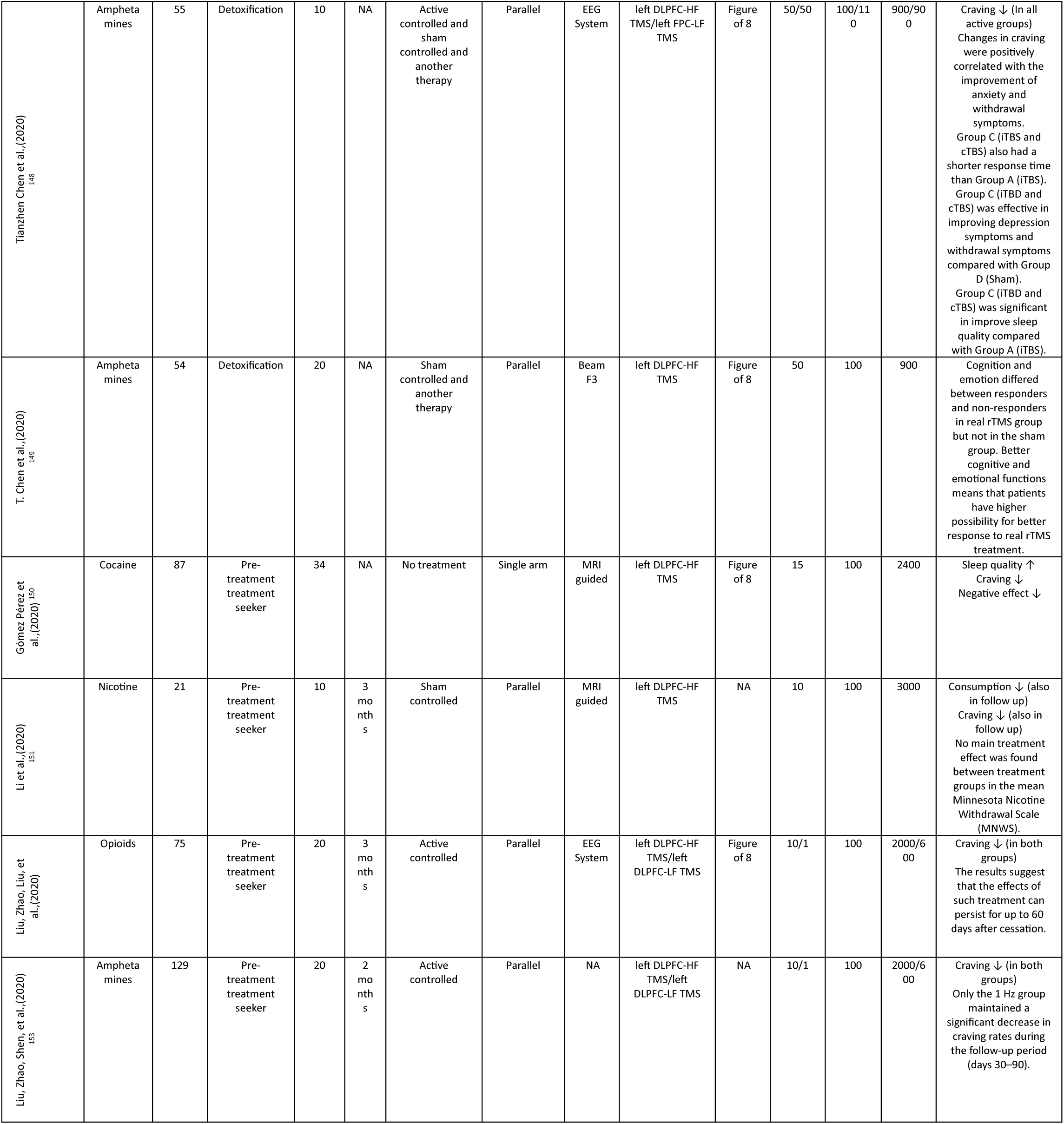
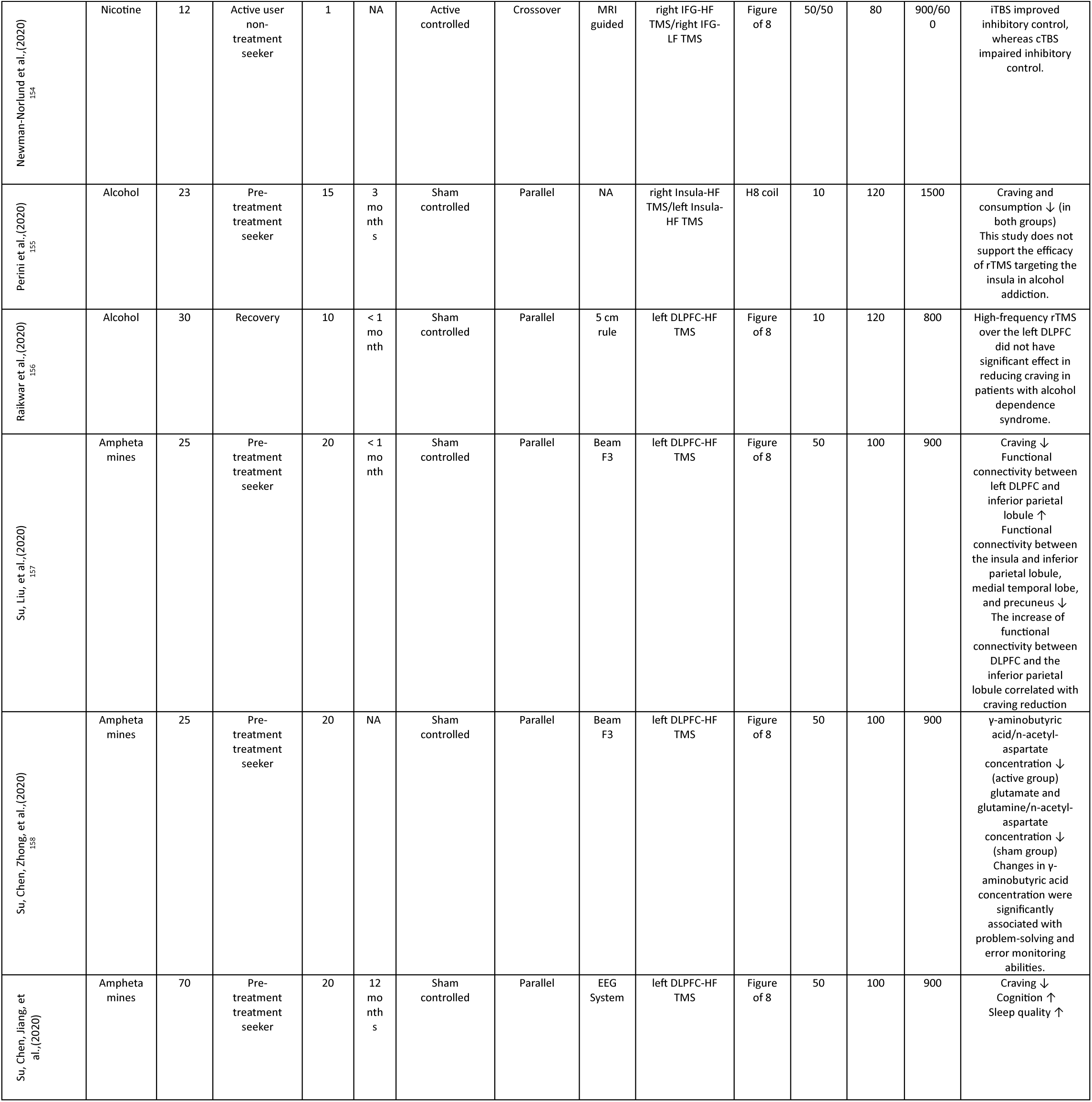
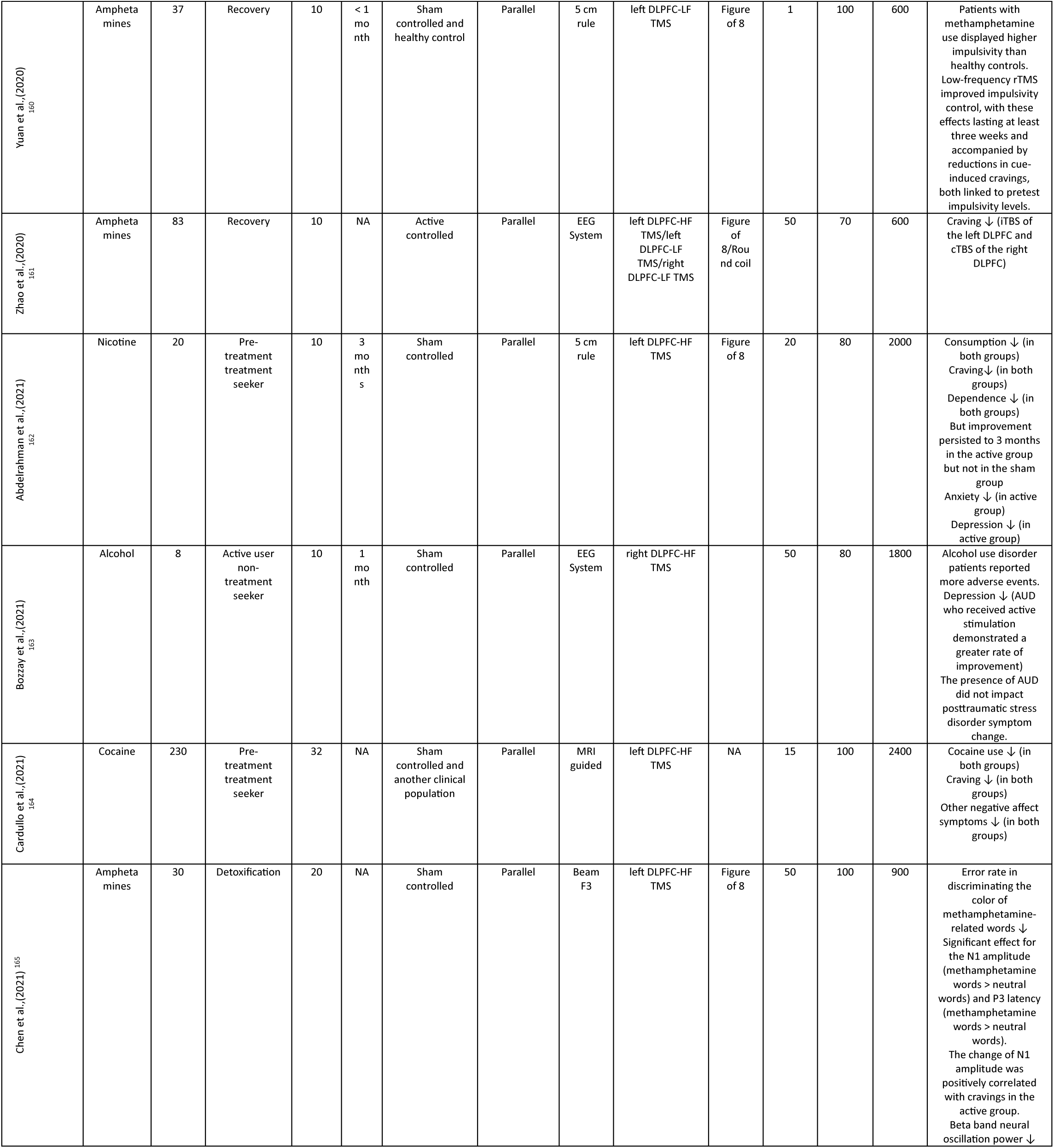
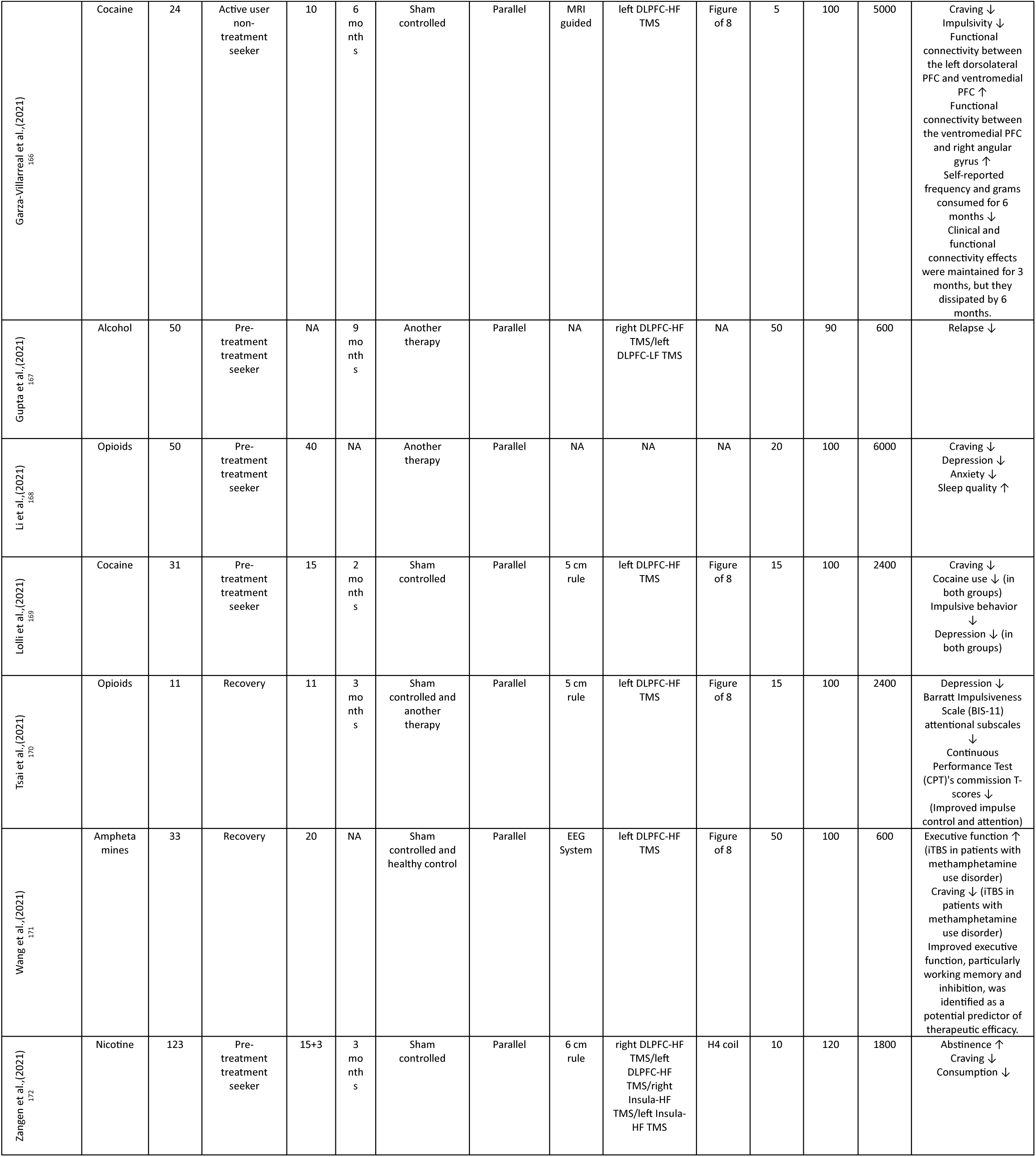
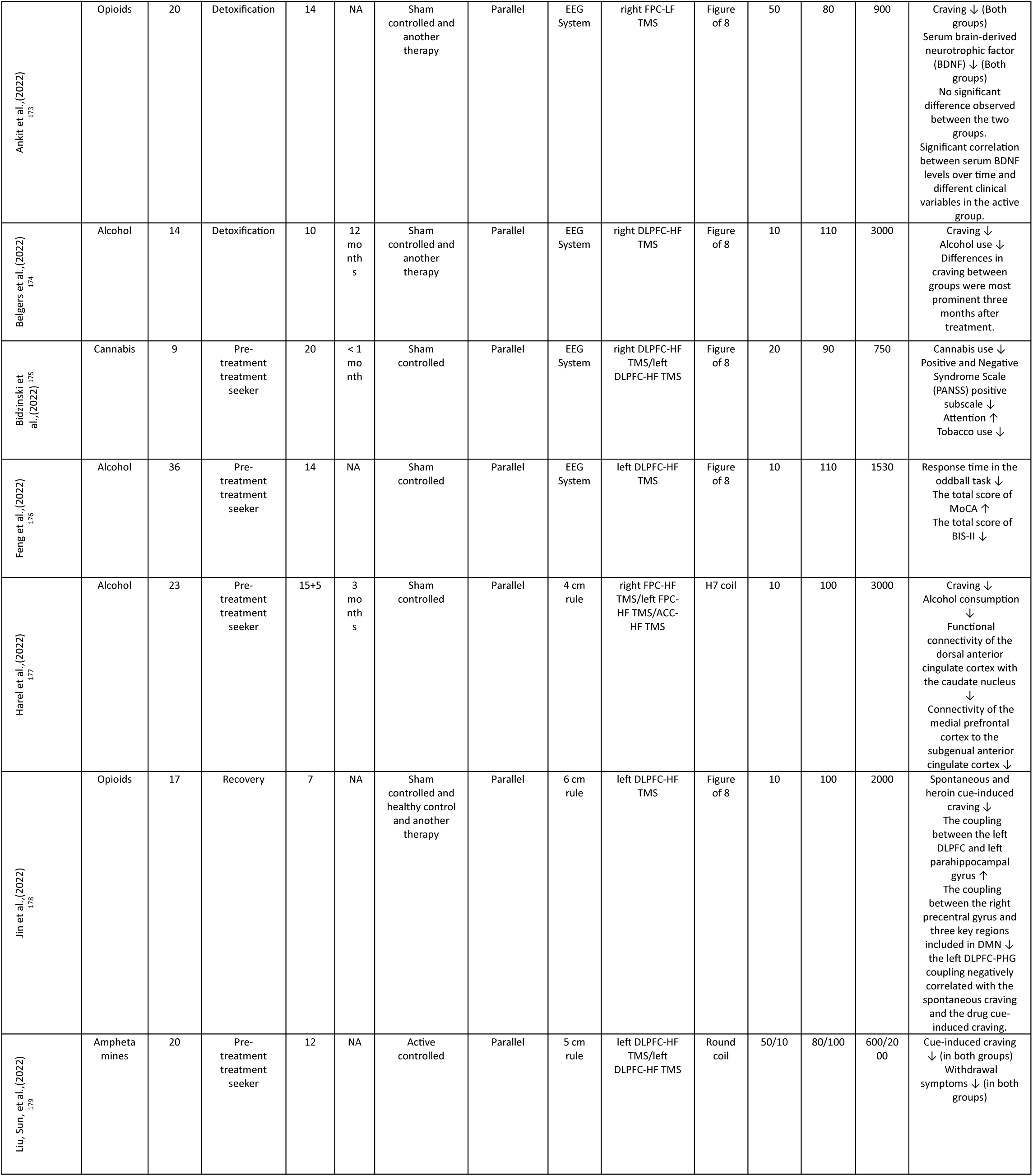
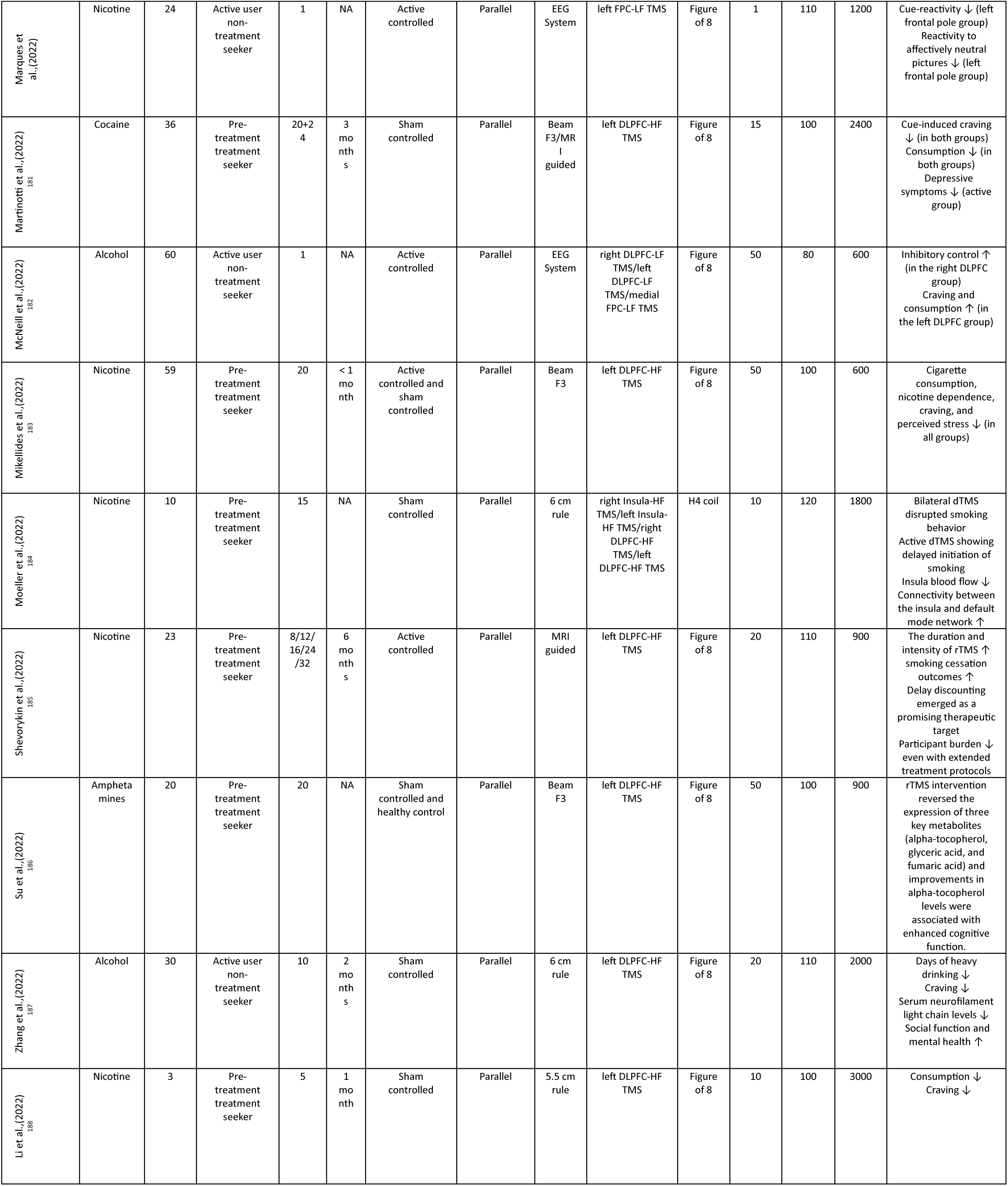
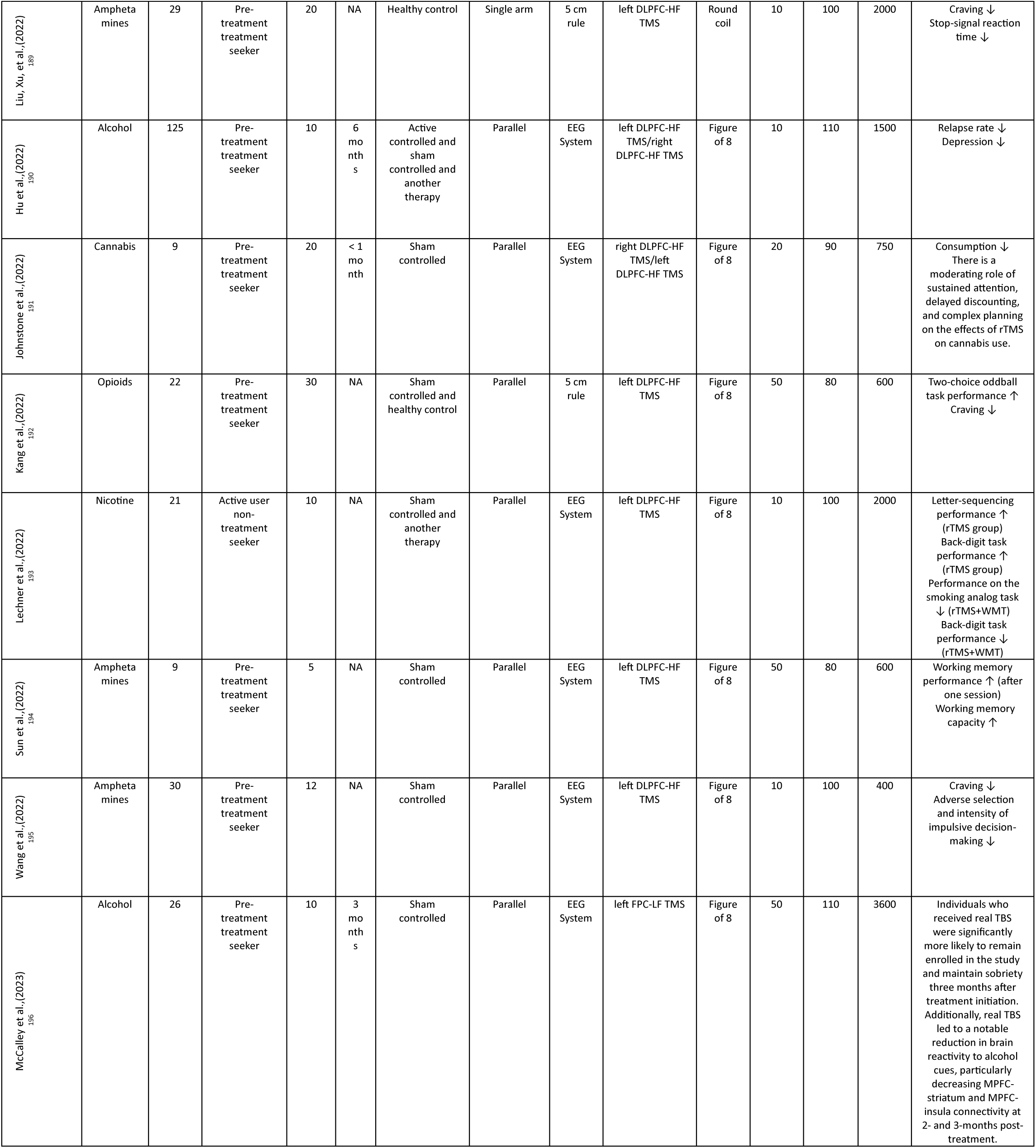
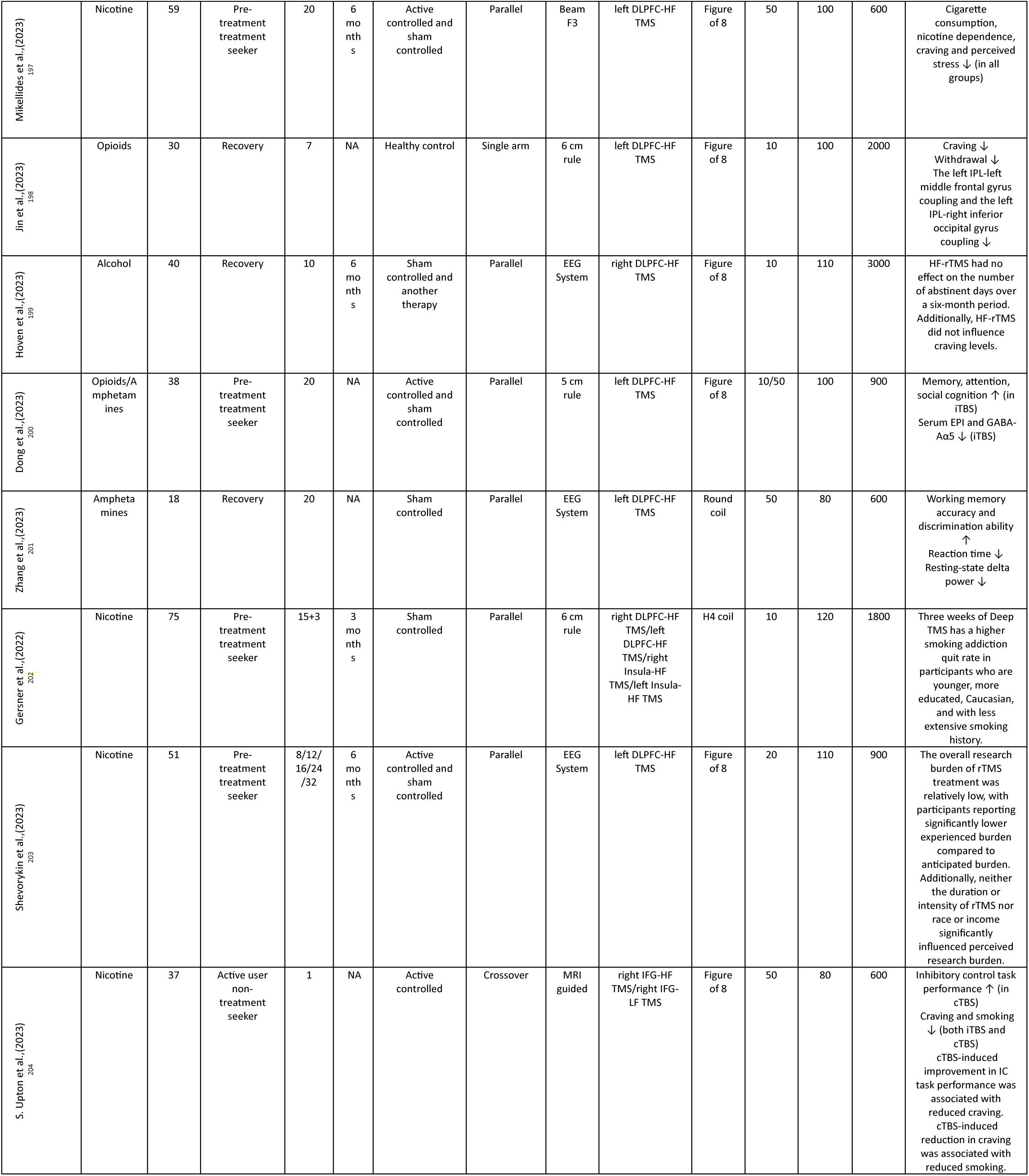
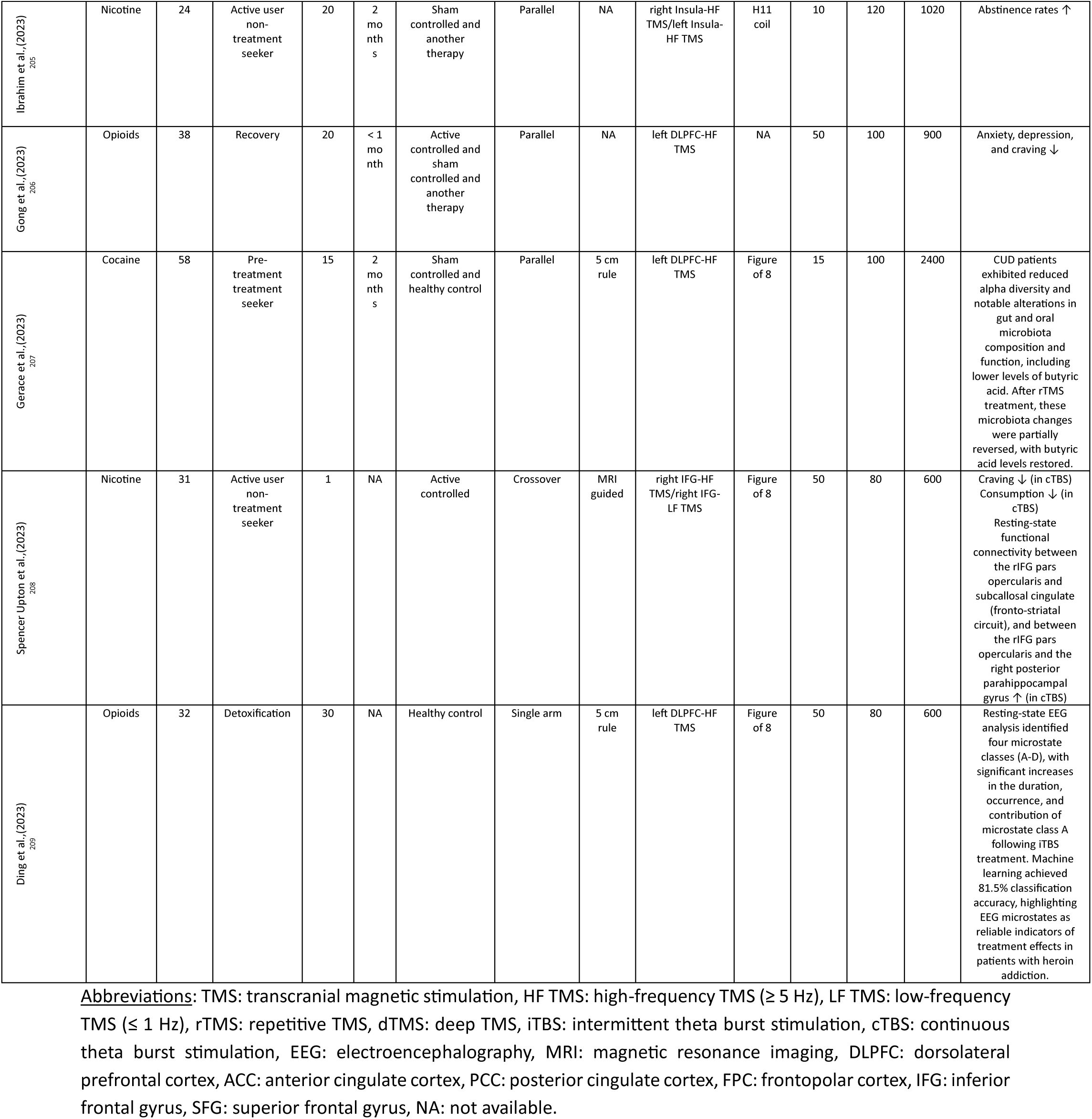
Data Extraction for TMS Studies.

| Study | Substance | Active arm | Current status | Session | Follow up | Control | Randomization | Positioning | Target | Coil | Frequency | Intensity | Pulses | Findings |
| --- | --- | --- | --- | --- | --- | --- | --- | --- | --- | --- | --- | --- | --- | --- |
| Eichhammer et al.,(2003)<br>85 | Nicotine | 14 | Pre-treatment treatment seeker | 2 | NA | Sham controlled | Crossover | 5 cm rule | left DLPFC-HF TMS | NA | 20 | 90 | 1000 | Number of cigarettes ↓ |
| Camprodon et al.,(2007)<br>30 | Cocaine | 6 | Detoxification | 1 | NA | Active controlled | Crossover | NA | right DLPFC-HF TMS/left DLPFC-HF TMS | NA | 10 | 90 | 2000 | Craving ↓ |
| Amiaz et al.,(2009)<br>31 | Nicotine | 26 | Pre-treatment treatment seeker | 10+6 | 6 months | Sham controlled | Parallel | 5 cm rule | left DLPFC-HF TMS | Figure of 8 | 10 | 100 | 1000 | Cigarette consumption ↓<br>Nicotine dependence ↓<br>Cue-induced craving ↓ |
| Mishra et al.,(2010)<br>32 | Alcohol | 30 | Detoxification | 10 | < 1 month | Sham controlled | Parallel | NA | right DLPFC-HF TMS | Figure of 8 | 10 | 110 | 1000 | Craving ↓ |
| Höppner et al.,(2011)<br>33 | Alcohol | 10 | Recovery | 10 | NA | Sham controlled | Parallel | EEG System | left DLPFC-HF TMS | NA | 20 | 90 | 1000 | There were no significant differences between the active and sham group in terms of alcohol craving or mood. |
| Rose et al.,(2011)<br>34 | Nicotine | 15 | Pre-treatment treatment seeker | 1 | NA | Active controlled | Crossover | EEG System | SFG-HF TMS/SFG-LF TMS | NA | 10/1 | 90 | 4500/450 | Craving after smoking cue ↑ (10 Hz SFG)<br>Craving after neutral cue ↓ (Only in 10 Hz)<br>Immediate craving ↓ (Only in 10 Hz)<br>Intensity of interoceptive airway sensations ↓ (Only in 10 Hz) |
| Herremans et al.,(2012) | Alcohol | 15 | Recovery | 1 | < 1 month | Sham controlled | Parallel | 5 cm rule | right DLPFC-HF TMS | Figure of 8 | 20 | 110 | 1560 | One HF-rTMS session did not result in changes in craving (neither immediately after the stimulation session, nor in patients' natural environment during the weekend). |
| Wing et al.,(2012)<br>36 | Nicotine | 6 | Pre-treatment treatment seeker | 20 | NA | Sham controlled | Parallel | EEG System | right DLPFC-HF TMS/left DLPFC-HF TMS | Figure of 8 | 20 | 90 | 1500 | Craving in the first week ↓ |
| Herremans et al.,(2013)<br>37 | Alcohol | 29 | Recovery | 1 | NA | Sham controlled | Crossover | 5 cm rule | right DLPFC-HF TMS | NA | 20 | 110 | 1560 | Commission error ↓ (both groups)<br>Intra-individual reaction time variability in the active group ↓ |
| Li, Malcolm, et al.,(2013)<br>38 | Amphetamine | 10 | Active user non-treatment seeker | 1 | NA | Sham controlled | Crossover | 6 cm rule | left DLPFC-LF TMS | Figure of 8 | 1 | 100 | 900 | Cue-induced craving ↓ |
| Li, Hartwell, et al., (2013)<br><sup>99</sup> | Nicotine | 14 | Active user non-treatment seeker | 1 | NA | Sham controlled | Crossover | 6 cm rule | left DLPFC-HF TMS | Figure of 8 | 10 | 100 | 3000 | Cue-induced craving ↓<br>Greater nicotine dependence is associated with a larger decrease in craving. |
| Sheffer et al., (2013)<br><sup>100</sup> | Nicotine | 47 | Active user non-treatment seeker | 1 | < 1 month | Active controlled and sham controlled | Crossover | 6 cm rule | left DLPFC-HF TMS/left DLPFC-HF TMS | NA | 20/10 | 110 | 900 | Discounting of monetary gains ↓<br>Discounting of monetary losses ↑ |
| Dieler et al., (2014)<br><sup>101</sup> | Nicotine | 38 | Pre-treatment treatment seeker | 4 | 12 months | Sham controlled and another therapy | Parallel | EEG System | right DLPFC-HF TMS | Figure of 8 | 50 | 80 | 600 | Abstinence rate in 3 months ↑ |
| Dinur-Klein et al., (2014)<br><sup>102</sup> | Nicotine | 32 | Pre-treatment treatment seeker | 13 | 6 months | Active controlled and sham controlled | Parallel | 6 cm rule | left DLPFC-HF TMS/right DLPFC-HF TMS/right Insula-HF TMS/left Insula-HF TMS/left DLPFC-LF TMS/right DLPFC-LF TMS/right Insula-LF TMS/left Insula-LF TMS | H4 coil | 10/1 | 120 | 990/600 | Cigarette consumption (in HF group) ↓<br>Nicotine dependence (in HF group) ↓<br>Abstinence rate (in HF group) ↑ |
| Prikyl et al., (2014)<br><sup>103</sup> | Nicotine | 18 | Active user non-treatment seeker | 21 | < 1 month | Sham controlled | Parallel | 5 cm rule | left DLPFC-HF TMS | Figure of 8 | 10 | 110 | 2000 | Cigarette consumption in the second week ↓ |
| Pripfl et al., (2014)<br><sup>104</sup> | Nicotine | 11 | Active user non-treatment seeker | 1 | NA | Sham controlled | Crossover | MRI guided | left DLPFC-HF TMS | Figure of 8 | 10 | 90 | 1200 | Cue-induced craving ↓<br>Delta power ↓ |
| Ceccanti et al., (2015)<br><sup>105</sup> | Alcohol | 9 | Detoxification | 10 | 6 months | Sham controlled | Parallel | 5 cm rule | right FP-HF TMS/left FP-HF TMS | H7 coil | 20 | 120 | 1500 | Blood cortisol ↓<br>Prolactinoma ↓<br>Craving ↓<br>Alcohol intake ↓ |
| Girardi et al., (2015)<br><sup>106</sup> | Alcohol | 10 | Recovery | 20 | NA | Another therapy | Parallel | 5.5 cm rule | right DLPFC-HF TMS/left DLPFC-HF TMS | H1 coil | 20 | 120 | 2200 | Craving ↓<br>Depression symptoms ↓ |
| Herremans et al., (2015)<br><sup>107</sup> | Alcohol | 26 | Detoxification | 1 | NA | Sham controlled | Crossover | 5 cm rule | right DLPFC-HF TMS | Figure of 8 | 20 | 110 | 1560 | Activity in the left inferior parietal lobule ↓ |
| Jansen et al., (2015)<br><sup>108</sup> | Alcohol | 20 | Detoxification | 1 | NA | Sham controlled | Parallel | MRI guided | right DLPFC-HF TMS | Figure of 8 | 10 | 110 | 3000 | Connectivity within the left frontoparietal network ↑ |
| Mishra et al., (2015)<br><sup>109</sup> | Alcohol | 20 | Detoxification | 10 | 1 month | Active controlled | Parallel | 5 cm rule | right DLPFC-HF TMS/left DLPFC-HF TMS | Figure of 8 | 10 | 110 | 1000 | Craving ↓ |
| Raphnesi et al., (2015)<br><sup>110</sup> | Alcohol | 11 | Recovery | 20 | 6 months | Another clinical population | Parallel | 5.5 cm rule | right DLPFC-HF TMS/left DLPFC-HF TMS | H1 coil | 20 | 120 | 1980 | Hamilton Depression Rating Scale, Clinical Global Impressions severity scale and Global Assessment of Functioning scores remained significantly improved after the 6-month follow-up. |
| Trojacek et al., (2015)<br><sup>111</sup> | Nicotine | 18 | Pre-treatment treatment seeker | 10 | 3 months | Sham controlled and another therapy | Parallel | MRI guided | right DLPFC-LF TMS | Figure of 8 | 1 | 120 | 360 | Abstinence rate ↑<br>The compulsive factor of craving ↓ |
| Bolloni et al., (2016)<br><sup>112</sup> | Cocaine | 10 | Pre-treatment treatment seeker | 12 | 6 months | Sham controlled | Parallel | NA | right DLPFC-HF TMS/left DLPFC-HF TMS | H1 coil | 10 | 100 | 1000 | Cocaine intake in the long period ↓ |
| Del Felice et al., (2016)<br><sup>113</sup> | Alcohol | 8 | Detoxification | 4 | 1 month | Sham controlled | Parallel | EEG System | left DLPFC-HF TMS | Figure of 8 | 10 | 100 | 1000 | Inhibitory control and selective attention ↑<br>Depression symptoms ↓<br>Power of fast frequencies ↓ |
| Hanlon et al., (2016)<br><sup>114</sup> | Cocaine | 18 | Active user non-treatment seeker | 2 | NA | Healthy control | Parallel | EEG System | left FPC-HF TMS/left DLPFC-HF TMS | Figure of 8 | NA | 110 | 12 | BOLD signal in putamen, caudate, thalamus, and anterior cingulate cortex ↑ |
| Herremans et al., (2016)<br><sup>115</sup> | Alcohol | 19 | Detoxification | 1 | < 1 month | Sham controlled | Crossover | 5 cm rule | right DLPFC-HF TMS | Figure of 8 | 20 | 110 | 1560 | Baseline dorsal anterior cingulate cortex (dACC) activation may act as a protective mechanism against relapse in alcohol-dependent. Accelerated HF-rTMS treatment influenced dACC activation in a rate-dependent manner (greater increases in dACC activity observed in patients with lower baseline activation). |
| Huang et al., (2016)<br><sup>116</sup> | Nicotine | 19 | Active user non-treatment seeker | 21 | < 1 month | Sham controlled | Parallel | 5 cm rule | left DLPFC-HF TMS | Figure of 8 | 10 | 110 | 2000 | Cigarette consumption in the first week ↓ |
| Mishra et al.,(2016)<br><sup>117</sup> | Alcohol | 25 | Detoxification | 10 | NA | Sham controlled | Parallel | 5 cm rule | right DLPFC-HF TMS | Figure of 8 | 10 | 110 | 1000 | High-frequency rTMS improved cerebral hemodynamic parameters in alcohol dependence. |
| Qiao et al.,(2016)<br><sup>118</sup> | Alcohol | 18 | Detoxification | 20 | NA | Sham controlled | Parallel | NA | right DLPFC-HF TMS | Figure of 8 | 10 | 80 | 800 | Memory function in recently detoxified alcohol-dependent patients ↑<br>These improvements correlated with increased levels of hippocampal brain metabolites, including N-acetyl aspartic acid (NAA) and choline (Cho), as detected by proton magnetic resonance spectroscopy. |
| Shen et al.,(2016)<br><sup>119</sup> | Opioids | 10 | Pre-treatment treatment seeker | 5 | NA | Sham controlled | Parallel | NA | left DLPFC-HF TMS | Figure of 8 | 10 | 100 | 2000 | Cue-induced craving ↓ |
| Terraneo et al.,(2016)<br><sup>120</sup> | Cocaine | 16 | Pre-treatment treatment seeker | 5+3 | 2 months | Another therapy | Parallel | MRI guided | left DLPFC-HF TMS | Figure of 8 | 15 | 100 | 2400 | Craving ↓<br>Relapse ↓ |
| Addolorato et al.,(2017)<br><sup>121</sup> | Alcohol | 5 | Pre-treatment treatment seeker | 12 | NA | Sham controlled | Parallel | 5.5 cm rule | right DLPFC-HF TMS/left DLPFC-HF TMS | H7 coil | 10 | 100 | 1000 | Dopamine transporter ↓<br>Alcohol intake ↓ |
| Baker et al.,(2017)<br><sup>122</sup> | Nicotine | 20 | Active user non-treatment seeker | 1 | NA | Sham controlled | Crossover | MRI guided | left DLPFC-HF TMS/left DLPFC-LF TMS | Figure of 8 | 10/1 | 110 | 500/500 | Abstinent smokers exhibited an increased reward response to cigarette rewards over monetary rewards. rTMS reversed this pattern. |
| Hanlon et al.,(2017)<br><sup>123</sup> | Cocaine/Alcohol | 49 | Active user non-treatment seeker | 1 | NA | Sham controlled | Crossover | EEG System | left FPC-LF TMS | Figure of 8 | 50 | 110 | 3600 | BOLD signal in the caudate, accumbens, anterior cingulate, orbitofrontal and parietal cortex ↓ |
| Li, Du, et al.,(2017)<br><sup>124</sup> | Nicotine | 10 | Active user non-treatment seeker | 1 | NA | Sham controlled | Crossover | 6 cm rule | left DLPFC-HF TMS | Figure of 8 | 10 | 100 | 3000 | Activity in the right insula and thalamus ↓<br>Connectivity from the site of rTMS to the left orbitomedial PFC ↓ |
| Li, Sahlem, et al.,(2017)<br><sup>125</sup> | Nicotine | 11 | Active user non-treatment seeker | 1 | NA | Sham controlled | Crossover | 6 cm rule | left DLPFC-HF TMS | Figure of 8 | 10 | 100 | 3000 | Activity in the Nac and mOFC ↓<br>Self-Reported Craving ↓ |
| Liu et al., (2017)<br><sup>126</sup> | Ampheta<br>mines | 10 | Recovery | 5 | NA | Active<br>controlled | Parallel | EEG<br>System | left DLPFC-HF<br>TMS/right<br>DLPFC-HF<br>TMS/left<br>DLPFC-LF<br>TMS/right<br>DLPFC-LF TMS | Round<br>coil | 1/10 | 100 | 600/20<br>00 | Cue-induced craving<br>↓ |
| Sahlem et al., (2018)<br><sup>127</sup> | Cannabis | 18 | Active user<br>non-treatment<br>seeker | 1 | NA | Sham<br>controlled | Crossover | Beam<br>F3 | left DLPFC-HF<br>TMS | Figure<br>of 8 | 10 | 110 | 4000 | Treatment is well<br>tolerated. |
| Su et al., (2017)<br><sup>128</sup> | Ampheta<br>mines | 15 | Recovery | 5 | NA | Sham<br>controlled and<br>healthy control | Parallel | 5 cm<br>rule | left DLPFC-HF<br>TMS | Figure<br>of 8 | 10 | 80 | 1200 | Cue-induced craving<br>↓<br>Verbal learning and<br>memory and social<br>cognition ↑<br>Depression ↓ |
| Kamp et al., (2018)<br><sup>129</sup> | Nicotine | NA | Active user<br>non-treatment<br>seeker | 15 | 3<br>mo<br>nth<br>s | Sham<br>controlled | Parallel | NA | left DLPFC-HF<br>TMS | NA | 10 | NA | 1000 | No significant main<br>effects or group<br>differences in<br>cigarette<br>consumption over<br>time, except for a<br>significant influence<br>of antidepressant use<br>on smoking<br>reduction.<br>A trend suggested<br>that rTMS treatment<br>might be effective<br>only for individuals<br>classified as strong<br>smokers. |
| Kearney-Ramos et<br>al., (2018) | Cocaine/<br>Alcohol | 49 | Active user<br>non-treatment<br>seeker | 1 | NA | Sham<br>controlled | Crossover | EEG<br>System | left FPC-LF TMS | Figure<br>of 8 | 50 | 110 | 3600 | Cue-related<br>functional<br>connectivity in<br>frontal-striatal<br>circuits ↓ |
| Kozak et al., (2018)<br><sup>131</sup> | Nicotine | 27 | Pre-treatment<br>treatment<br>seeker | 6 | NA | Sham<br>controlled | Crossover | NA | right DLPFC-HF<br>TMS/left<br>DLPFC-HF TMS | NA | 20 | 90 | 750 | Overnight smoking<br>abstinence led to<br>increased tobacco<br>craving, withdrawal<br>symptoms, and<br>memory impairment<br>in both smokers with<br>schizophrenia and<br>non-psychiatric<br>controls, with these<br>effects being reversed<br>after smoking<br>reinstatement.<br>Short-term bilateral<br>rTMS showed no<br>significant effect on<br>craving, withdrawal,<br>or cognitive<br>outcomes,<br>emphasizing the<br>necessity for longer-term studies to<br>evaluate its potential<br>benefits. |
| Q. Liang et al., (2018)<br><sup>132</sup> | Ampheta<br>mines | 15 | Recovery | 1 | NA | Sham<br>controlled and<br>healthy control | Parallel | 5 cm<br>rule | left DLPFC-HF<br>TMS | Figure<br>of 8 | 10 | 100 | 2000 | Post-error slowing<br>effect in response<br>times of<br>methamphetamine<br>addicts ↓<br>Reaction times during<br>post-error trials ↓ |
| Martinez et al.,(2018) | Cocaine | 6 | Active user non-treatment seeker | 13 | NA | Active controlled and sham controlled | Parallel | NA | right FPC-HF TMS/left FPC-HF TMS/ACC-HF TMS/right FPC-LF TMS/left FPC-LF TMS/ACC-LF TMS | H7 coil | 10/1 | 110 | 1200/900 | Number of choices for cocaine ↓ (Only in 10 Hz) |
| Sheffer et al.,(2018)<br><sup>134</sup> | Nicotine | 16 | Pre-treatment treatment seeker | 8 | 3 months | Sham controlled | Parallel | 6 cm rule | left DLPFC-HF TMS | Figure of 8 | 20 | 110 | 900 | Delay discounting rate ↓<br>Relative risk of relapse ↓<br>Abstinence rates ↑<br>Uptake of the self-help intervention ↑ |
| Zhang et al.,(2018)<br><sup>135</sup> | Ampheta mines | 15 | Recovery | 1 | < 1 month | Sham controlled and healthy control | Parallel | 5 cm rule | left DLPFC-HF TMS | Figure of 8 | 10 | 90 | 2000 | Emotional attention ↑ |
| Y. Liang et al.,(2018)<br><sup>136</sup> | Ampheta mines | 24 | Detoxification | 10 | NA | Sham controlled | Parallel | NA | left DLPFC-HF TMS | Round coil | 10 | 100 | 2000 | Craving ↓<br>Withdrawal ↓<br>Sleep quality ↑<br>Depression ↓<br>Anxiety ↓ |
| McNeill et al.,(2018)<br><sup>137</sup> | Alcohol | 20 | Active user non-treatment seeker | 1 | NA | Sham controlled | Crossover | EEG System | right DLPFC-LF TMS | Figure of 8 | 50 | 80 | 600 | Consumption ↑<br>Stop-signal reaction time ↑ |
| Wu et al.,(2018)<br><sup>138</sup> | Alcohol | 22 | Recovery | 15 | NA | Healthy control | Single arm | MRI guided | right DLPFC-HF TMS | Figure of 8 | 20 | 110 | 1560 | Alcohol-dependent patients showed reduced baseline grey matter volume (GMV) in various brain areas compared to healthy controls, with relapsers displaying greater GMV decreases in midline structures, insular, hippocampal, and amygdala regions. Accelerated high-frequency rTMS did not significantly affect GMV baseline, but baseline GMV emerged as a predictor of future relapse. |
| Hanlon et al.,(2019)<br><sup>139</sup> | Alcohol | 24 | Active user non-treatment seeker | 1 | NA | Sham controlled | Crossover | EEG System | left FPC-LF TMS | Figure of 8 | 50 | 110 | 3600 | Cue-evoked connectivity between the MPFC-Putamen ↓<br>Scalp-to-cortex distance was the strongest predictor, followed by fractional anisotropy tract integrity and gray matter volume. |
| Jansen et al.,(2019)<br><sup>140</sup> | Alcohol | 39 | Detoxification | 1 | NA | Sham controlled and healthy control | Parallel | MRI guided | right DLPFC-HF TMS | Figure of 8 | 10 | 110 | 3000 | Self-reported experienced emotions in response to positive and negative images ↓<br>Experienced emotions ↓ (in healthy control)<br>Right dlPFC activity during appraisal of affective images ↓ |
| Keamey-Ramos et al., (2019)<br>141 | Cocaine | 25 | Active user non-treatment seeker | 1 | NA | Sham controlled | Crossover | EEG System | left FPC-LF TMS | Figure of 8 | 50 | 110 | 3600 | Individuals with a high striatal response to cocaine cues at baseline had significantly attenuated striatal activity after real but not sham. |
| Lin et al., (2019)<br>142 | Opioids/Amphetamines | 40 | Recovery | 30 | NA | Sham controlled | Parallel | EEG System | left DLPFC-HF TMS | Figure of 8 | 10 | 100 | 2000 | Sleep quality ↑<br>Depression ↓<br>Anxiety ↓ |
| Liu et al., (2019)<br>143 | Amphetamines | 52 | Pre-treatment treatment seeker | 20 | 2 months | Another therapy | Parallel | 5 cm rule | left DLPFC-HF TMS | NA | 10 | 100 | 2000 | Craving ↓<br>Drug abuse history predicts the efficacy of chronic treatment, and the effects of TMS treatment were more pronounced in young, high-craving subjects. |
| Prashad et al., (2019)<br>144 | Cannabis | 10 | Active user non-treatment seeker | 1 | NA | Active controlled and healthy control | Crossover | MRI guided | PCC-HF TMS/precuneus -HF TMS/PCC-LF TMS/precuneus -LF TMS | Double cone | 10/1 | 80 | 2000 | Amplitude in P3 ↓<br>latencies in the P3, N2, and P2 ↓ |
| Sanna et al., (2019)<br>145 | Cocaine | 47 | Pre-treatment treatment seeker | 20 | NA | Active controlled | Parallel | NA | right DLPFC-HF TMS/left DLPFC-HF TMS/right DLPFC-HF TMS/left DLPFC-HF TMS | H4 coil | 50/15 | 80/100 | 600/2400 | Consumption ↓ (in both groups)<br>Craving ↓ (in both groups) |
| Schluter et al., (2019)<br>146 | Alcohol | 40 | Recovery | 10 | NA | Sham controlled and another therapy | Parallel | EEG System | right DLPFC-HF TMS | Figure of 8 | 10 | 110 | 3000 | HF-rTMS did not influence performance on the delay discounting, Go-NoGo, or stop signal tasks.<br>No significant correlations were observed between changes in task performance and baseline impulsivity or the severity of alcohol use disorder. |
| Biernacki et al., (2020)<br>147 | NA | 11 | Pre-treatment treatment seeker | 1 | NA | Sham controlled | Crossover | EEG System | left DLPFC-HF TMS | Figure of 8 | 10 | 110 | 1500 | Applying robot-assisted TMS enhanced the amplitude of the reward positivity in the Active group.<br>The active group performed relatively better at reward-based learning than the sham group |
| Tianzhen Chen et al., (2020)<br>148 | Ampheta<br>mines | 55 | Detoxification | 10 | NA | Active<br>controlled and<br>sham<br>controlled and<br>another<br>therapy | Parallel | EEG<br>System | left DLPFC-HF<br>TMS/left FPC-LF<br>TMS | Figure<br>of 8 | 50/50 | 100/11<br>0 | 900/90<br>0 | Craving ↓ (In all<br>active groups)<br>Changes in craving<br>were positively<br>correlated with the<br>improvement of<br>anxiety and<br>withdrawal<br>symptoms.<br>Group C (iTBS and<br>cTBS) also had a<br>shorter response time<br>than Group A (iTBS).<br>Group C (iTBD and<br>cTBS) was effective in<br>improving depression<br>symptoms and<br>withdrawal symptoms<br>compared with Group<br>D (Sham).<br>Group C (iTBD and<br>cTBS) was significant<br>in improve sleep<br>quality compared<br>with Group A (iTBS). |
| T. Chen et al., (2020)<br>149 | Ampheta<br>mines | 54 | Detoxification | 20 | NA | Sham<br>controlled and<br>another<br>therapy | Parallel | Beam<br>F3 | left DLPFC-HF<br>TMS | Figure<br>of 8 | 50 | 100 | 900 | Cognition and<br>emotion differed<br>between responders<br>and non-responders<br>in real rTMS group<br>but not in the sham<br>group. Better<br>cognitive and<br>emotional functions<br>means that patients<br>have higher<br>possibility for better<br>response to real rTMS<br>treatment. |
| Gómez Pérez et<br>al., (2020) 150 | Cocaine | 87 | Pre-treatment<br>treatment<br>seeker | 34 | NA | No treatment | Single arm | MRI<br>guided | left DLPFC-HF<br>TMS | Figure<br>of 8 | 15 | 100 | 2400 | Sleep quality ↑<br>Craving ↓<br>Negative effect ↓ |
| Li et al., (2020)<br>151 | Nicotine | 21 | Pre-treatment<br>treatment<br>seeker | 10 | 3<br>mo<br>nth<br>s | Sham<br>controlled | Parallel | MRI<br>guided | left DLPFC-HF<br>TMS | NA | 10 | 100 | 3000 | Consumption ↓ (also<br>in follow up)<br>Craving ↓ (also in<br>follow up)<br>No main treatment<br>effect was found<br>between treatment<br>groups in the mean<br>Minnesota Nicotine<br>Withdrawal Scale<br>(MNWS). |
| Liu, Zhao, Liu, et<br>al., (2020) | Opioids | 75 | Pre-treatment<br>treatment<br>seeker | 20 | 3<br>mo<br>nth<br>s | Active<br>controlled | Parallel | EEG<br>System | left DLPFC-HF<br>TMS/left<br>DLPFC-LF TMS | Figure<br>of 8 | 10/1 | 100 | 2000/6<br>00 | Craving ↓ (in both<br>groups)<br>The results suggest<br>that the effects of<br>such treatment can<br>persist for up to 60<br>days after cessation. |
| Liu, Zhao, Shen, et al., (2020)<br>153 | Ampheta<br>mines | 129 | Pre-treatment<br>treatment<br>seeker | 20 | 2<br>mo<br>nth<br>s | Active<br>controlled | Parallel | NA | left DLPFC-HF<br>TMS/left<br>DLPFC-LF TMS | NA | 10/1 | 100 | 2000/6<br>00 | Craving ↓ (in both<br>groups)<br>Only the 1 Hz group<br>maintained a<br>significant decrease in<br>craving rates during<br>the follow-up period<br>(days 30–90). |
| Newman-Norlund et al.,(2020)<br>134 | Nicotine | 12 | Active user non-treatment seeker | 1 | NA | Active controlled | Crossover | MRI guided | right IFG-HF TMS/right IFG-LF TMS | Figure of 8 | 50/50 | 80 | 900/600 | ITBS improved inhibitory control, whereas cTBS impaired inhibitory control. |
| Perini et al.,(2020)<br>135 | Alcohol | 23 | Pre-treatment treatment seeker | 15 | 3 months | Sham controlled | Parallel | NA | right insula-HF TMS/left insula-HF TMS | H8 coil | 10 | 120 | 1500 | Craving and consumption ↓ (in both groups)<br>This study does not support the efficacy of rTMS targeting the insula in alcohol addiction. |
| Raikwar et al.,(2020)<br>136 | Alcohol | 30 | Recovery | 10 | < 1 month | Sham controlled | Parallel | 5 cm rule | left DLPFC-HF TMS | Figure of 8 | 10 | 120 | 800 | High-frequency rTMS over the left DLPFC did not have significant effect in reducing craving in patients with alcohol dependence syndrome. |
| Su, Liu, et al.,(2020)<br>137 | Amphetamine | 25 | Pre-treatment treatment seeker | 20 | < 1 month | Sham controlled | Parallel | Beam F3 | left DLPFC-HF TMS | Figure of 8 | 50 | 100 | 900 | Craving ↓<br>Functional connectivity between left DLPFC and inferior parietal lobule ↑<br>Functional connectivity between the insula and inferior parietal lobule, medial temporal lobe, and precuneus ↓<br>The increase of functional connectivity between DLPFC and the inferior parietal lobule correlated with craving reduction |
| Su, Chen, Zhong, et al.,(2020)<br>138 | Amphetamine | 25 | Pre-treatment treatment seeker | 20 | NA | Sham controlled | Parallel | Beam F3 | left DLPFC-HF TMS | Figure of 8 | 50 | 100 | 900 | γ-aminobutyric acid/n-acetyl-aspartate concentration ↓ (active group)<br>glutamate and glutamine/n-acetyl-aspartate concentration ↓ (sham group)<br>Changes in γ-aminobutyric acid concentration were significantly associated with problem-solving and error monitoring abilities. |
| Su, Chen, Jiang, et al.,(2020) | Amphetamine | 70 | Pre-treatment treatment seeker | 20 | 12 months | Sham controlled | Parallel | EEG System | left DLPFC-HF TMS | Figure of 8 | 50 | 100 | 900 | Craving ↓<br>Cognition ↑<br>Sleep quality ↑ |
| Yuan et al., (2020)<br><sup>160</sup> | Ampheta<br>mines | 37 | Recovery | 10 | < 1<br>mo<br>nth | Sham<br>controlled and<br>healthy control | Parallel | 5 cm<br>rule | left DLPFC-LF<br>TMS | Figure<br>of 8 | 1 | 100 | 600 | Patients with<br>methamphetamine<br>use displayed higher<br>impulsivity than<br>healthy controls.<br>Low-frequency rTMS<br>improved impulsivity<br>control, with these<br>effects lasting at least<br>three weeks and<br>accompanied by<br>reductions in cue-induced cravings,<br>both linked to pretest<br>impulsivity levels. |
| Zhao et al., (2020)<br><sup>161</sup> | Ampheta<br>mines | 83 | Recovery | 10 | NA | Active<br>controlled | Parallel | EEG<br>System | left DLPFC-HF<br>TMS/left<br>DLPFC-LF<br>TMS/right<br>DLPFC-LF TMS | Figure<br>of<br>8/Rou<br>nd coil | 50 | 70 | 600 | Craving ↓ (ITBS of<br>the left DLPFC and<br>cTBS of the right<br>DLPFC) |
| Abdelrahman et al., (2021)<br><sup>162</sup> | Nicotine | 20 | Pre-treatment<br>treatment<br>seeker | 10 | 3<br>mo<br>nth<br>s | Sham<br>controlled | Parallel | 5 cm<br>rule | left DLPFC-HF<br>TMS | Figure<br>of 8 | 20 | 80 | 2000 | Consumption ↓ (in<br>both groups)<br>Craving ↓ (in both<br>groups)<br>Dependence ↓ (in<br>both groups)<br>But improvement<br>persisted to 3 months<br>in the active group<br>but not in the sham<br>group<br>Anxiety ↓ (in active<br>group)<br>Depression ↓ (in<br>active group) |
| Bozzay et al., (2021)<br><sup>163</sup> | Alcohol | 8 | Active user<br>non-treatment<br>seeker | 10 | 1<br>mo<br>nth | Sham<br>controlled | Parallel | EEG<br>System | right DLPFC-HF<br>TMS |  | 50 | 80 | 1800 | Alcohol use disorder<br>patients reported<br>more adverse events.<br>Depression ↓ (AUD<br>who received active<br>stimulation<br>demonstrated a<br>greater rate of<br>improvement)<br>The presence of AUD<br>did not impact<br>posttraumatic stress<br>disorder symptom<br>change. |
| Cardullo et al., (2021)<br><sup>164</sup> | Cocaine | 230 | Pre-treatment<br>treatment<br>seeker | 32 | NA | Sham<br>controlled and<br>another clinical<br>population | Parallel | MRI<br>guided | left DLPFC-HF<br>TMS | NA | 15 | 100 | 2400 | Cocaine use ↓ (in<br>both groups)<br>Craving ↓ (in both<br>groups)<br>Other negative affect<br>symptoms ↓ (in both<br>groups) |
| Chen et al., (2021)<br><sup>165</sup> | Ampheta<br>mines | 30 | Detoxification | 20 | NA | Sham<br>controlled | Parallel | Beam<br>F3 | left DLPFC-HF<br>TMS | Figure<br>of 8 | 50 | 100 | 900 | Error rate in<br>discriminating the<br>color of<br>methamphetamine-<br>related words ↓<br>Significant effect for<br>the N1 amplitude<br>(methamphetamine<br>words > neutral<br>words) and P3 latency<br>(methamphetamine<br>words > neutral<br>words).<br>The change of N1<br>amplitude was<br>positively correlated<br>with cravings in the<br>active group.<br>Beta band neural<br>oscillation power ↓ |
| Garza-Villarreal et al., (2021)<br>306 | Cocaine | 24 | Active user non-treatment seeker | 10 | 6 months | Sham controlled | Parallel | MRI guided | left DLPFC-HF TMS | Figure of 8 | 5 | 100 | 5000 | Craving ↓<br>Impulsivity ↓<br>Functional connectivity between the left dorsolateral PFC and ventromedial PFC ↑<br>Functional connectivity between the ventromedial PFC and right angular gyrus ↑<br>Self-reported frequency and grams consumed for 6 months ↓<br>Clinical and functional connectivity effects were maintained for 3 months, but they dissipated by 6 months. |
| Gupta et al., (2021)<br>307 | Alcohol | 50 | Pre-treatment treatment seeker | NA | 9 months | Another therapy | Parallel | NA | right DLPFC-HF TMS/left DLPFC-LF TMS | NA | 50 | 90 | 600 | Relapse ↓ |
| Li et al., (2021)<br>308 | Opioids | 50 | Pre-treatment treatment seeker | 40 | NA | Another therapy | Parallel | NA | NA | NA | 20 | 100 | 6000 | Craving ↓<br>Depression ↓<br>Anxiety ↓<br>Sleep quality ↑ |
| Lolli et al., (2021)<br>309 | Cocaine | 31 | Pre-treatment treatment seeker | 15 | 2 months | Sham controlled | Parallel | 5 cm rule | left DLPFC-HF TMS | Figure of 8 | 15 | 100 | 2400 | Craving ↓<br>Cocaine use ↓ (in both groups)<br>Impulsive behavior ↓<br>Depression ↓ (in both groups) |
| Tsai et al., (2021)<br>310 | Opioids | 11 | Recovery | 11 | 3 months | Sham controlled and another therapy | Parallel | 5 cm rule | left DLPFC-HF TMS | Figure of 8 | 15 | 100 | 2400 | Depression ↓<br>Barratt Impulsiveness Scale (BIS-11) attentional subscales ↓<br>Continuous Performance Test (CPT)'s commission T-scores ↓<br>(Improved impulse control and attention) |
| Wang et al., (2021)<br>311 | Amphetamine | 33 | Recovery | 20 | NA | Sham controlled and healthy control | Parallel | EEG System | left DLPFC-HF TMS | Figure of 8 | 50 | 100 | 600 | Executive function ↑ (iTBS in patients with methamphetamine use disorder)<br>Craving ↓ (iTBS in patients with methamphetamine use disorder)<br>Improved executive function, particularly working memory and inhibition, was identified as a potential predictor of therapeutic efficacy. |
| Zangen et al., (2021)<br>312 | Nicotine | 123 | Pre-treatment treatment seeker | 15+3 | 3 months | Sham controlled | Parallel | 6 cm rule | right DLPFC-HF TMS/left DLPFC-HF TMS/right Insula-HF TMS/left Insula-HF TMS | H4 coil | 10 | 120 | 1800 | Abstinence ↑<br>Craving ↓<br>Consumption ↓ |
| Ankit et al.,(2022)<br>173 | Opioids | 20 | Detoxification | 14 | NA | Sham controlled and another therapy | Parallel | EEG System | right FPC-LF TMS | Figure of 8 | 50 | 80 | 900 | Craving ↓ (Both groups)<br>Serum brain-derived neurotrophic factor (BDNF) ↓ (Both groups)<br>No significant difference observed between the two groups.<br>Significant correlation between serum BDNF levels over time and different clinical variables in the active group. |
| Belgers et al.,(2022)<br>174 | Alcohol | 14 | Detoxification | 10 | 12 months | Sham controlled and another therapy | Parallel | EEG System | right DLPFC-HF TMS | Figure of 8 | 10 | 110 | 3000 | Craving ↓<br>Alcohol use ↓<br>Differences in craving between groups were most prominent three months after treatment. |
| Bidzinski et al.,(2022)<br>175 | Cannabis | 9 | Pre-treatment treatment seeker | 20 | < 1 month | Sham controlled | Parallel | EEG System | right DLPFC-HF TMS/left DLPFC-HF TMS | Figure of 8 | 20 | 90 | 750 | Cannabis use ↓<br>Positive and Negative Syndrome Scale (PANSS) positive subscale ↓<br>Attention ↑<br>Tobacco use ↓ |
| Feng et al.,(2022)<br>176 | Alcohol | 36 | Pre-treatment treatment seeker | 14 | NA | Sham controlled | Parallel | EEG System | left DLPFC-HF TMS | Figure of 8 | 10 | 110 | 1530 | Response time in the oddball task ↓<br>The total score of MoCA ↑<br>The total score of BIS-II ↓ |
| Harel et al.,(2022)<br>177 | Alcohol | 23 | Pre-treatment treatment seeker | 15+5 | 3 months | Sham controlled | Parallel | 4 cm rule | right FPC-HF TMS/left FPC-HF TMS/ACC-HF TMS | H7 coil | 10 | 100 | 3000 | Craving ↓<br>Alcohol consumption ↓<br>Functional connectivity of the dorsal anterior cingulate cortex with the caudate nucleus ↓<br>Connectivity of the medial prefrontal cortex to the subgenual anterior cingulate cortex ↓ |
| Jin et al.,(2022)<br>178 | Opioids | 17 | Recovery | 7 | NA | Sham controlled and healthy control and another therapy | Parallel | 6 cm rule | left DLPFC-HF TMS | Figure of 8 | 10 | 100 | 2000 | Spontaneous and heroin cue-induced craving ↓<br>The coupling between the left DLPFC and left parahippocampal gyrus ↑<br>The coupling between the right precentral gyrus and three key regions included in DMN ↓<br>the left DLPFC-PHG coupling negatively correlated with the spontaneous craving and the drug cue-induced craving. |
| Liu, Sun, et al.,(2022)<br>179 | Amphetamine | 20 | Pre-treatment treatment seeker | 12 | NA | Active controlled | Parallel | 5 cm rule | left DLPFC-HF TMS/left DLPFC-HF TMS | Round coil | 50/10 | 80/100 | 600/2000 | Cue-induced craving ↓ (in both groups)<br>Withdrawal symptoms ↓ (in both groups) |
| Marques et al.,(2022) | Nicotine | 24 | Active user non-treatment seeker | 1 | NA | Active controlled | Parallel | EEG System | left FPC-LF TMS | Figure of 8 | 1 | 110 | 1200 | Cue-reactivity ↓ (left frontal pole group)<br>Reactivity to affectively neutral pictures ↓ (left frontal pole group) |
| Martinotti et al.,(2022)<br><sup>281</sup> | Cocaine | 36 | Pre-treatment treatment seeker | 20+24 | 3 months | Sham controlled | Parallel | Beam F3/MRI guided | left DLPFC-HF TMS | Figure of 8 | 15 | 100 | 2400 | Cue-induced craving ↓ (in both groups)<br>Consumption ↓ (in both groups)<br>Depressive symptoms ↓ (active group) |
| McNeill et al.,(2022)<br><sup>282</sup> | Alcohol | 60 | Active user non-treatment seeker | 1 | NA | Active controlled | Parallel | EEG System | right DLPFC-LF TMS/left DLPFC-LF TMS/medial FPC-LF TMS | Figure of 8 | 50 | 80 | 600 | Inhibitory control ↑ (in the right DLPFC group)<br>Craving and consumption ↑ (in the left DLPFC group) |
| Mikellides et al.,(2022)<br><sup>283</sup> | Nicotine | 59 | Pre-treatment treatment seeker | 20 | < 1 month | Active controlled and sham controlled | Parallel | Beam F3 | left DLPFC-HF TMS | Figure of 8 | 50 | 100 | 600 | Cigarette consumption, nicotine dependence, craving, and perceived stress ↓ (in all groups) |
| Moeller et al.,(2022)<br><sup>284</sup> | Nicotine | 10 | Pre-treatment treatment seeker | 15 | NA | Sham controlled | Parallel | 6 cm rule | right Insula-HF TMS/left Insula-HF TMS/right DLPFC-HF TMS/left DLPFC-HF TMS | H4 coil | 10 | 120 | 1800 | Bilateral dTMS disrupted smoking behavior<br>Active dTMS showing delayed initiation of smoking<br>Insula blood flow ↓<br>Connectivity between the insula and default mode network ↑ |
| Shevorykin et al.,(2022)<br><sup>285</sup> | Nicotine | 23 | Pre-treatment treatment seeker | 8/12/16/24/32 | 6 months | Active controlled | Parallel | MRI guided | left DLPFC-HF TMS | Figure of 8 | 20 | 110 | 900 | The duration and intensity of rTMS ↑ smoking cessation outcomes ↑<br>Delay discounting emerged as a promising therapeutic target<br>Participant burden ↓ even with extended treatment protocols |
| Su et al.,(2022)<br><sup>286</sup> | Amphetamine | 20 | Pre-treatment treatment seeker | 20 | NA | Sham controlled and healthy control | Parallel | Beam F3 | left DLPFC-HF TMS | Figure of 8 | 50 | 100 | 900 | rTMS intervention reversed the expression of three key metabolites (alpha-tocopherol, glyceric acid, and fumaric acid) and improvements in alpha-tocopherol levels were associated with enhanced cognitive function. |
| Zhang et al.,(2022)<br><sup>287</sup> | Alcohol | 30 | Active user non-treatment seeker | 10 | 2 months | Sham controlled | Parallel | 6 cm rule | left DLPFC-HF TMS | Figure of 8 | 20 | 110 | 2000 | Days of heavy drinking ↓<br>Craving ↓<br>Serum neurofilament light chain levels ↓<br>Social function and mental health ↑ |
| Li et al.,(2022)<br><sup>288</sup> | Nicotine | 3 | Pre-treatment treatment seeker | 5 | 1 month | Sham controlled | Parallel | 5.5 cm rule | left DLPFC-HF TMS | Figure of 8 | 10 | 100 | 3000 | Consumption ↓<br>Craving ↓ |
| Liu, Xu, et al.,(2022)<br>189 | Ampheta<br>mines | 29 | Pre-treatment<br>treatment<br>seeker | 20 | NA | Healthy control | Single arm | 5 cm<br>rule | left DLPFC-HF<br>TMS | Round<br>coil | 10 | 100 | 2000 | Craving ↓<br>Stop-signal reaction<br>time ↓ |
| Hu et al.,(2022)<br>190 | Alcohol | 125 | Pre-treatment<br>treatment<br>seeker | 10 | 6<br>mo<br>nth<br>s | Active<br>controlled and<br>sham<br>controlled and<br>another<br>therapy | Parallel | EEG<br>System | left DLPFC-HF<br>TMS/right<br>DLPFC-HF TMS | Figure<br>of 8 | 10 | 110 | 1500 | Relapse rate ↓<br>Depression ↓ |
| Johnstone et al.,(2022)<br>191 | Cannabis | 9 | Pre-treatment<br>treatment<br>seeker | 20 | < 1<br>mo<br>nth | Sham<br>controlled | Parallel | EEG<br>System | right DLPFC-HF<br>TMS/left<br>DLPFC-HF TMS | Figure<br>of 8 | 20 | 90 | 750 | Consumption ↓<br>There is a<br>moderating role of<br>sustained attention,<br>delayed discounting,<br>and complex planning<br>on the effects of rTMS<br>on cannabis use. |
| Kang et al.,(2022)<br>192 | Opioids | 22 | Pre-treatment<br>treatment<br>seeker | 30 | NA | Sham<br>controlled and<br>healthy control | Parallel | 5 cm<br>rule | left DLPFC-HF<br>TMS | Figure<br>of 8 | 50 | 80 | 600 | Two-choice oddball<br>task performance ↑<br>Craving ↓ |
| Lechner et al.,(2022)<br>193 | Nicotine | 21 | Active user<br>non-treatment<br>seeker | 10 | NA | Sham<br>controlled and<br>another<br>therapy | Parallel | EEG<br>System | left DLPFC-HF<br>TMS | Figure<br>of 8 | 10 | 100 | 2000 | Letter-sequencing<br>performance ↑<br>(rTMS group)<br>Back-digit task<br>performance ↑<br>(rTMS group)<br>Performance on the<br>smoking analog task<br>↓ (rTMS+WMT)<br>Back-digit task<br>performance ↓<br>(rTMS+WMT) |
| Sun et al.,(2022)<br>194 | Ampheta<br>mines | 9 | Pre-treatment<br>treatment<br>seeker | 5 | NA | Sham<br>controlled | Parallel | EEG<br>System | left DLPFC-HF<br>TMS | Figure<br>of 8 | 50 | 80 | 600 | Working memory<br>performance ↑ (after<br>one session)<br>Working memory<br>capacity ↑ |
| Wang et al.,(2022)<br>195 | Ampheta<br>mines | 30 | Pre-treatment<br>treatment<br>seeker | 12 | NA | Sham<br>controlled | Parallel | EEG<br>System | left DLPFC-HF<br>TMS | Figure<br>of 8 | 10 | 100 | 400 | Craving ↓<br>Adverse selection<br>and intensity of<br>impulsive decision-<br>making ↓ |
| McCauley et al.,(2023)<br>196 | Alcohol | 26 | Pre-treatment<br>treatment<br>seeker | 10 | 3<br>mo<br>nth<br>s | Sham<br>controlled | Parallel | EEG<br>System | left FPC-LF TMS | Figure<br>of 8 | 50 | 110 | 3600 | Individuals who<br>received real TBS<br>were significantly<br>more likely to remain<br>enrolled in the study<br>and maintain sobriety<br>three months after<br>treatment initiation.<br>Additionally, real TBS<br>led to a notable<br>reduction in brain<br>reactivity to alcohol<br>cues, particularly<br>decreasing MPFC-striatum and MPFC-insula connectivity at<br>2- and 3-months post-<br>treatment. |
| Mikellides et al., (2023)<br>237 | Nicotine | 59 | Pre-treatment treatment seeker | 20 | 6 months | Active controlled and sham controlled | Parallel | Beam F3 | left DLPFC-HF TMS | Figure of 8 | 50 | 100 | 600 | Cigarette consumption, nicotine dependence, craving and perceived stress ↓ (in all groups) |
| Jin et al., (2023)<br>238 | Opioids | 30 | Recovery | 7 | NA | Healthy control | Single arm | 6 cm rule | left DLPFC-HF TMS | Figure of 8 | 10 | 100 | 2000 | Craving ↓<br>Withdrawal ↓<br>The left IPL-left middle frontal gyrus coupling and the left IPL-right inferior occipital gyrus coupling ↓ |
| Hoven et al., (2023)<br>239 | Alcohol | 40 | Recovery | 10 | 6 months | Sham controlled and another therapy | Parallel | EEG System | right DLPFC-HF TMS | Figure of 8 | 10 | 110 | 3000 | HF-rTMS had no effect on the number of abstinent days over a six-month period. Additionally, HF-rTMS did not influence craving levels. |
| Dong et al., (2023)<br>240 | Opioids/Amphetamines | 38 | Pre-treatment treatment seeker | 20 | NA | Active controlled and sham controlled | Parallel | 5 cm rule | left DLPFC-HF TMS | Figure of 8 | 10/50 | 100 | 900 | Memory, attention, social cognition ↑ (in iTBS)<br>Serum EPI and GABA-Aα5 ↓ (iTBS) |
| Zhang et al., (2023)<br>241 | Amphetamines | 18 | Recovery | 20 | NA | Sham controlled | Parallel | EEG System | left DLPFC-HF TMS | Round coil | 50 | 80 | 600 | Working memory accuracy and discrimination ability ↑<br>Reaction time ↓<br>Resting-state delta power ↓ |
| Gersner et al., (2022)<br>241 | Nicotine | 75 | Pre-treatment treatment seeker | 15+3 | 3 months | Sham controlled | Parallel | 6 cm rule | right DLPFC-HF TMS/left DLPFC-HF TMS/right Insula-HF TMS/left Insula-HF TMS | H4 coil | 10 | 120 | 1800 | Three weeks of Deep TMS has a higher smoking addiction quit rate in participants who are younger, more educated, Caucasian, and with less extensive smoking history. |
| Shevorykin et al., (2023)<br>243 | Nicotine | 51 | Pre-treatment treatment seeker | 8/12/16/24/32 | 6 months | Active controlled and sham controlled | Parallel | EEG System | left DLPFC-HF TMS | Figure of 8 | 20 | 110 | 900 | The overall research burden of rTMS treatment was relatively low, with participants reporting significantly lower experienced burden compared to anticipated burden. Additionally, neither the duration or intensity of rTMS nor race or income significantly influenced perceived research burden. |
| S. Upton et al., (2023)<br>244 | Nicotine | 37 | Active user non-treatment seeker | 1 | NA | Active controlled | Crossover | MRI guided | right IFG-HF TMS/right IFG-LF TMS | Figure of 8 | 50 | 80 | 600 | Inhibitory control task performance ↑ (in cTBS)<br>Craving and smoking ↓ (both iTBS and cTBS)<br>cTBS-induced improvement in IC task performance was associated with reduced craving.<br>cTBS-induced reduction in craving was associated with reduced smoking. |
| Ibrahim et al.,(2023)<br>205 | Nicotine | 24 | Active user non-treatment seeker | 20 | 2 months | Sham controlled and another therapy | Parallel | NA | right Insula-HF TMS/left Insula-HF TMS | H11 coil | 10 | 120 | 1020 | Abstinence rates ↑ |
| Gong et al.,(2023)<br>206 | Opioids | 38 | Recovery | 20 | < 1 month | Active controlled and sham controlled and another therapy | Parallel | NA | left DLPFC-HF TMS | NA | 50 | 100 | 900 | Anxiety, depression, and craving ↓ |
| Gerace et al.,(2023)<br>207 | Cocaine | 58 | Pre-treatment treatment seeker | 15 | 2 months | Sham controlled and healthy control | Parallel | 5 cm rule | left DLPFC-HF TMS | Figure of 8 | 15 | 100 | 2400 | CUD patients exhibited reduced alpha diversity and notable alterations in gut and oral microbiota composition and function, including lower levels of butyric acid. After rTMS treatment, these microbiota changes were partially reversed, with butyric acid levels restored. |
| Spencer Upton et al.,(2023)<br>208 | Nicotine | 31 | Active user non-treatment seeker | 1 | NA | Active controlled | Crossover | MRI guided | right IFG-HF TMS/right IFG-LF TMS | Figure of 8 | 50 | 80 | 600 | Craving ↓ (in cTBS)<br>Consumption ↓ (in cTBS)<br>Resting-state functional connectivity between the rIFG pars opercularis and subcallosal cingulate (fronto-striatal circuit), and between the rIFG pars opercularis and the right posterior parahippocampal gyrus ↑ (in cTBS) |
| Ding et al.,(2023)<br>209 | Opioids | 32 | Detoxification | 30 | NA | Healthy control | Single arm | 5 cm rule | left DLPFC-HF TMS | Figure of 8 | 50 | 80 | 600 | Resting-state EEG analysis identified four microstate classes (A-D), with significant increases in the duration, occurrence, and contribution of microstate class A following iTBS treatment. Machine learning achieved 81.5% classification accuracy, highlighting EEG microstates as reliable indicators of treatment effects in patients with heroin addiction. |

**Abbreviations:** TMS: transcranial magnetic stimulation, HF TMS: high-frequency TMS ( $\geq 5$ Hz), LF TMS: low-frequency TMS ( $\leq 1$ Hz), rTMS: repetitive TMS, dTMS: deep TMS, iTBS: intermittent theta burst stimulation, cTBS: continuous theta burst stimulation, EEG: electroencephalography, MRI: magnetic resonance imaging, DLPFC: dorsolateral prefrontal cortex, ACC: anterior cingulate cortex, PCC: posterior cingulate cortex, FPC: frontopolar cortex, IFG: inferior frontal gyrus, SFG: superior frontal gyrus, NA: not available.

### Stimulation targets

**Figure S.1.**
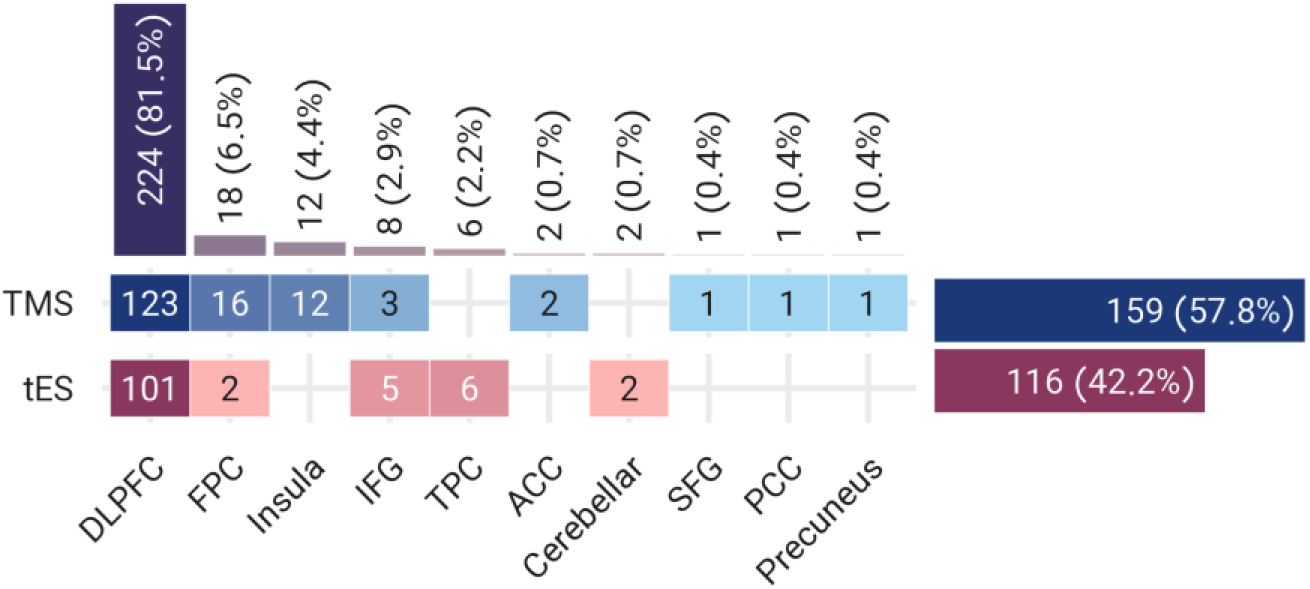
Variations in Targets in tES/TMS Studies for Substance Use Disorders. This figure displays the number of studies tackling specific target areas. The studies are color-coded, with tES in red and TMS in blue. Note that some studies utilize multiple stimulation protocols, resulting in 275 total targets in this figure, exceeding the actual number of studies included. <u>Abbreviations</u>: tES: transcranial electrical stimulation, TMS: transcranial magnetic stimulation, DLPFC: dorsolateral prefrontal cortex, FPC: frontopolar cortex, IFG: inferior frontal gyrus, TPC: temporoparietal cortex, ACC: anterior cingulate cortex, SFG: superior frontal gyrus, PCC: posterior cingulate cortex.

### Electrode/coil positioning system

Of the 121 TMS studies reviewed, the majority focused on conventional repetitive transcranial magnetic stimulation (rTMS) (n=75). The remaining studies utilized intermittent theta burst stimulation (iTBS) (n=16), continuous theta burst stimulation (cTBS) (n=8), deep transcranial magnetic stimulation (dTMS) (n=13), or combinations of different TMS types, such as cTBS and iTBS (n=9). Among the 86 tES studies analyzed, the vast majority utilized transcranial direct current stimulation (tDCS) (n=83), while only three studies employed transcranial alternating current stimulation (tACS) (n=3). Among the tDCS studies, only one study utilized 4x1 high-definition (HD) electrodes (n=1), whereas the remaining tES studies employed large conventional electrode pads (n=78). For 7 tDCS studies, the type of electrodes used was not reported.

**Figure S.2.**
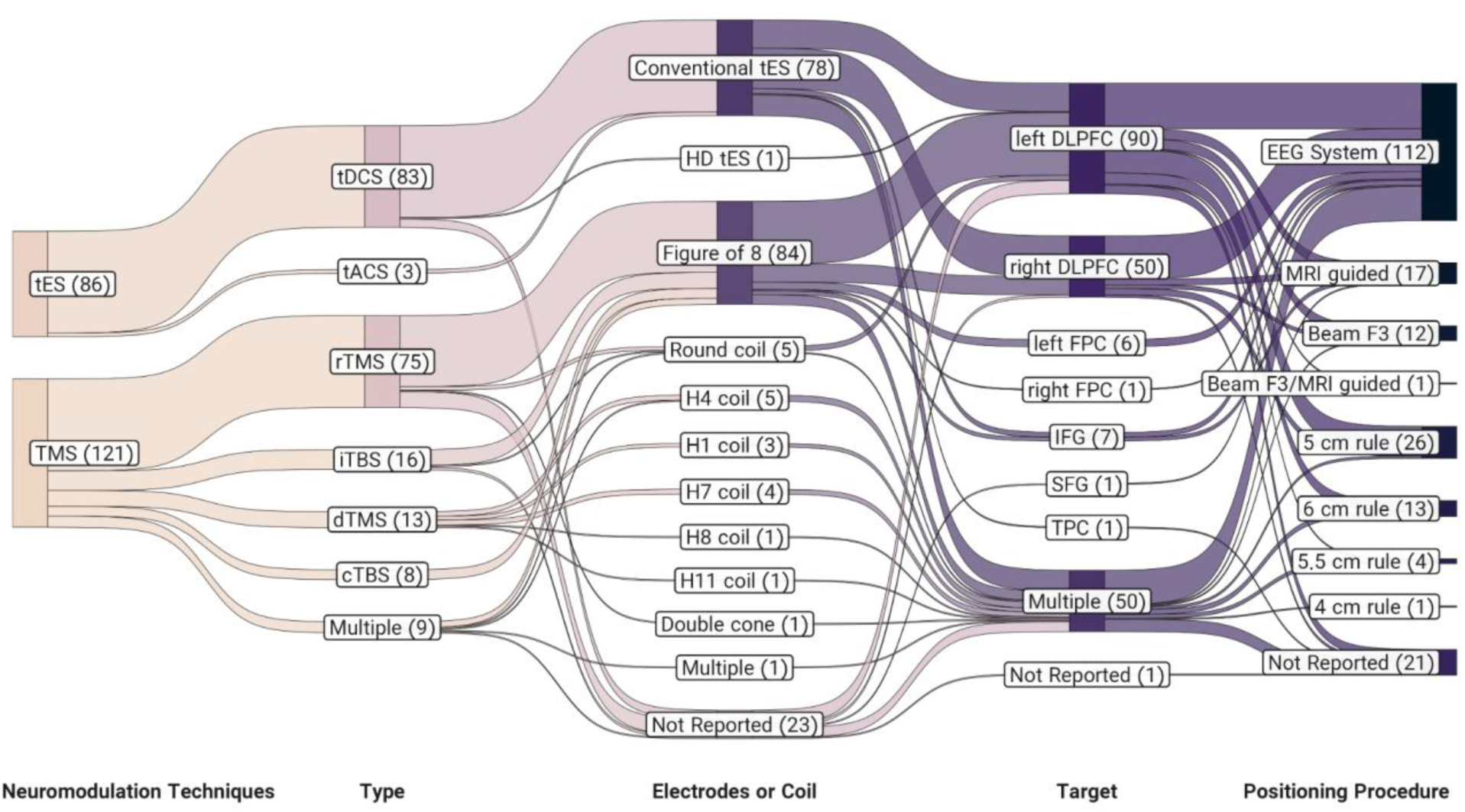
Types of Stimulation, Coils/Electrodes, Targets, and Positioning Systems in tES/TMS Studies for Substance Use Disorders. This Sankey diagram depicts the distribution of 207 tES/TMS studies, categorized by the type of stimulation, coil/electrode, target area, and positioning system used. The number of studies is indicated in parentheses within the boxes. Note: ’Multiple’ in the ’Type’ layer refers to studies that employed a combination of two different types of stimulation (e.g., cTBS and iTBS). ’Multiple’ in the ’Electrodes or Coil’ layer refers to studies that utilized a combination of two different coil shapes (e.g., deep and figure 8). ’Multiple’ in the ’Target’ layer refers to studies that employed a combination of different target areas. <u>Abbreviations</u>: TMS: transcranial magnetic stimulation, tES: transcranial electrical stimulation, rTMS: repetitive TMS, dTMS: deep TMS, iTBS: intermittent theta burst stimulation, cTBS: continuous theta burst stimulation, tDCS: transcranial direct current stimulation, tACS: transcranial alternating current stimulation, EEG: electroencephalography, MRI: magnetic resonance imaging, DLPFC: dorsolateral prefrontal cortex, FPC: frontopolar cortex, IFG: inferior frontal gyrus, TPC: temporoparietal cortex, SFG: superior frontal gyrus.

In total, 207 TMS/tES studies were included in the analysis, employing various methods for positioning tES electrodes or TMS coils over the scalp. The most commonly utilized method was based on the EEG standard system (n=112). Additionally, other methods such as MRI-guided techniques (n=17) and Beam F3 methods (n=12) were used. Some studies utilized scalp measurements and applied different rules for electrode or coil placement using anatomical landmarks, including the 5 cm rule (n=26), 6 cm rule (n=13), 5.5 cm rule (n=4), and 4 cm rule (n=1). For 21 studies, the positioning system used was not reported.

### Outcome measures

Each study included in our systematic review reported a diverse range of primary and secondary outcome measures. To organize and categorize these measures, we extracted data from the full text of the studies and identified seven main categories. We recorded both significant and non-significant results reported in the abstract for each outcome measure (see Figure S.6.). The most commonly reported outcome measure across the majority of studies was "craving" (n=159), with 84 significant reductions and 39 non-significant changes in response to TMS/tES reported in the abstracts. Other frequently reported outcome measures included "consumption/relapse/abstinence" (n=111), "cognitive functions" (n=95), "physiological measures" (n=76), "negative valence" (n=59), "mental/general health" (n=33), and "positive valence" (n=8). Additionally, a few other outcome measures were reported, categorized as “other” (n=10). Across all studies, a total of 317 significant effects and 99 non-significant alterations in response to TMS/tES on the outcome measures were reported.

**Figure S.3.**
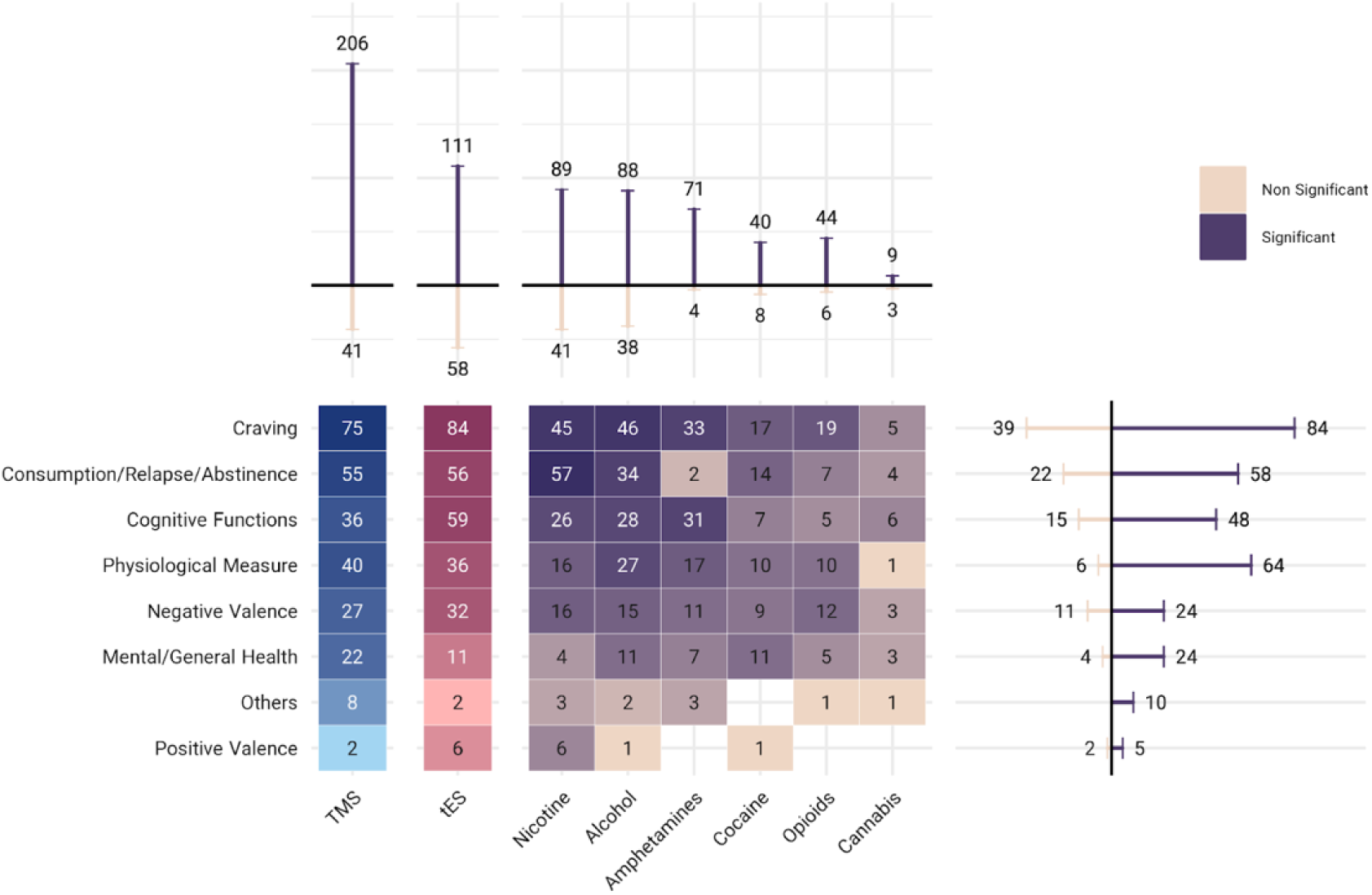
Reported Outcome Measures and Their Significance in tES/TMS Studies for Substance Use Disorders Considering Type of Substances. This figure illustrates the various outcome measures from past tES and TMS studies, color-coded by the type of stimulation—tES in red and TMS in blue—and categorized by the type of substance use disorder. Colors range from light yellow to purple, with darker shades indicating a higher number of publications. Note that some studies have included more than one type of substance and outcome measures, resulting in the total items in this figure equaling 551 and exceeding the actual number of 207 studies included. The number of significant and non-significant outcome measures is depicted in line bar graphs, which are color-coded by significance: dark colors indicate significant, and light colors indicate non-significant results. <u>Abbreviations</u>: tES: transcranial electrical stimulation, TMS: transcranial magnetic stimulation.

### Outcome measures based on stimulation targets

To examine the impact of the stimulation target, type of stimulation (excitatory or inhibitory), and coil/electrode positioning system on the reported results (significant or non-significant alterations), our analysis revealed noteworthy findings. Specifically, for TMS studies, the most significant responses were observed in high-frequency stimulation of either the left or right DLPFC. Left DLPFC stimulation yielded 153 significant and 32 non-significant responses, while right DLPFC high-frequency stimulation resulted in 50 significant and 14 non-significant responses. Similar patterns were identified in tES studies, where excitatory DLPFC stimulation proved to be the most effective protocol. Notably, anodal tDCS targeting the right DLPFC exhibited 61 significant and 25 non-significant responses, while anodal tDCS targeting the left DLPFC produced 53 significant and 28 non-significant alterations.

**Figure S.4-1.**
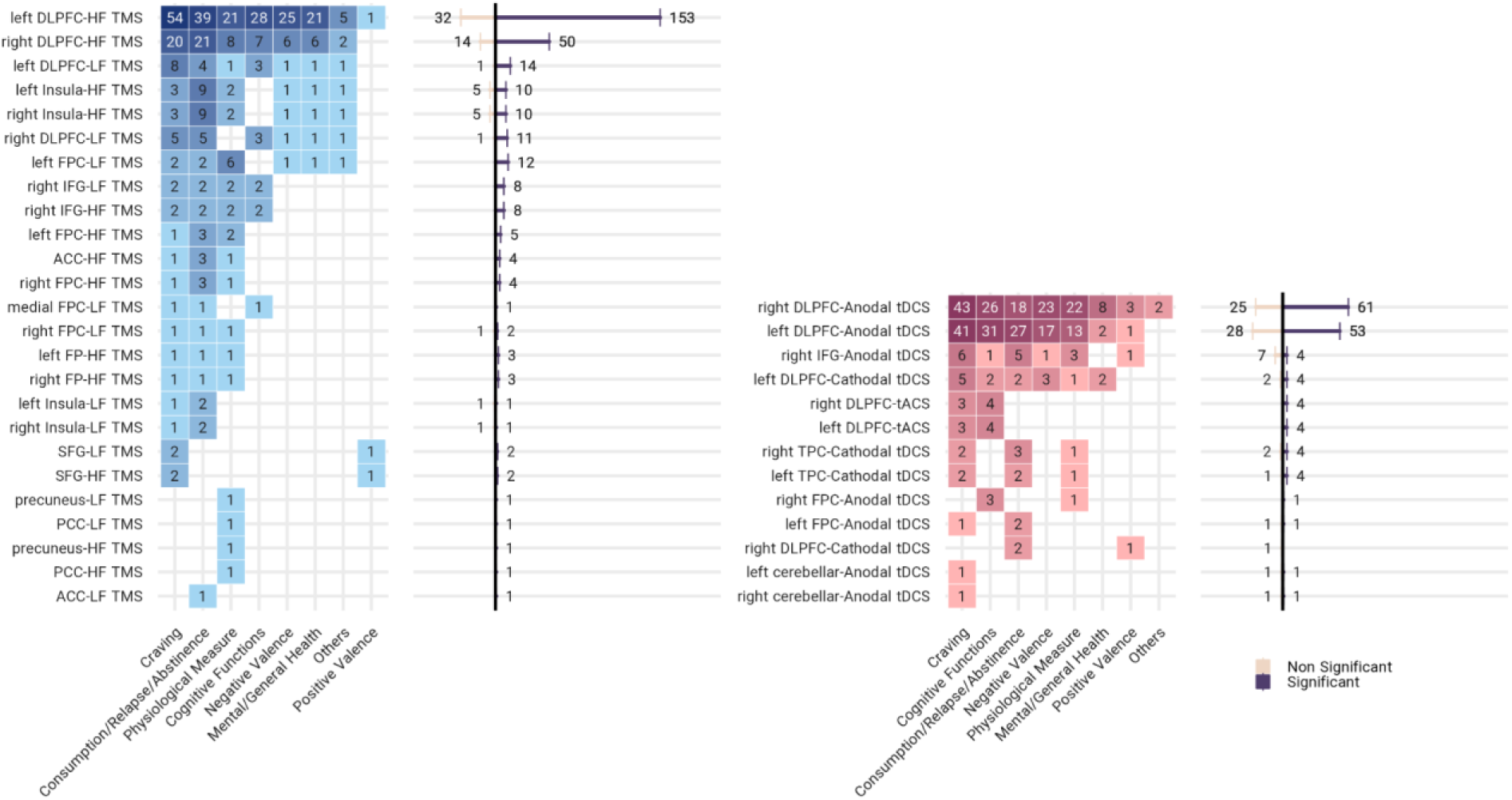
Reported Outcome Measures and Their Significance in tES/TMS Studies for Substance Use Disorders Considering Target Location and Stimulation Type. This figure illustrates the various outcome measures from 207 tES and TMS studies, color-coded by the type of stimulation—tES in red and TMS in blue—and categorized by the target location and type of stimulation (excitatory: anodal tES or HF TMS, inhibitory: cathodal tES or LF TMS). It is important to note that the total numbers reported for outcome measures do not add up to 207, as some studies reported multiple outcome measures. The number of significant and non-significant outcome measures is depicted in line bar graphs, color-coded by significance: dark colors indicate significant, and light colors indicate non-significant results. <u>Abbreviations</u>: tES: transcranial electrical stimulation, tDCS: transcranial direct current stimulation, tACS: transcranial alternating stimulation, TMS: transcranial magnetic stimulation, HF TMS: high-frequency TMS (≥ 5 Hz), LF TMS: low-frequency TMS (≤ 1 Hz), DLPFC: dorsolateral prefrontal cortex, FPC: frontopolar cortex, IFG: inferior frontal gyrus, TPC: temporoparietal cortex, ACC: anterior cingulate cortex, SFG: superior frontal gyrus, PCC: posterior cingulate cortex.

Considering that DLPFC stimulation emerged as the most frequently utilized target, yielding a substantial number of significant outcomes, we further explored the influence of electrode/coil positioning systems on the reported results. In TMS studies, several positioning methods demonstrated effectiveness in targeting the left DLPFC with high-frequency stimulation. The EEG standard system resulted in 39 significant and 9 non-significant responses, the 5 cm rule yielded 33 significant and 9 non-significant responses, MRI-guided techniques showed 20 significant and 1 non-significant responses, and the 6 cm rule resulted in 26 significant and 3 non-significant responses. In targeting the right DLPFC, the EEG standard system resulted in 15 significant and 7 non-significant responses. For tES studies, the EEG standard system proved to be the most effective positioning method for both right DLPFC (54 significant and 23 non-significant responses) and left DLPFC (44 significant and 24 non-significant responses) for anodal tDCS over DLPFC.

**Figure S.4-2.**
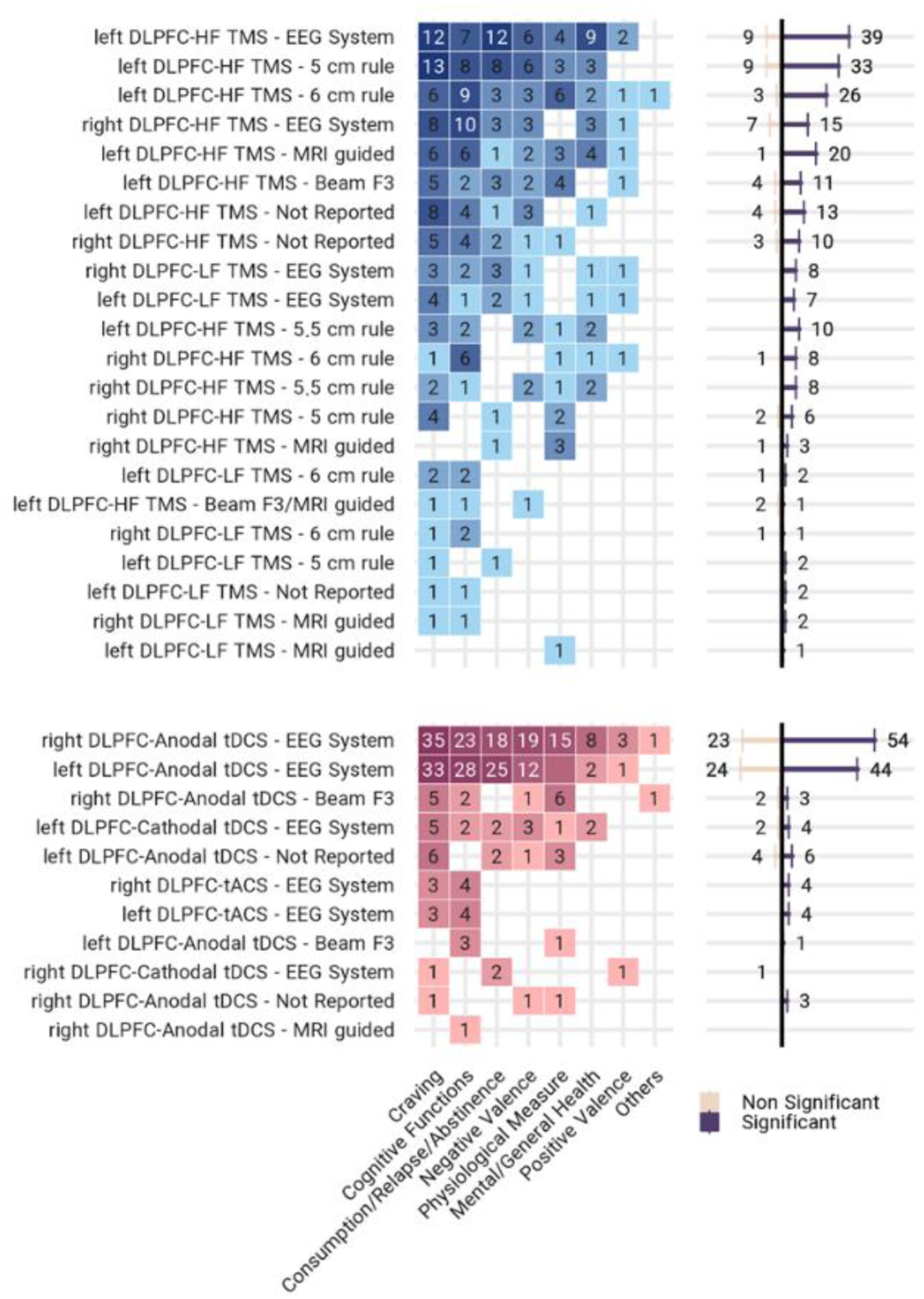
Relationship Between Stimulation Type, Positioning Method, and Significant Outcomes in DLPFC-Targeted tES/TMS Studies. This figure illustrates the various outcome measures from 207 tES and TMS studies that targeted DLPFC, color-coded by the type of stimulation—tES in red and TMS in blue—and categorized by the type of stimulation (excitatory: anodal tES or HF TMS, inhibitory: cathodal tES or LF TMS) and electrode/coil positioning system. The number of significant and non-significant outcome measures is depicted in line bar graphs, color-coded by significance: dark colors indicate significant, and light colors indicate non-significant results. In the square panels, studies are color-coded by the type of intervention: tES in red and TMS in blue and darker shades indicating a higher number of studies. <u>Abbreviations:</u> tES: transcranial electrical stimulation, tDCS: transcranial direct current stimulation, tACS: transcranial alternating stimulation, TMS: transcranial magnetic stimulation, HF TMS: high-frequency TMS (≥ 5 Hz), LF TMS: low-frequency TMS (≤ 1 Hz), DLPFC: dorsolateral prefrontal cortex.

### Study design and control condition

The study designs in our systematic review were categorized into two main types, parallel and crossover, and excluded all studies without a control condition. The most commonly utilized control condition across both TMS (n=62) and tES (n=48) studies was "sham control." However, other control methods were also employed, including active control, therapy other than TMS/tES, healthy control, other types of clinical populations, or combinations of different control conditions.

**Figure S.5.**
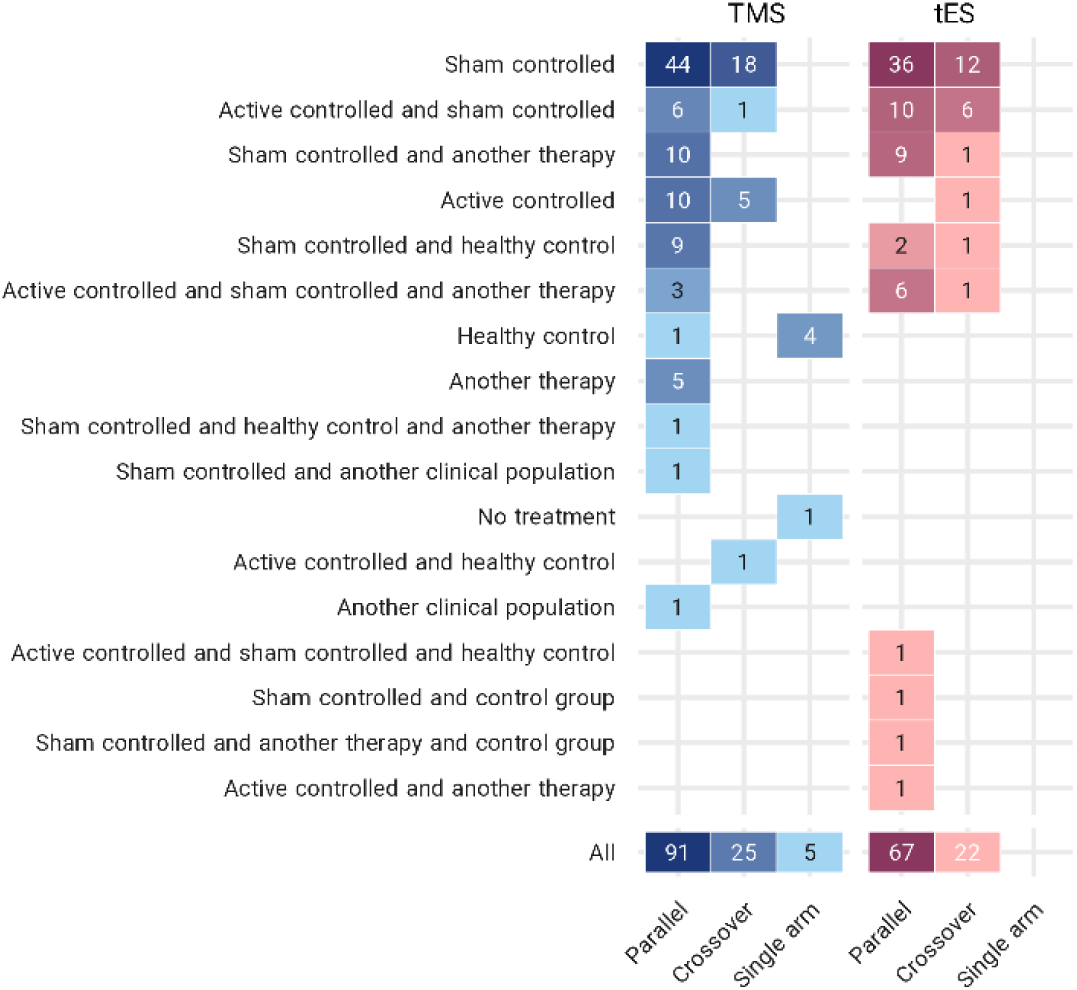
Variations in Study Design and Control Conditions in tES/TMS Studies for Substance Use Disorders. This figure presents the number of tES/TMS studies in substance use disorders, categorized by the type of randomization and divided by the control condition. The studies are color-coded according to the type of intervention, with tES in red and TMS in blue. <u>Abbreviations</u>: tES: transcranial electrical stimulation, TMS: transcranial magnetic stimulation.

### Dosage, number of sessions, and follow-up

Considering the stimulation dose, our analysis revealed the utilization of various pulse frequencies in the included TMS studies. The most commonly used pulse frequency was 10 Hz (n=57). Additionally, other stimulation frequencies were employed, including 50 Hz (n=39), 20 Hz (n=21), 1 Hz (n=12), 15 Hz (n=8), and 5 Hz (n=1) which can be assigned to excitatory (high-frequency TMS (≥ 5 Hz)), or inhibitory (low-frequency TMS (≤ 1 Hz)) stimulation. Among the tES studies, the dominant stimulation dose was found to be 2 mA (n=65). The second most frequently used dose was 1 mA, with 11 trials reporting its utilization. Additionally, 1.5 mA (n=8) and 0.45 mA (n=2) were employed in previous studies. Out of the 86 tES studies, 3 used tACS. Among the remaining 83 tDCS studies, 75 used anodal electrodes (excitatory), while 7 used cathodal electrodes (inhibitory) over their main brain target. One study employed both anodal and cathodal electrodes over their target across two different study arms. One TMS and one tES study did not report the stimulation frequency/intensity.

**Figure S.6-1.**
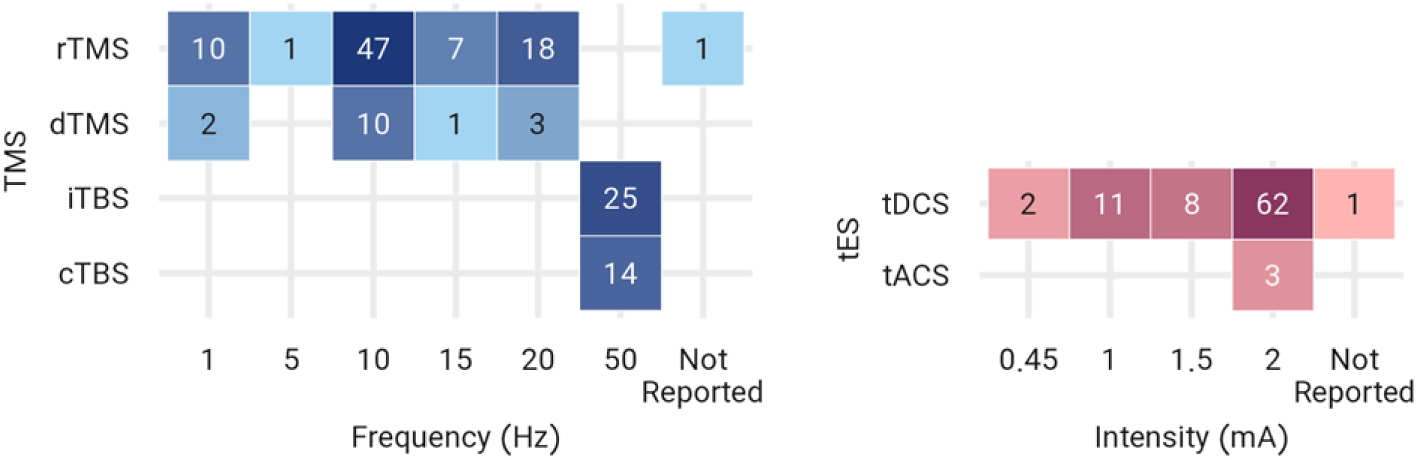
Variations in Stimulation Parameters in tES/TMS Studies for Substance Use Disorders. This figure displays the number of studies with different stimulation frequencies, which serve as an indicator of TMS dose, and electrical current intensity, which indicates the dose in tES studies (for tACS studies, the peak-to-peak current amplitude is considered). The studies are color-coded, with tES in red and TMS in blue. <u>Abbreviations</u>: tES: transcranial electrical stimulation; TMS: transcranial magnetic stimulation. In terms of the number of sessions included in the studies analyzed, we found that 30 TMS studies and 37 tES studies comprised single-session trials. For multi-session trials, 22 TMS and 15 tES studies applied 10 active stimulation sessions. 24 TMS and 2 tES studies applied 20 sessions of stimulation. 1 TMS trial applied 44 active stimulation sessions. Most of these studies had no follow-up, however, the most common duration for follow-up was less than one month (TMS: n=14, tES: n=10).

**Figure S.6-2.**
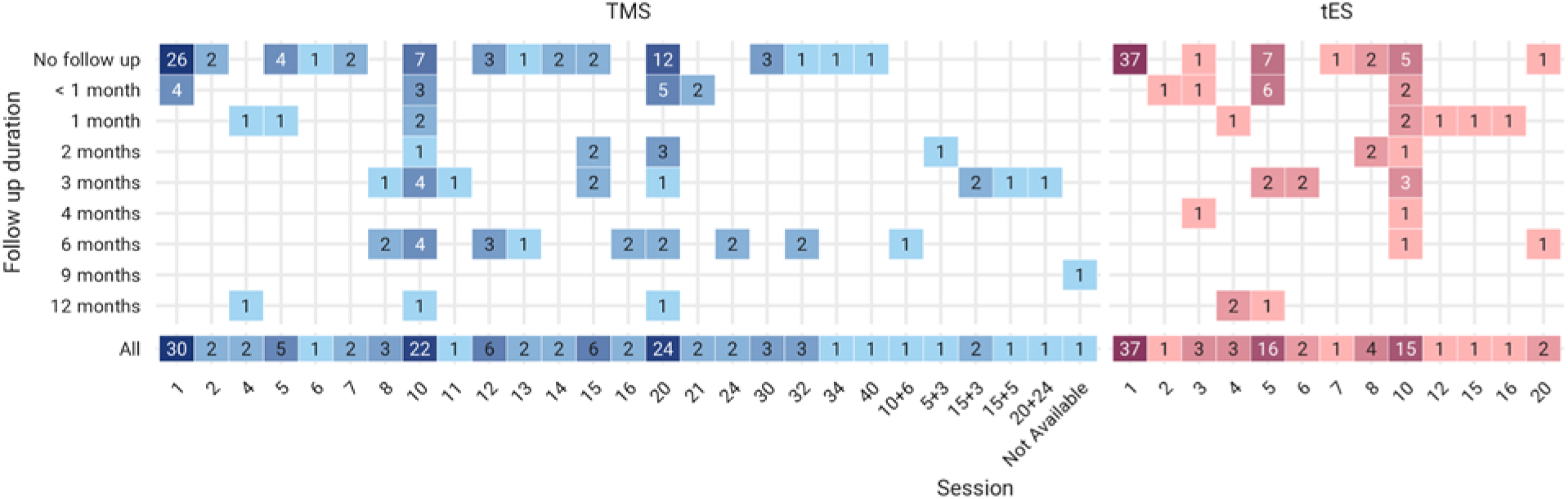
Variation in the Number of Sessions and Follow-Ups in tES/TMS Studies for Substance Use Disorders. This figure illustrates the follow-up durations, categorized by the number of stimulation sessions, with TMS represented in blue and tES in red. The follow-up durations are organized into 9 main groups to summarize the results succinctly. <u>Abbreviations:</u> tES: transcranial electrical stimulation; TMS: transcranial magnetic stimulation.

### Risk of bias

**Figure S.7-1.**
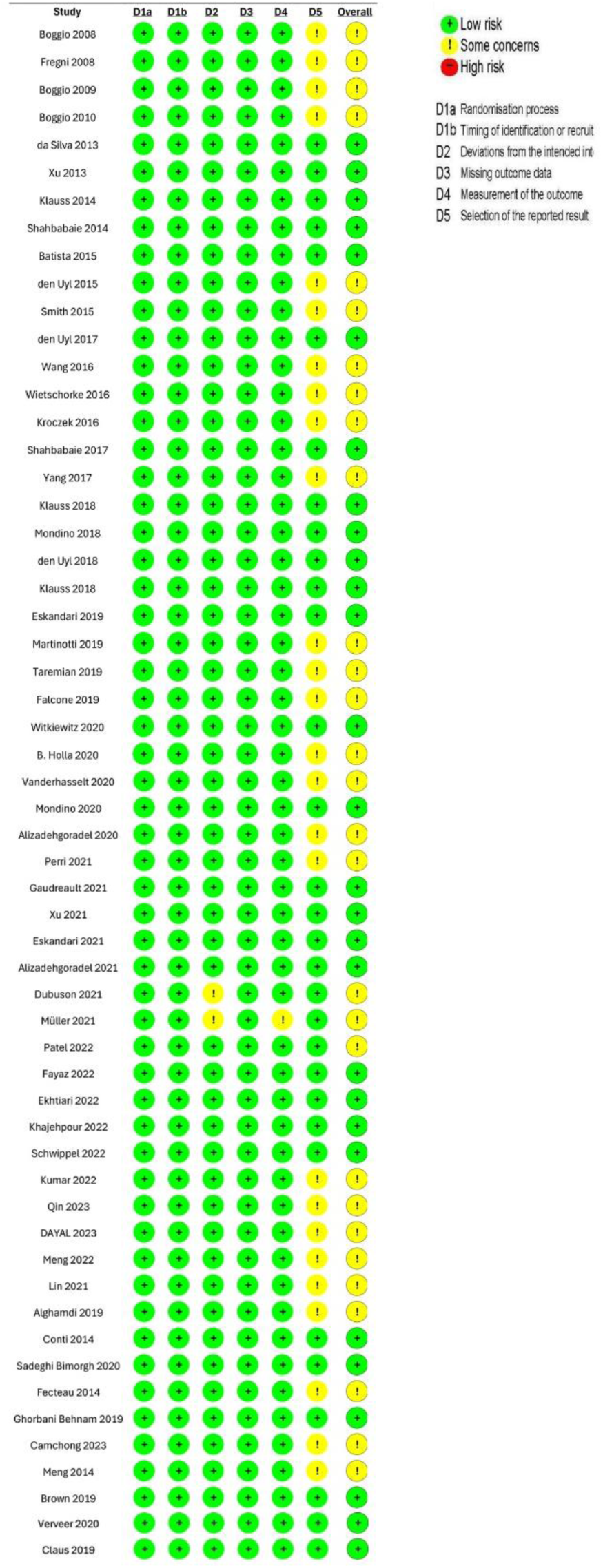
Risk of Bias of tES studies using the Cochrane Risk of Bias (RoB) 2 tool. The figures summarize the judgments across key domains of bias, including (1) bias due to the randomization process and timing of identification or recruitment of participants, (2) deviation from intended intervention, (3) missing outcome data, (4) measurement of outcomes, (5) selection of the reported result. Each domain is categorized as “Low risk”, “Some concerns”, or “High risk” bias. The overall judgment for risk of bias is shown in the last column.

**Figure S.7-2.**
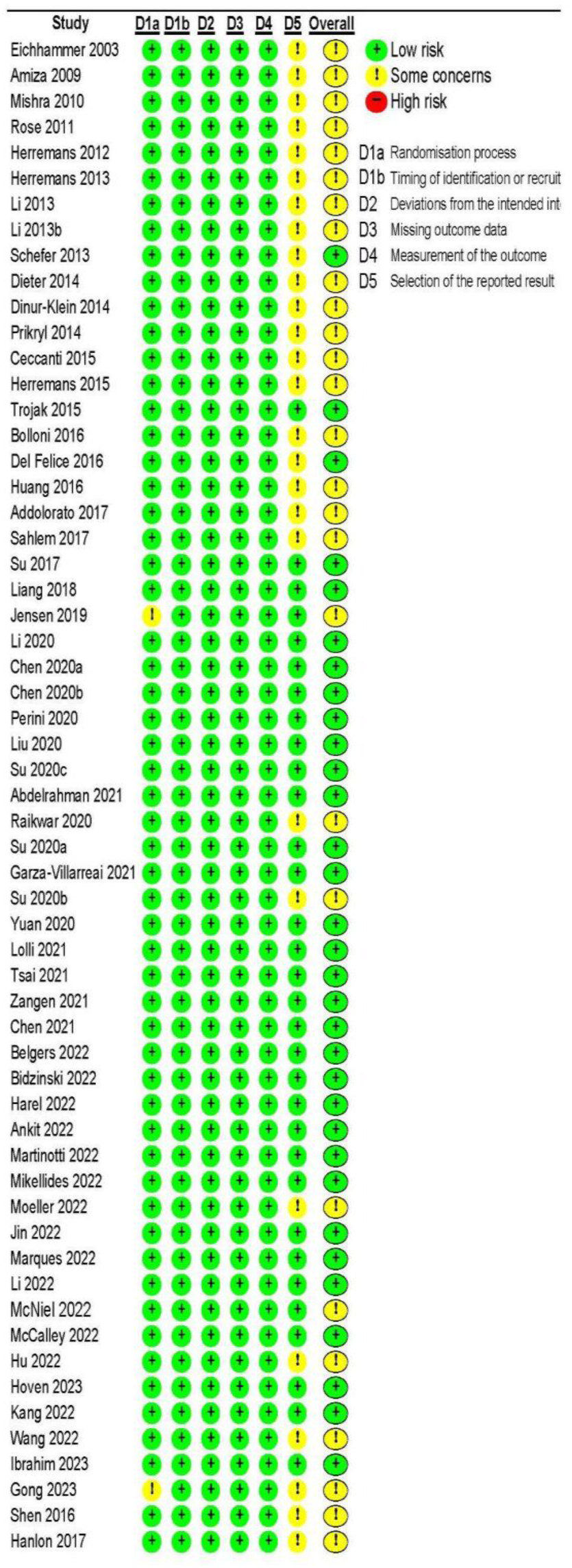
Risk of Bias of TMS studies using the Cochrane Risk of Bias (RoB) 2 tool. The figures summarize the judgments across key domains of bias, including (1) bias due to the randomization process and timing of identification or recruitment of participants, (2) deviation from intended intervention, (3) missing outcome data, (4) measurement of outcomes, (5) selection of the reported result. Each domain is categorized as “Low risk”, “Some concerns”, or “High risk” bias. The overall judgement for risk of bias is shown in the last column.

### Funnel plot

**Figure S.8-1.**
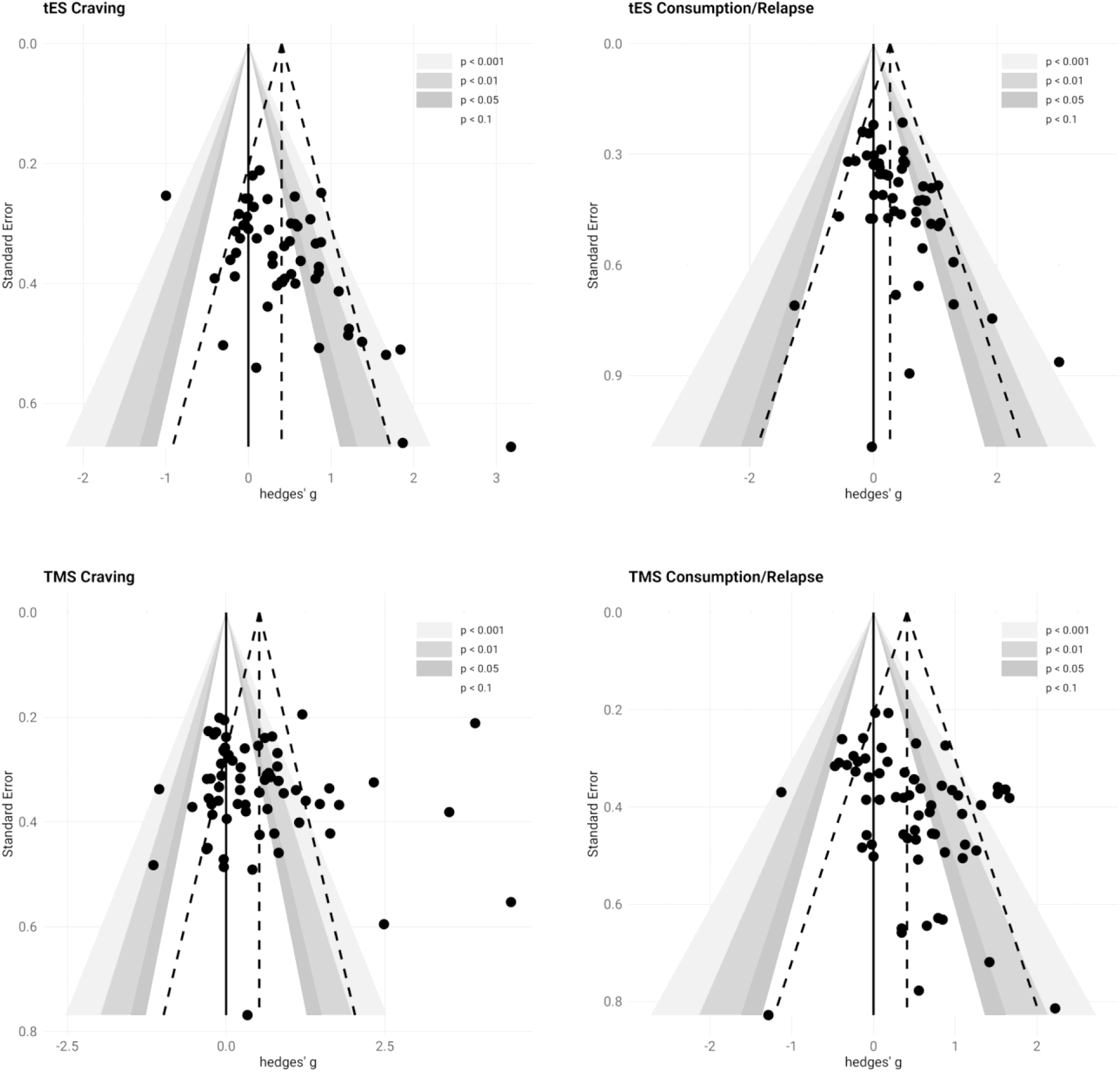
Funnel Plots for tES and TMS Studies on Craving and Consumption/Relapse. This figure presents funnel plots illustrating the heterogeneity across studies on craving and consumption/relapse for both tES and TMS protocols. Panel A (upper left) shows the funnel plot for tES studies on craving, while Panel B (upper right) displays the funnel plot for tES studies on consumption/relapse. Panel C (lower left) illustrates the funnel plot for TMS studies on craving, and Panel D (lower right) shows the funnel plot for TMS studies on consumption/relapse. In each plot, dots represent individual studies, with the y-axis representing study precision (standard error) and the x-axis representing the study’s estimated effect size (Hedges’ g). Larger studies with greater precision are shown at the top, while smaller, less precise studies scatter more widely at the bottom. <u>Abbreviations:</u> tES: transcranial electrical stimulation; TMS: transcranial magnetic stimulation.

### Forest plots

**Figure S.8-2.**
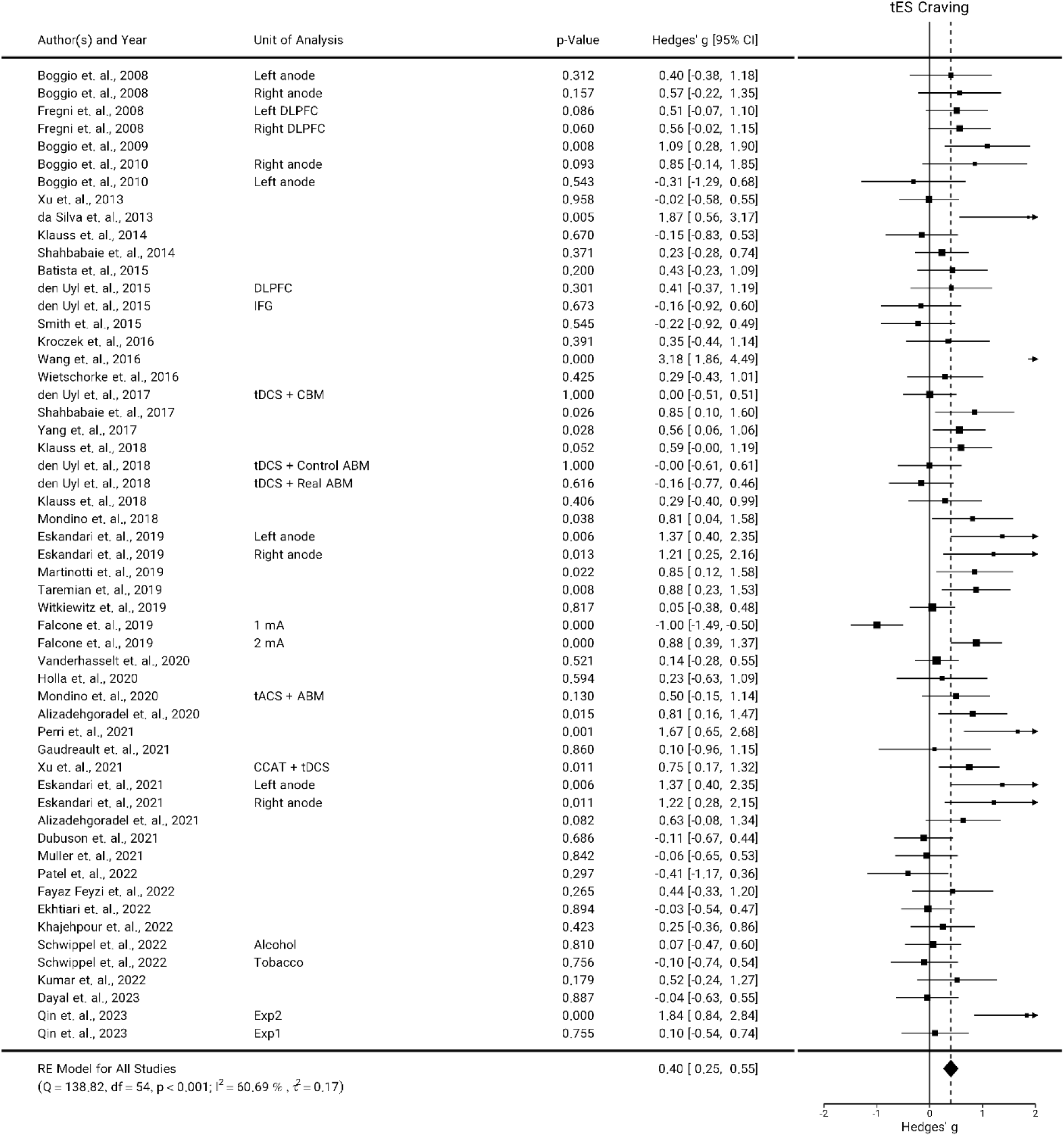
Forest plot for tES effects on Craving. Hedges’ g values for studies that reported craving in response to transcranial electrical stimulation (tES, with a total number of n=45 studies and 55 experiments). This forest plot provides a detailed analysis of the influence of tES on craving in various studies. Each horizontal line represents a specific study, with squares denoting effect sizes and horizontal lines indicating confidence intervals. Positive values indicate that active stimulation was effective in reducing craving. The diamond at the bottom illustrates the overall effect size across all studies, with the width representing the confidence interval. Units of analysis are added to studies with multiple experiments to highlight differences in study design. <u>Abbreviations</u>: tES: transcranial electrical stimulation, tDCS: transcranial direct current stimulation, tACS: transcranial alternating stimulation, DLPFC: dorsolateral prefrontal cortex, IFG: inferior frontal gyrus, CBM: cognitive bias modification, ABM: attentional bias modification, CCAT: computerized cognitive addiction therapy, Exp: experimental.

**Figure S.8-3.**
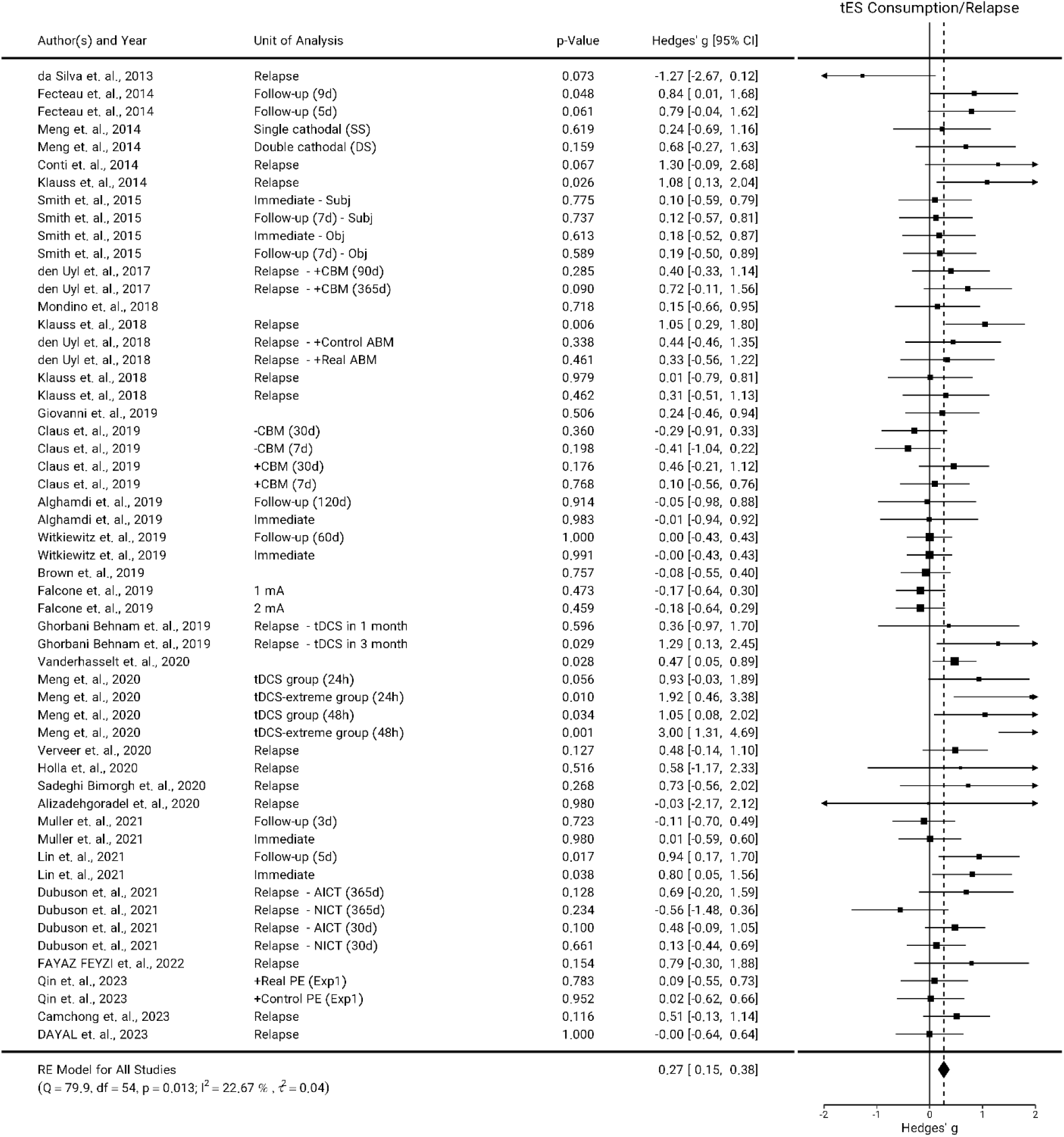
Forest plot for tES effects on Consumption/Relapse. Hedges’ g values for studies that reported consumption/relapse in response to transcranial electrical stimulation (tES, with a total number of n=31 studies and 55 experiments). This forest plot provides a detailed analysis of the influence of tES on consumption/relapse in various studies. Each horizontal line represents a specific study, with squares denoting effect sizes and horizontal lines indicating confidence intervals. Positive values indicate that active stimulation was effective in reducing consumption/relapse. The diamond at the bottom illustrates the overall effect size across all studies, with the width representing the confidence interval. Units of analysis are added to studies with multiple experiments to highlight differences in study design. <u>Abbreviations:</u> tES: transcranial electrical stimulation, tDCS: transcranial direct current stimulation, CBM: cognitive bias modification, ABM: attentional bias modification, AICT: alcohol cue inhibitory control training, NICT: neutral inhibitory control training, PE: psycho-education, Exp: experiment, mA: milliampere.

**Figure S.8-4.**
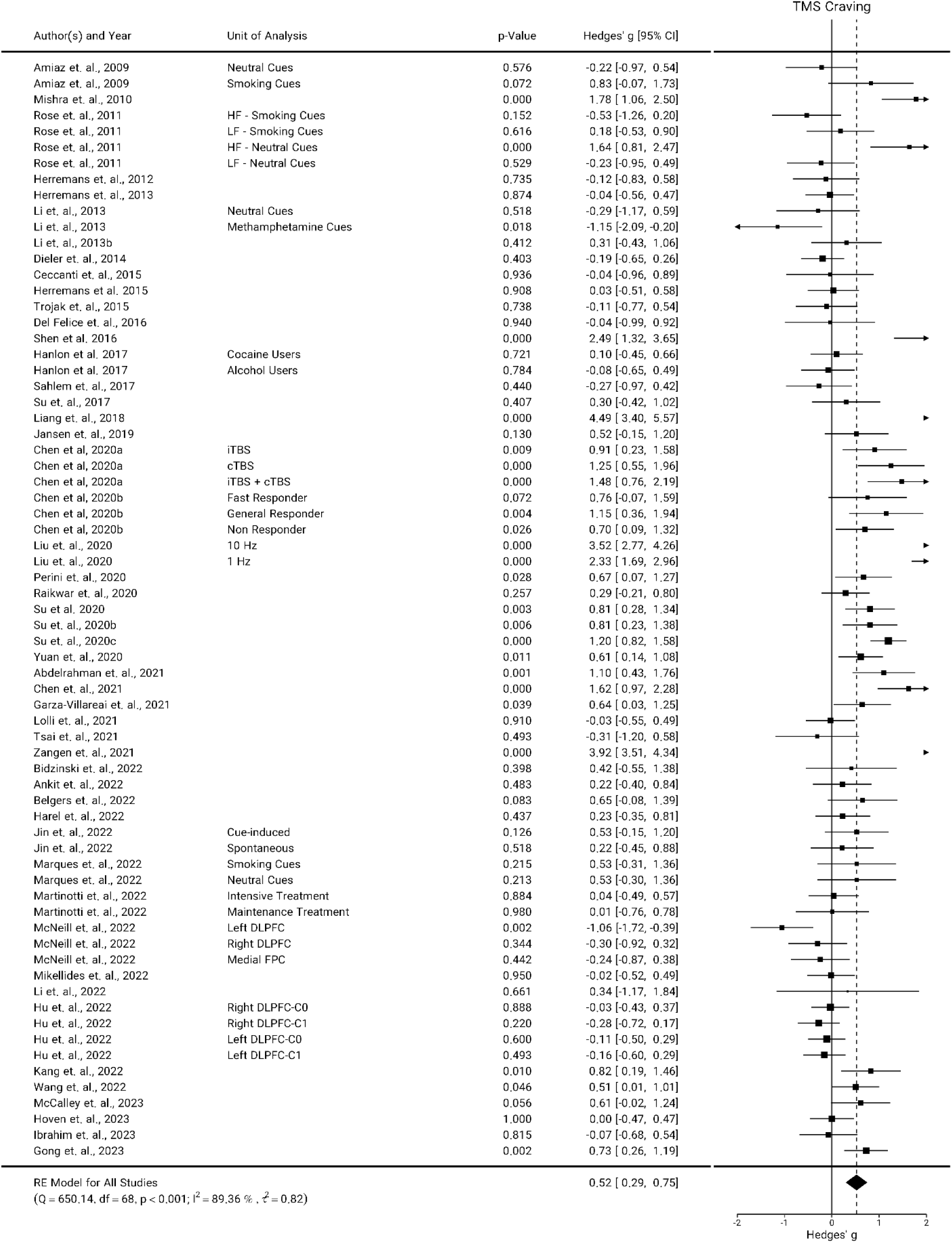
Forest plot for TMS effects on Craving. Hedges’ g values for studies that reported craving in response to transcranial magnetic stimulation (TMS, with a total number of n=50 studies and 69 experiments). This forest plot provides a detailed analysis of the influence of TMS on craving in various studies. Each horizontal line represents a specific study, with squares denoting effect sizes and horizontal lines indicating confidence intervals. Positive values indicate that active stimulation was effective in reducing craving. The diamond at the bottom illustrates the overall effect size across all studies, with the width representing the confidence interval. Units of analysis are added to studies with multiple experiments to highlight differences in study design. <u>Abbreviations</u>: TMS: transcranial magnetic stimulation, cTBS: continuous theta burst stimulation, iTBS: intermittent theta burst stimulation, HF TMS: high-frequency TMS (≥ 5 Hz), LF TMS: low-frequency TMS (≤ 1 Hz), DLPFC: dorsolateral prefrontal cortex, FPC: frontopolar cortex, MA: methamphetamine, C0: cognitive behavioral therapy treatment without fixed schedule, C1: cognitive behavioral therapy treatment with fixed schedule.

**Figure S.8-5.**
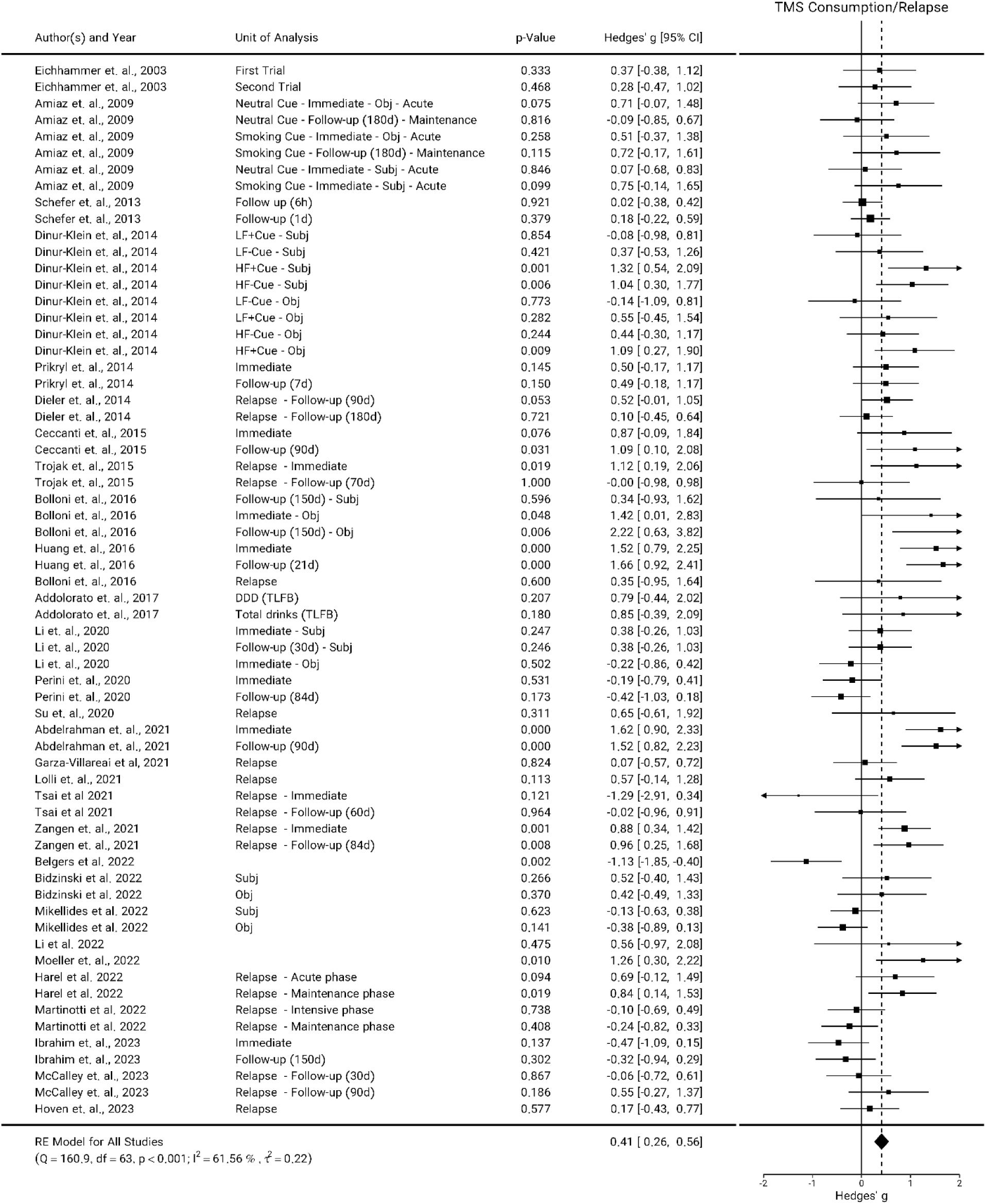
Forest plot for TMS effects on Consumption/Relapse. Hedges’ g values for studies that reported consumption/relapse in response to transcranial magnetic stimulation (TMS, with a total number of n=29 studies and 64 experiments). This forest plot provides a detailed analysis of the influence of TMS on consumption/relapse in various studies. Each horizontal line represents a specific study, with squares denoting effect sizes and horizontal lines indicating confidence intervals. Positive values indicate that active stimulation was effective in reducing consumption. The diamond at the bottom illustrates the overall effect size across all studies, with the width representing the confidence interval. Units of analysis are added to studies with multiple experiments to highlight differences in study design. <u>Abbreviations:</u> TMS: transcranial magnetic stimulation, HF TMS: high-frequency TMS (≥ 5 Hz), LF TMS: low-frequency TMS (≤ 1 Hz), d: day, Obj: objective, Subj: subjective, DDD: drinks per drinking day, TLFB: timeline follow back, d: day.

### Subgroup analysis

**Figure S.9-1.**
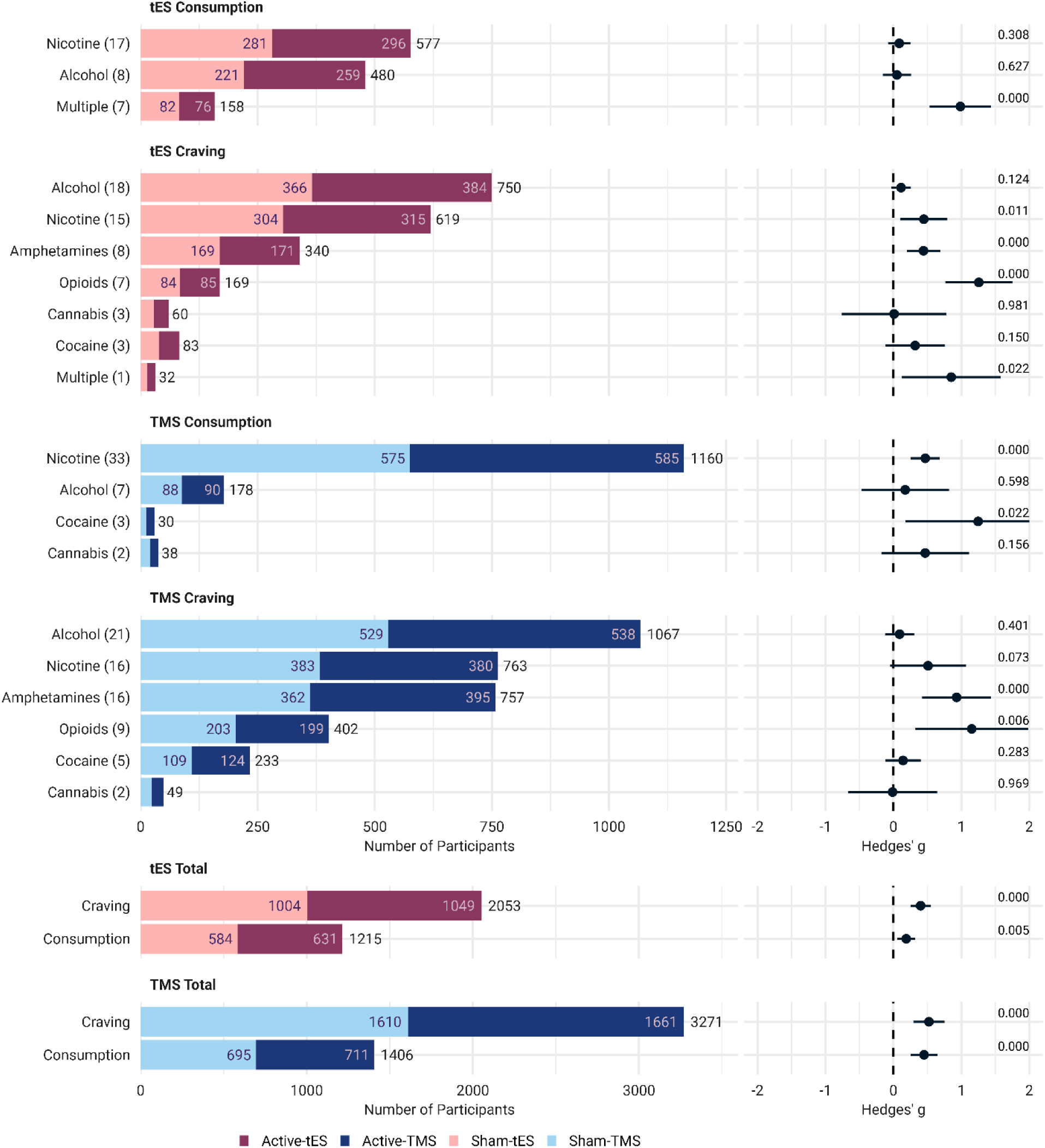
Exploring Overall Effects and Subgroup Analyses. This figure presents the results of a subgroup analysis on craving and consumption, featuring forest plots that detail the effect sizes by type of substance. These plots offer an in-depth look at the impact of tES/TMS on craving and consumption by examining the role of the type of stimulation. Each horizontal line within the plots represents a distinct substance use disorder, with dots for effect sizes and horizontal lines for confidence intervals. Additionally, bar plots show the number of participants in the active (dark colors) and sham (light colors) arms. The total number of experiments is noted in parentheses adjacent to the related moderator. The final four rows illustrate the cumulative effects of stimulation on craving and consumption, with bars color-coded by the type of stimulation: tES studies in red and TMS studies in blue. <u>Abbreviations</u>: tES: transcranial electrical stimulation, TMS: transcranial magnetic stimulation.

**Figure S.9-2.**
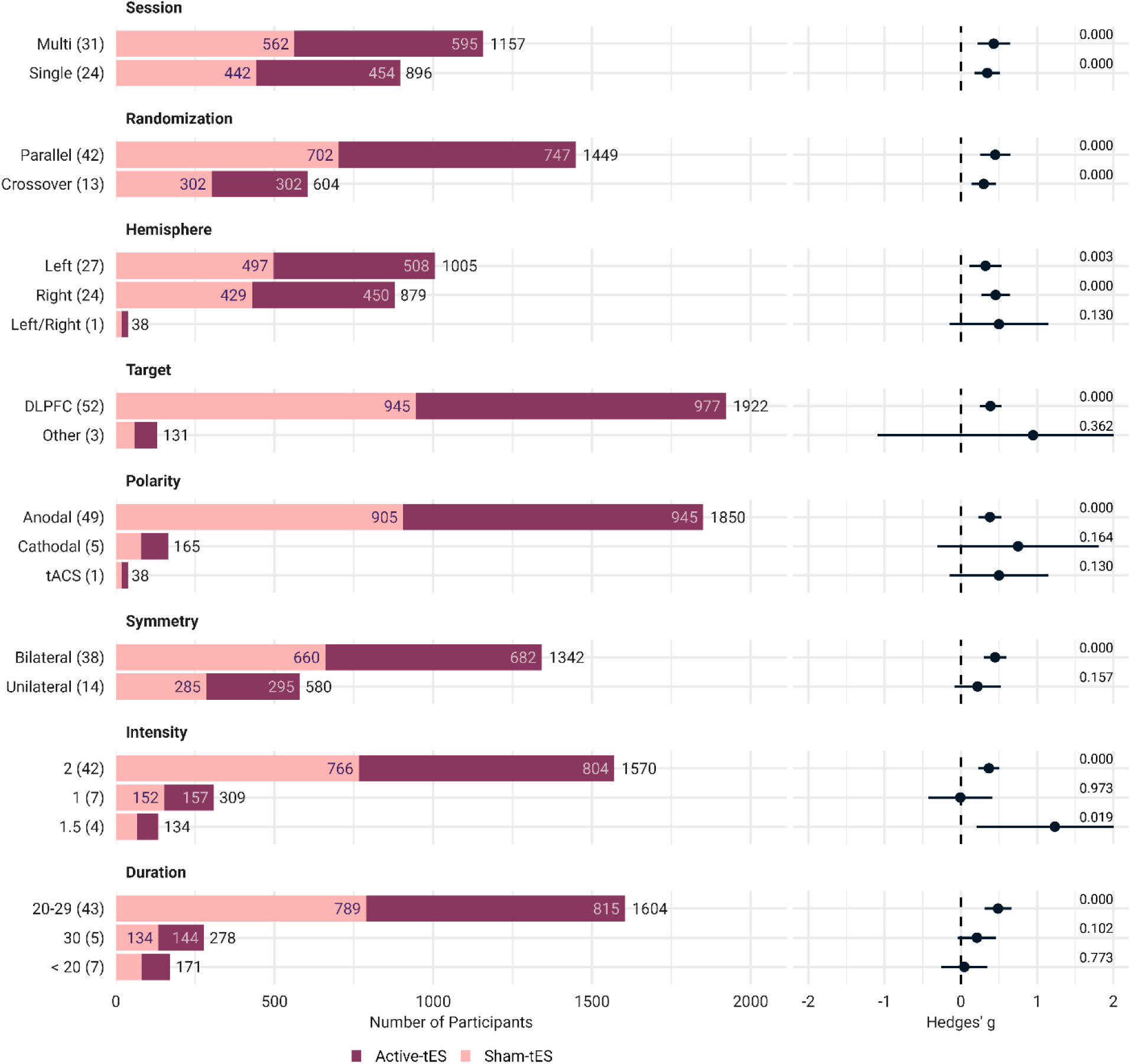
Exploring Subgroup Analyses of tES Effects on Craving. This figure displays the results of a subgroup analysis focusing on craving, featuring forest plots that illustrate the effect sizes by different types of moderators. These plots provide a comprehensive analysis of the impact of tES on craving, considering key factors such as number of sessions, randomization, hemisphere, target, polarity, symmetry in DLPFC stimulation (in minutes), intensity (in mA), and stimulation duration. Each horizontal line within the plots signifies a specific moderator, with dots representing effect sizes and horizontal lines denoting confidence intervals. In addition, bar plots indicate the number of participants in the active (dark colors) and sham (light colors) arms, with the total number of experiments mentioned in parentheses next to each relevant moderator. Note that two studies did not report tES intensity. The hemisphere subgroup pertains to studies targeting the DLPFC. The left/right refers to tACS studies. <u>Abbreviations:</u> tES: transcranial electrical stimulation, tACS: transcranial alternating stimulation, DLPFC: dorsolateral prefrontal cortex.

**Figure S.9-3.**
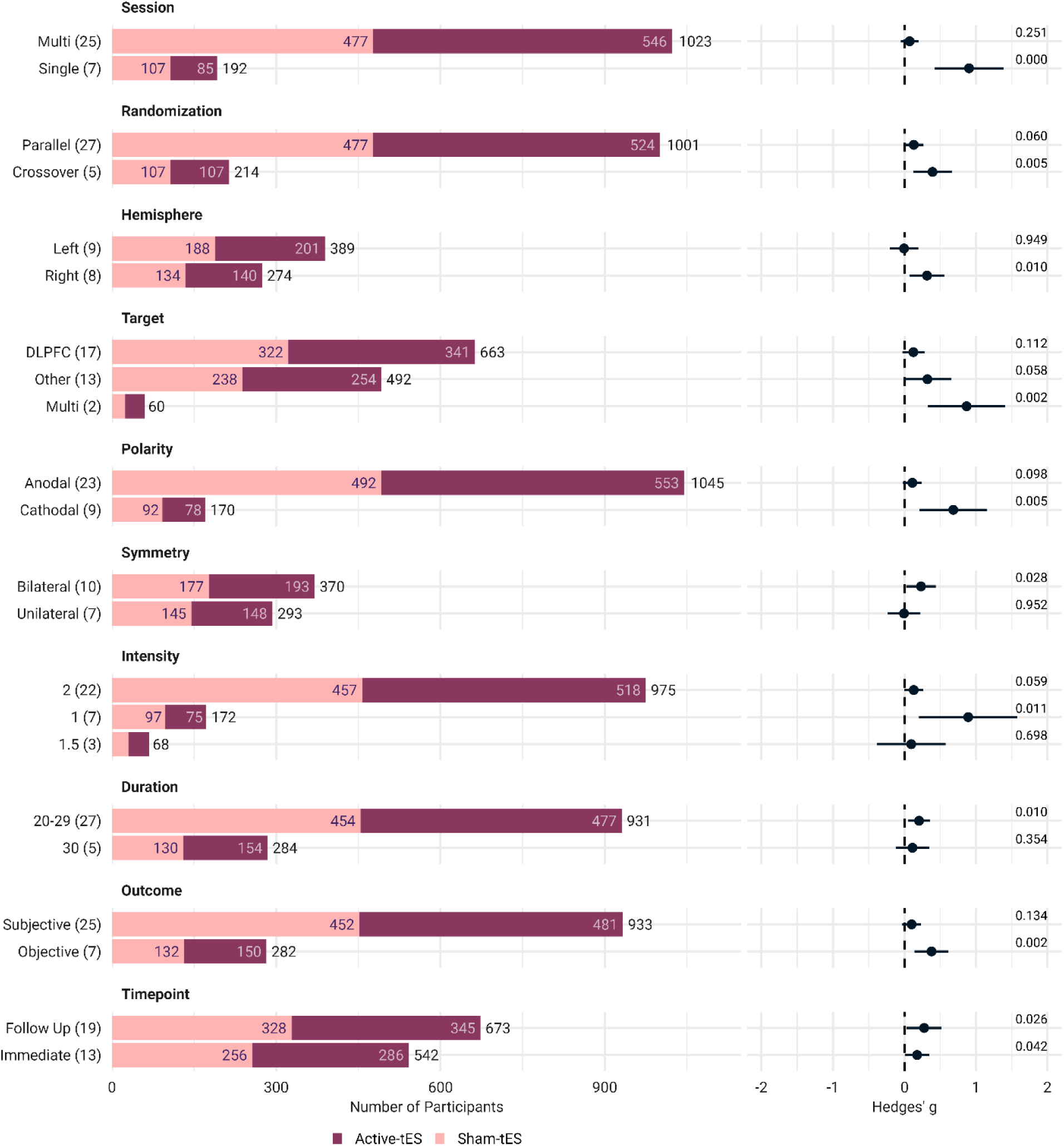
Exploring Subgroup Analyses of tES Effects on Consumption. This figure displays the results of a subgroup analysis focusing on consumption, featuring forest plots that illustrate the effect sizes by different types of moderators. These plots provide a comprehensive analysis of the impact of tES on consumption, considering key factors such as the type of outcome measurement, outcome assessment timepoint, number of sessions, randomization, hemisphere, target, polarity, symmetry in DLPFC stimulation, intensity, and stimulation duration. Each horizontal line within the plots signifies a specific moderator, with dots representing effect sizes and horizontal lines denoting confidence intervals. In addition, bar plots indicate the number of participants in the active (dark colors) and sham (light colors) arms, with the total number of experiments mentioned in parentheses next to each relevant moderator. The hemisphere subgroup pertains to studies targeting the DLPFC. <u>Abbreviations:</u> tES: transcranial electrical stimulation, DLPFC: dorsolateral prefrontal cortex.

**Figure S.9-4.**
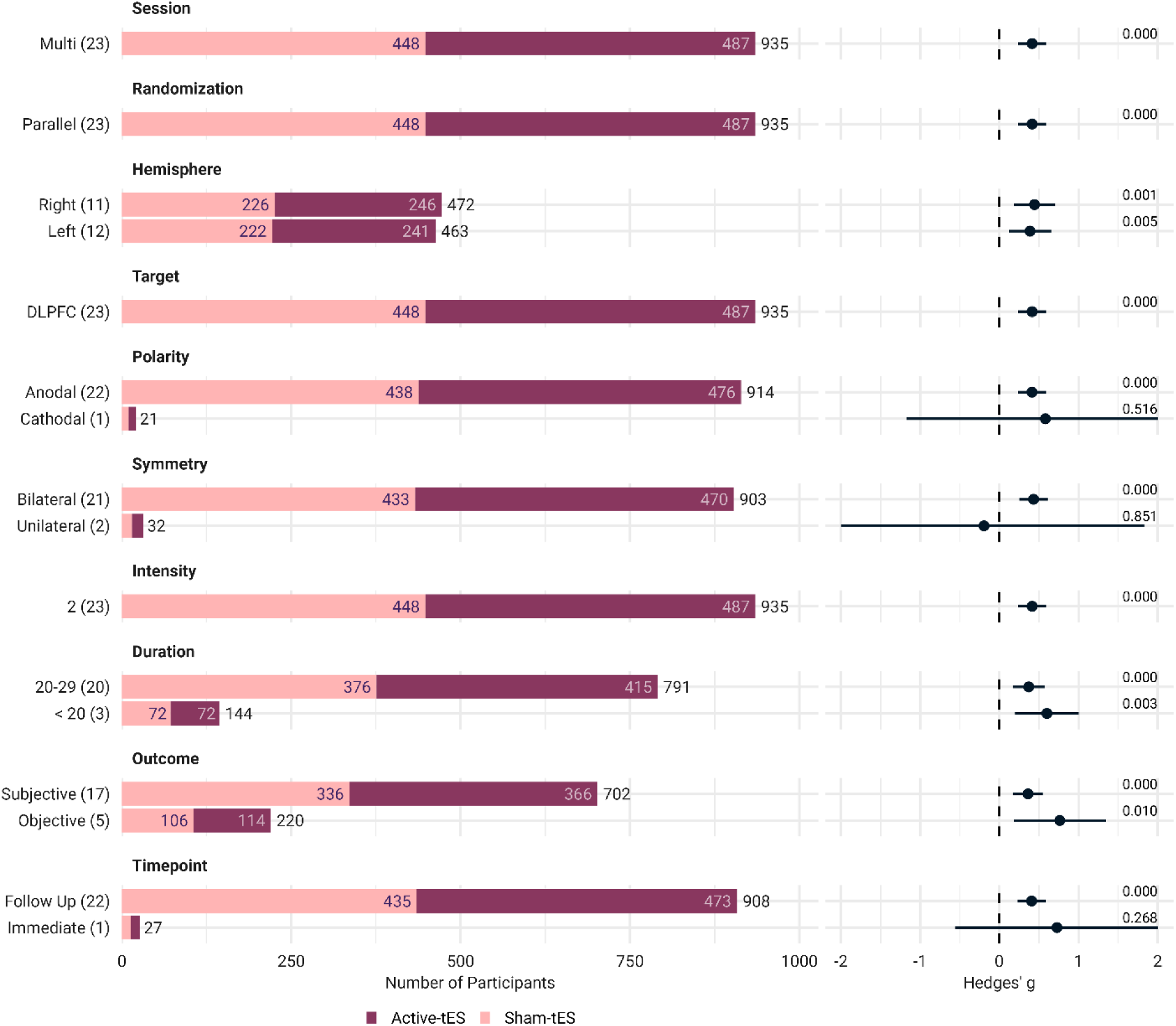
Exploring Subgroup Analyses of tES Effects on Relapse. This figure displays the results of a subgroup analysis focusing on relapse, featuring forest plots that illustrate the effect sizes by different types of moderators. These plots provide a comprehensive analysis of the impact of tES on relapse, considering key factors such as the type of outcome measurement, outcome assessment timepoint, number of sessions, randomization, hemisphere, target, polarity, symmetry in DLPFC stimulation, intensity, and stimulation duration. Each horizontal line within the plots signifies a specific moderator, with dots representing effect sizes and horizontal lines denoting confidence intervals. In addition, bar plots indicate the number of participants in the active (dark colors) and sham (light colors) arms, with the total number of experiments mentioned in parentheses next to each relevant moderator. The hemisphere subgroup pertains to studies targeting the DLPFC. <u>Abbreviations:</u> tES: transcranial electrical stimulation, DLPFC: dorsolateral prefrontal cortex.

**Figure S.9-5.**
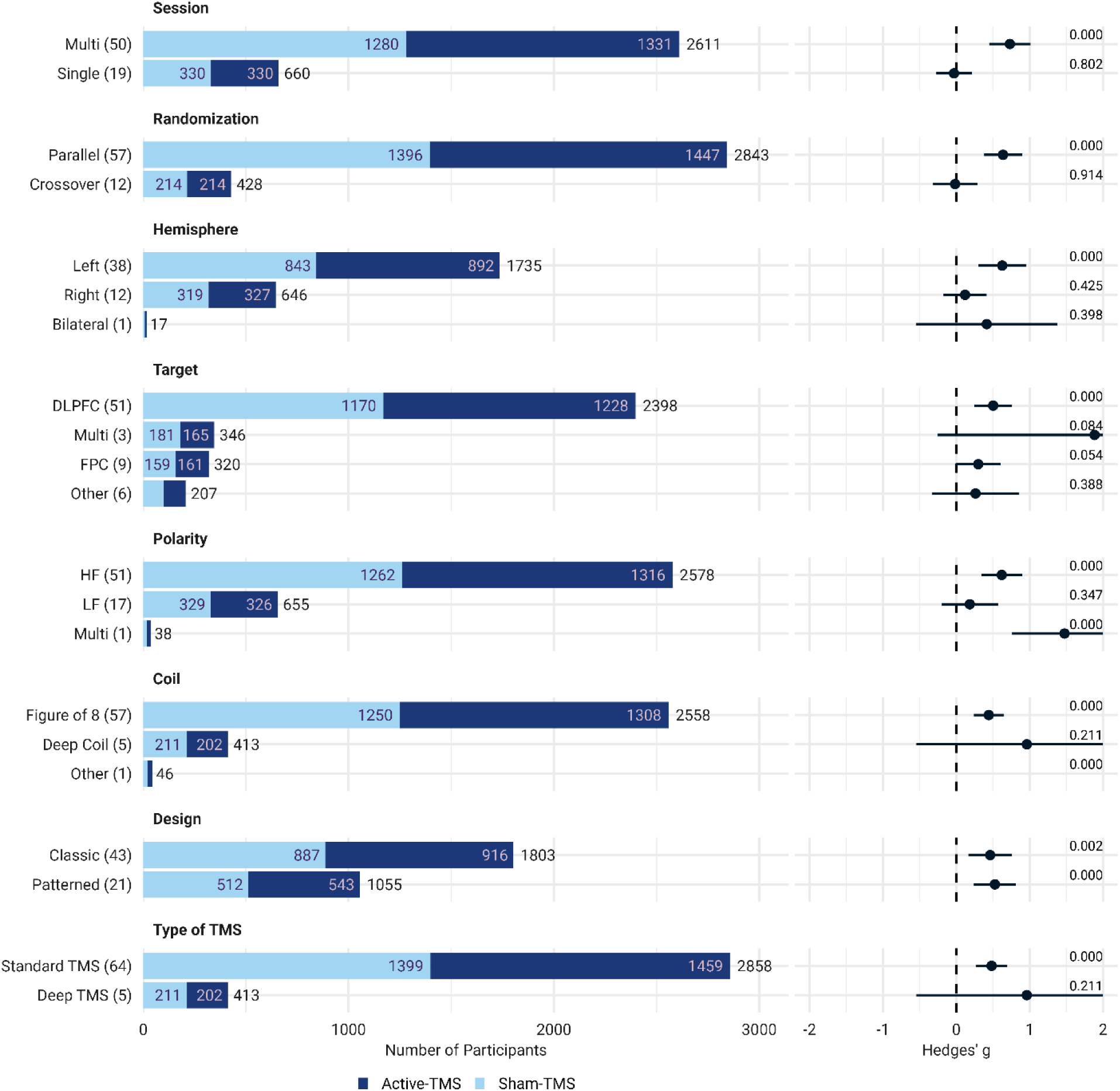
Exploring Subgroup Analyses of TMS Effects on Craving. This figure presents the results of a subgroup analysis focused on craving, featuring forest plots that demonstrate the effect sizes across various moderators. These plots offer a thorough analysis of TMS’s impact on craving, considering key factors such as number of sessions, randomization, hemisphere in DLPFC stimulation, target, frequency, coil type, stimulation patterns, and type of TMS. ’Multi’ in the ’Target’ group refers to studies that employed a combination of different target areas. ’Multi’ in the ’Polarity’ group refers to studies that employed both HF and LF TMS simultaneously. Each horizontal line in the plots represents a specific moderator, with dots indicating effect sizes and horizontal lines showing confidence intervals. Additionally, bar plots display the number of participants in the active (dark colors) and sham (light colors) arms, with the total number of experiments indicated in parentheses alongside each moderator. Note that six studies did not report the type of TMS coil. The hemisphere subgroup pertains to standard TMS (classic or patterned) studies targeting the DLPFC. <u>Abbreviations</u>: TMS: transcranial magnetic stimulation, HF: high frequency, LF: low frequency, FPC: frontopolar cortex, DLPFC: dorsolateral prefrontal cortex.

**Figure S.9-6.**
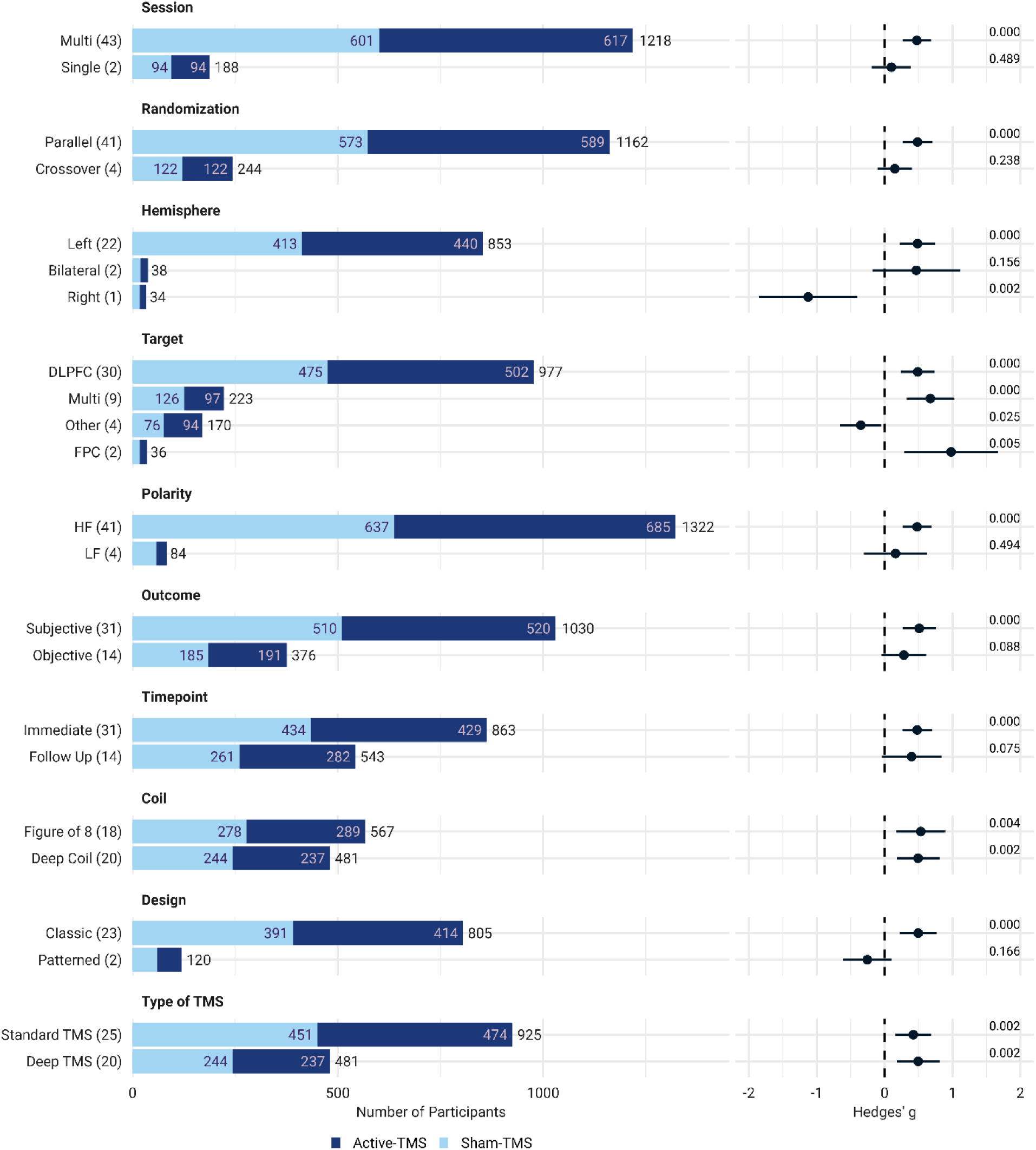
Exploring Subgroup Analyses of TMS Effects on Consumption. This figure presents the results of a subgroup analysis focused on consumption, featuring forest plots that demonstrate the effect sizes across various moderators. These plots offer a thorough analysis of the impact of TMS on consumption, taking into account key factors such as the type of outcome measurement, outcome assessment timepoint, number of sessions, randomization, hemisphere in DLPFC stimulation, target, frequency, coil type, stimulation patterns, and type of TMS. Each horizontal line in the plots represents a specific moderator, with dots indicating effect sizes and horizontal lines showing confidence intervals. Additionally, bar plots display the number of participants in the active (dark colors) and sham (light colors) arms, with the total number of experiments indicated in parentheses alongside each moderator. The hemisphere subgroup pertains to standard TMS (classic or patterned) studies targeting the DLPFC. <u>Abbreviations</u>: TMS: transcranial magnetic stimulation, HF: high frequency, LF: low frequency, FPC: frontopolar cortex, DLPFC: dorsolateral prefrontal cortex.

**Figure S.9-7.**
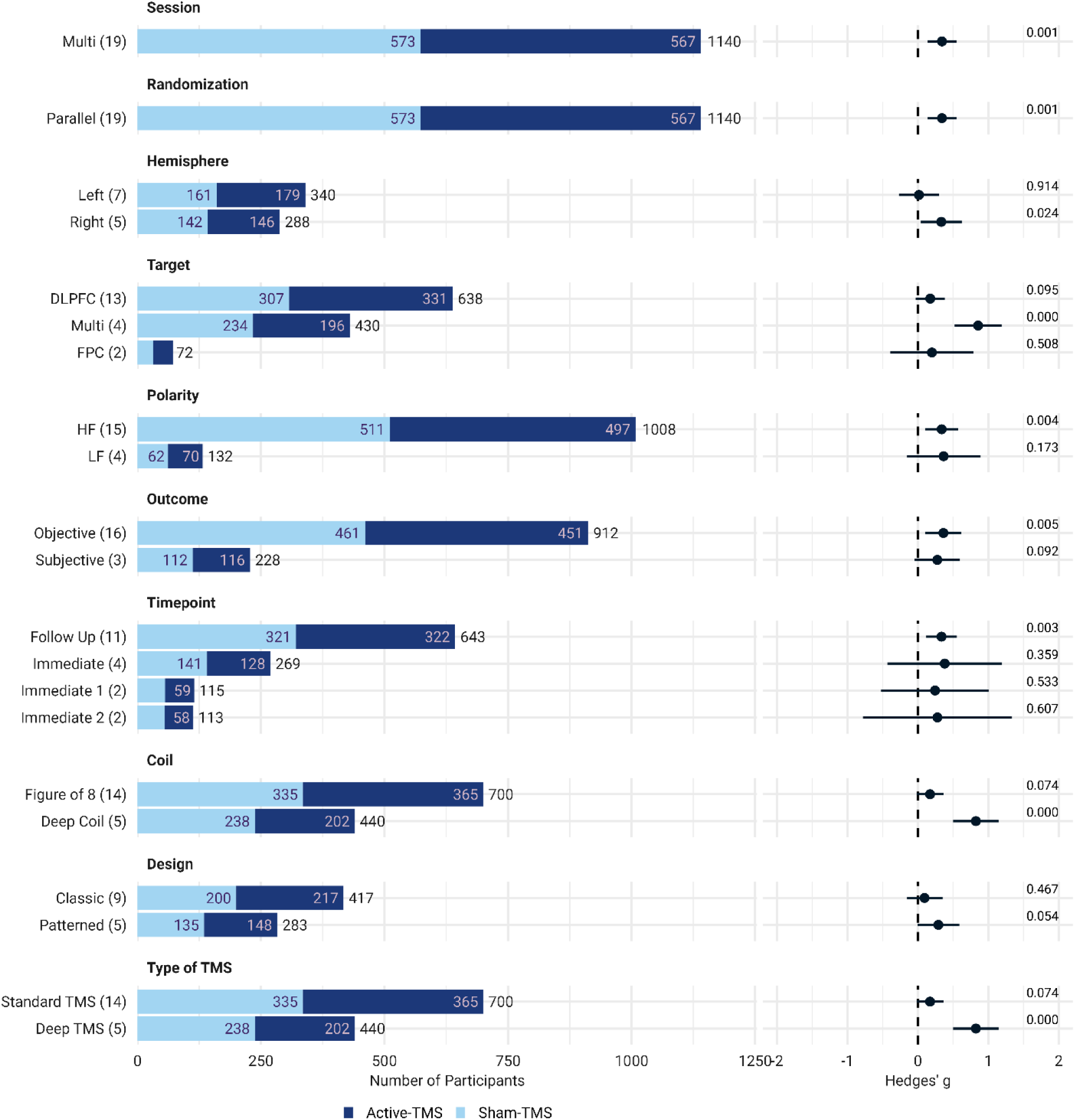
Exploring Subgroup Analyses of TMS Effects on Relapse. This figure presents the results of a subgroup analysis focused on relapse, featuring forest plots that demonstrate the effect sizes across various moderators. These plots offer a thorough analysis of the impact of TMS on relapse, taking into account key factors such as type of outcome measurement, outcome assessment timepoint, number of sessions, randomization, hemisphere in DLPFC stimulation, target, frequency, coil type, stimulation patterns, and type of TMS. Each horizontal line in the plots represents a specific moderator, with dots indicating effect sizes and horizontal lines showing confidence intervals. Additionally, bar plots display the number of participants in the active (dark colors) and sham (light colors) arms, with the total number of experiments indicated in parentheses alongside each moderator. The hemisphere subgroup pertains to standard TMS (classic or patterned) studies targeting the DLPFC. <u>Abbreviations</u>: TMS: transcranial magnetic stimulation, HF: high frequency, LF: low frequency, FPC: frontopolar cortex, DLPFC: dorsolateral prefrontal cortex.

**Figure S.9-8.**
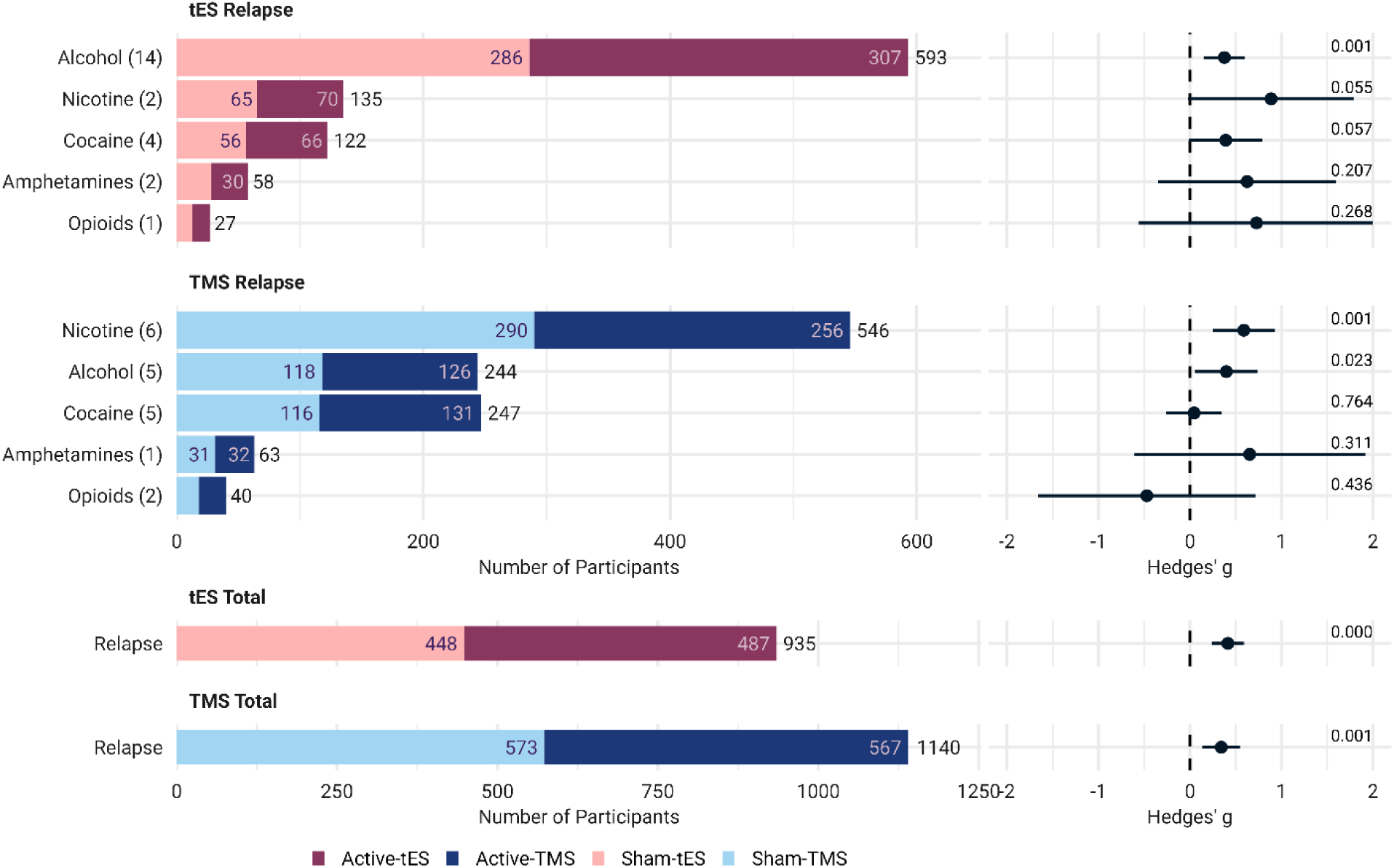
Exploring Subgroup Analyses of Substance on Relapse for tES/TMS. This figure presents the results of a subgroup analysis on relapse, featuring forest plots that detail the effect sizes by type of substance. These plots offer an in-depth look at the impact of tES/TMS on relapse by examining the role of the type of stimulation. Each horizontal line within the plots represents a distinct substance use disorder, with dots for effect sizes and horizontal lines for confidence intervals. Additionally, bar plots show the number of participants in the active (dark colors) and sham (light colors) arms. The total number of experiments is noted in parentheses adjacent to the related moderator. The final two rows illustrate the cumulative effects of stimulation on relapse, with bars color-coded by the type of stimulation: tES studies in red and TMS studies in blue. <u>Abbreviations</u>: tES: transcranial electrical stimulation, TMS: transcranial magnetic stimulation.

### Heterogeneity of craving and consumption measures in the meta-analysis

**Figure S.10-1.**
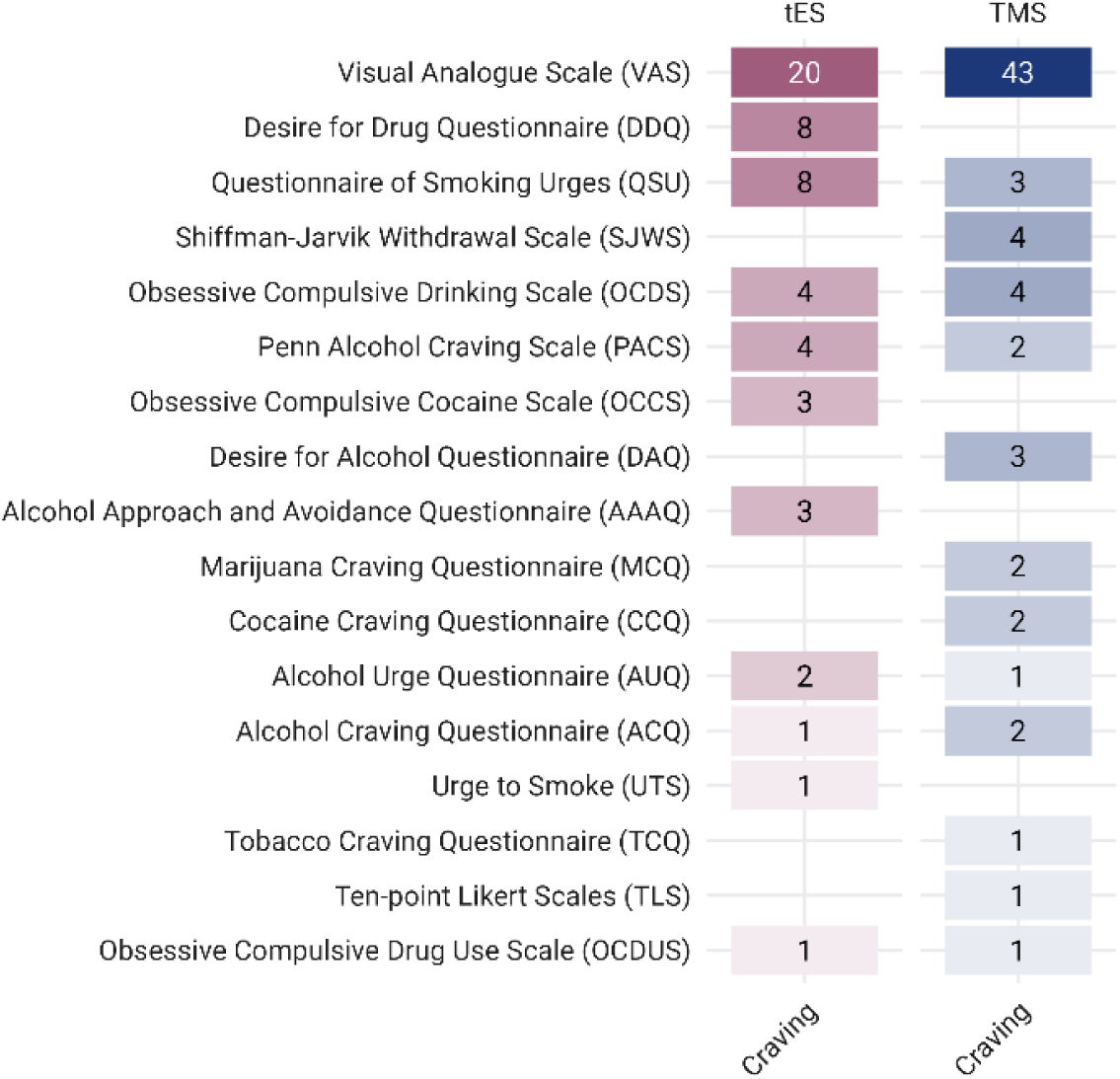
Distribution of Craving Scales Used in tES and TMS Studies Included in the Meta-Analysis. This figure illustrates the various scales included in the meta-analysis, color-coded by the type of stimulation—tES in red and TMS in blue—and categorized by outcome measure (craving). <u>Abbreviations</u>: tES: transcranial electrical stimulation, TMS: transcranial magnetic stimulation.

**Figure S.10-2.**
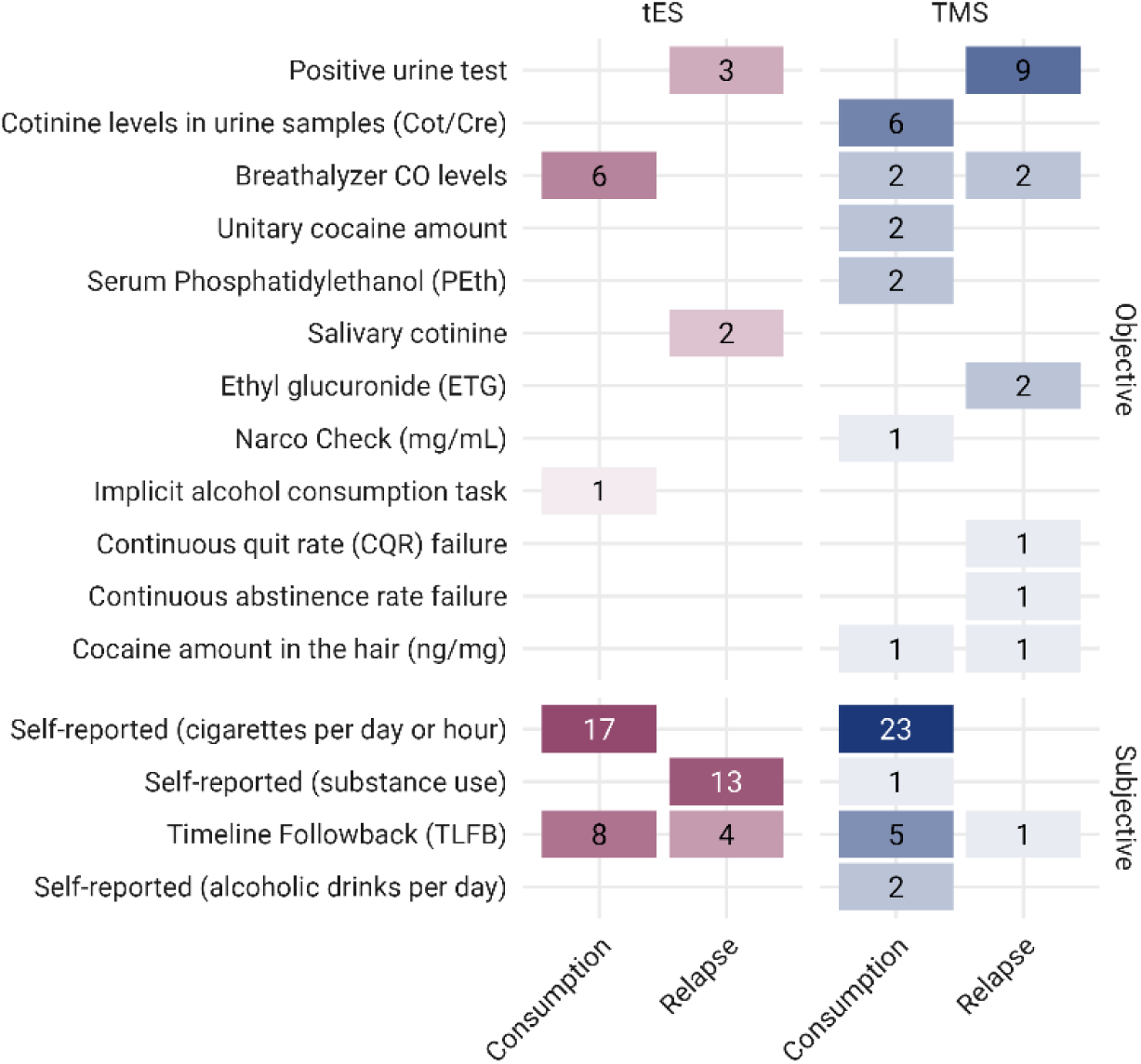
Consumption/Relapse Outcome Measures Used in tES and TMS Studies Included in the Meta-Analysis. This figure illustrates the various scales used in the meta-analysis, color-coded by the type of stimulation—tES in red and TMS in blue—and categorized by outcome measure (consumption and relapse). <u>Abbreviations</u>: tES: transcranial electrical stimulation, TMS: transcranial magnetic stimulation, ng/mg: nanogram per milligram, mg/mL: milligrams per milliliter, CO: carbon monoxide, Cot/Cre: cotinine-creatinine ratio.

### All available outcome measures

**Table S.4.**
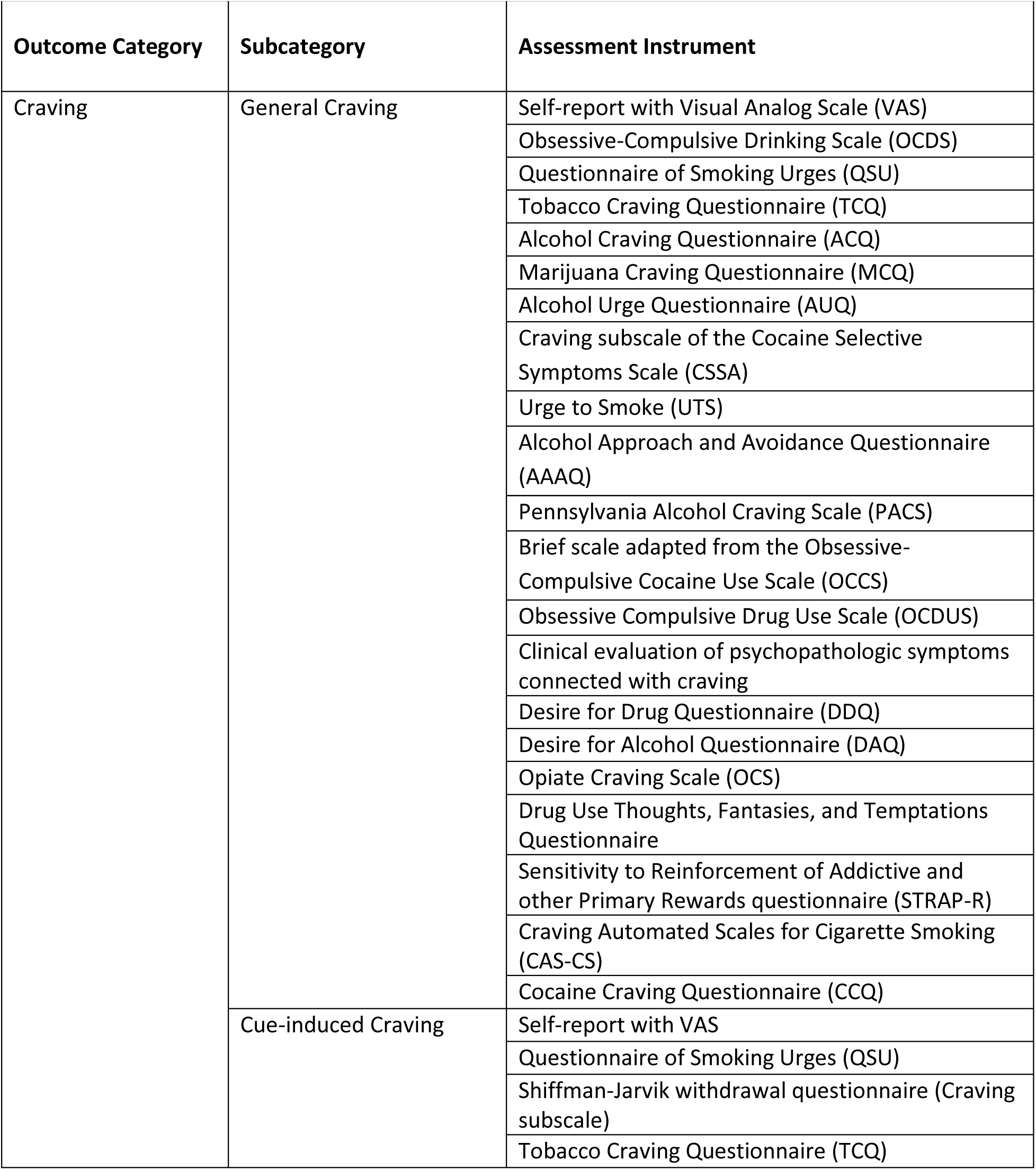
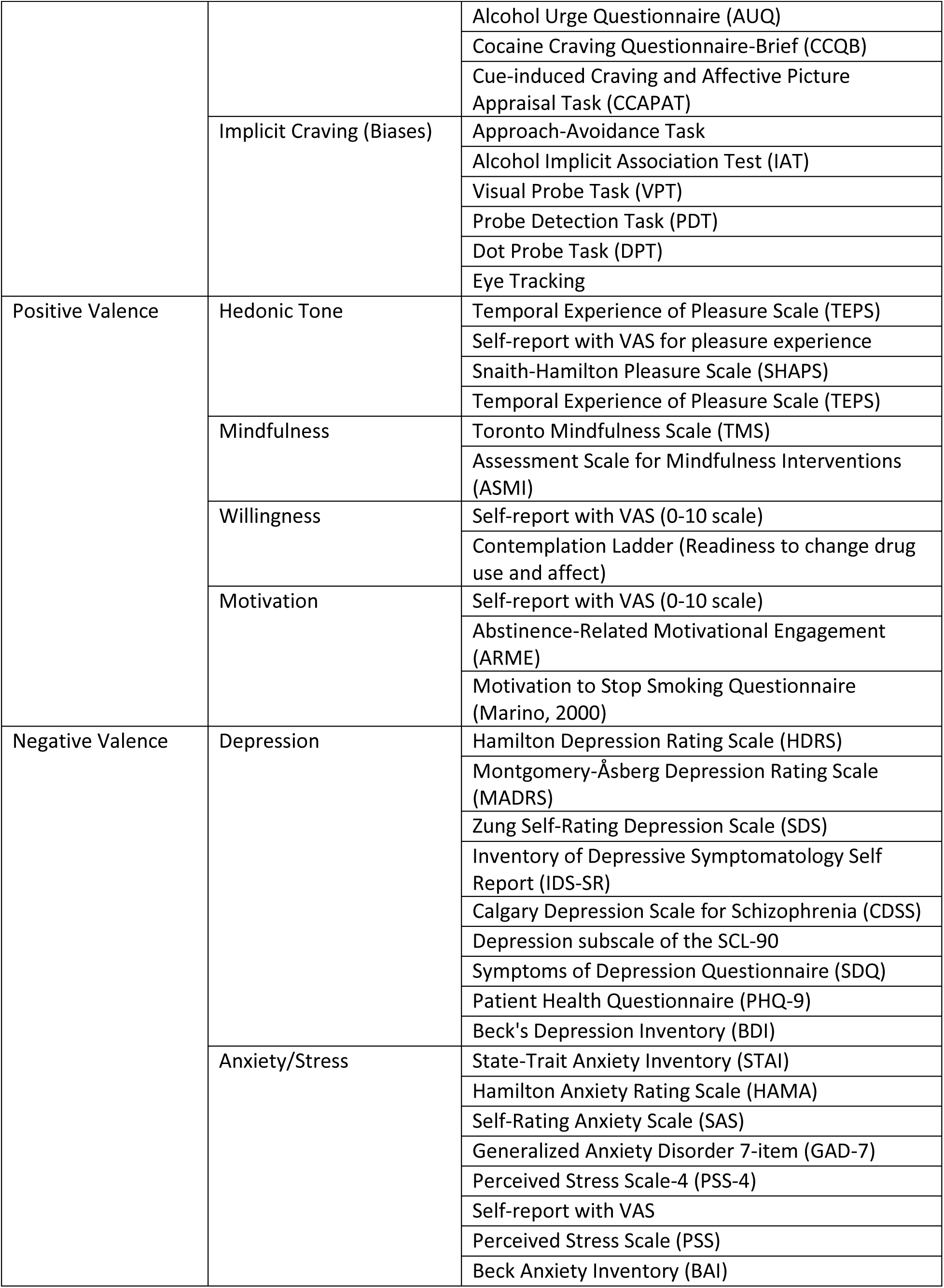
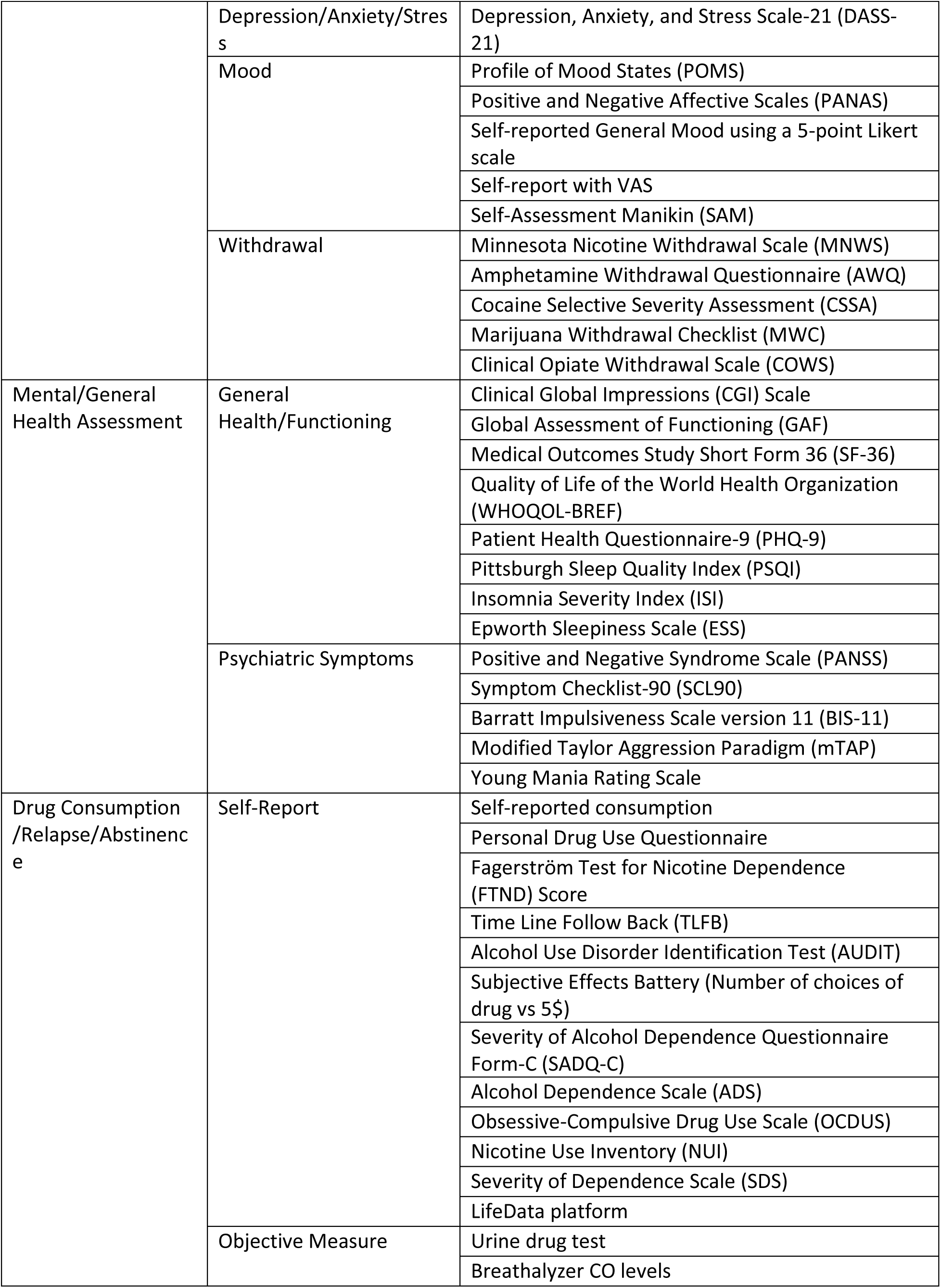
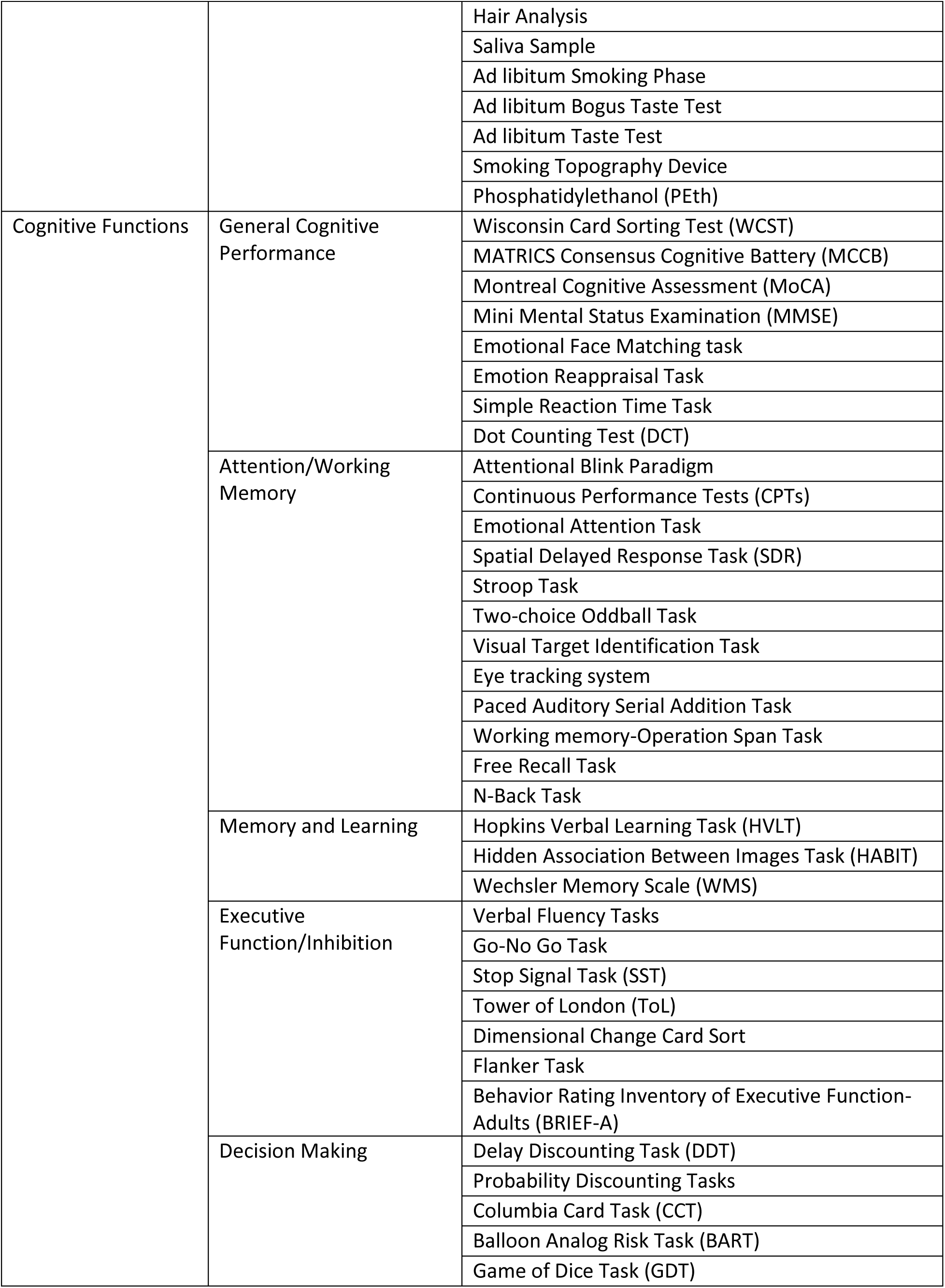
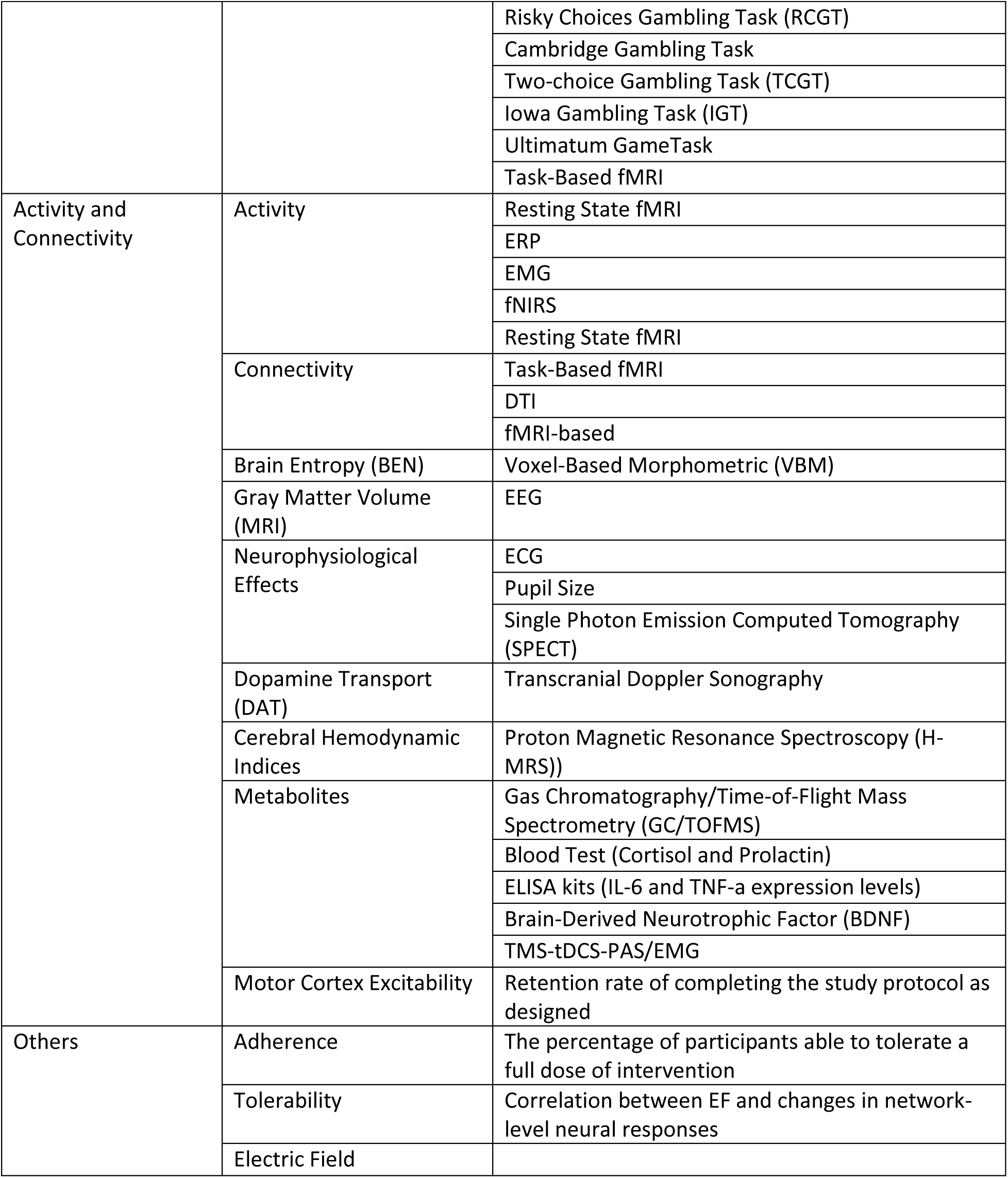
Outcome measures for tES/TMS studies for addiction medicine treatment.

